# ATN Classification and Machine-Learned Plasma Biomarker Phenotypes Reveal Distinct Alzheimer’s Pathology in a Population-Based Cohort

**DOI:** 10.64898/2026.01.02.26343331

**Authors:** Emmanuel Fle Chea

## Abstract

**Background:** The ATN (Amyloid/Tau/Neurodegeneration) framework provides a theory-driven approach to Alzheimer’s disease (AD) classification using binary biomarker cutoffs, while unsupervised machine learning offers data-driven phenotyping. The concordance between these approaches in population-representative samples remains incompletely characterized.

**Objective:** To compare plasma ATN classification with data-driven clustering methods and evaluate their associations with cognitive outcomes in a nationally representative cohort.

**Methods:** We analyzed plasma biomarkers (Aβ42/40 ratio, p-tau181, NfL, GFAP) from 4,465 participants aged ≥51 years in the Health and Retirement Study 2016 Venous Blood Study. ATN profiles were classified using literature-based cutoffs. We applied k-means clustering, Gaussian mixture modeling, and variational autoencoder (VAE) dimensionality reduction to identify data-driven biomarker-defined subgroups. Agreement between ATN and clustering was quantified using adjusted Rand index (ARI) and normalized mutual information (NMI). Longitudinal analyses examined associations with cognitive decline over 4 years (2016-2020).

**Results:** The analytic sample included 4,465 individuals (mean age 69.7±10.4 years; 58.7% female; 75.8% non-Hispanic White). ATN classification yielded 14 profiles, with A+/T-/N-(27.4%) and A-/T-/N-(22.6%) most prevalent (Figure 2). K-means clustering identified 4 optimal clusters with distinct biomarker signatures. Agreement between ATN and clusters was modest (ARI=0.119, NMI=0.113). Sensitivity analysis excluding GFAP from clustering reduced agreement substantially (ARI=0.03 vs 0.119 with GFAP, -74.5% decrease), demonstrating that GFAP accounts for most of the observed concordance between clustering and ATN classification, with only one-third arising from the shared three biomarkers.[Table S12] Additional sensitivity analyses confirmed that k=4 provides finer biomarker resolution than k=3 by retaining biomarker-extreme subgroups[Table S13], and that Cluster 4 represents a stable biological structure across distance metrics[Table S14] despite its small size. Cluster 1 (n=51, 1.2%) showed severe pathology; Cluster 3 (n=3,479, 78.6%) represented the largest and most heterogeneous group, encompassing the broad spectrum of minimal to moderate pathology across all ATN profiles; Cluster 4 (n=14, 0.3%) represented a small but stable non-AD biomarker-defined subgroup (Jaccard=0.779). The VAE revealed a localized nonlinear structure. Silhouette values in the latent space are not directly comparable to clustering silhouettes, but the VAE embedding showed clearer local separation, whereas PCA explained more variance (67.1%). Both ATN and clusters predicted 4-year cognitive decline (ATN R²=0.024, p<0.001; Clusters R²=0.019, p<0.001).

**Conclusions:** Theory-driven ATN classification and data-driven biomarker phenotyping capture partially overlapping but largely distinct information. Modest concordance (ARI=0.119) reflects GFAP’s contribution to shared structure, with most alignment arising from GFAP rather than from the three ATN biomarkers alone (ARI=0.03). The primary source of discordance remains the binary versus continuous representation of biomarker variation. Sensitivity analyses showed that k=4 provides finer biomarker resolution than k=3, and that Cluster 4 represents a small but reproducible biomarker-defined subgroup. Both approaches predicted cognitive decline with modest effect sizes (R²=1.9**-**2.4%), consistent with population-based studies. Integrating theory-driven and data-driven frameworks may support a more comprehensive characterization of AD-related pathology in population research.

## 1. INTRODUCTION

Alzheimer’s disease (AD) represents a major public health challenge, affecting over 6 million Americans and imposing substantial socioeconomic burdens.[1,2] The development of blood-based biomarkers has revolutionized AD research by enabling large-scale population screening and reducing reliance on invasive cerebrospinal fluid (CSF) sampling or expensive neuroimaging.[3–5,52,60,62] Within this context, two fundamentally different approaches have emerged for characterizing AD-related pathology: theory-driven classification frameworks and data-driven phenotyping methods.

The ATN (Amyloid/Tau/Neurodegeneration) research framework, proposed by the National Institute on Aging and Alzheimer’s Association, provides a biologically grounded classification system.[6,7] This framework categorizes individuals using binary biomarker designations, A for amyloid-β (positive/negative), T for tau pathology (positive/negative), and N for neurodegeneration (positive/negative), yielding eight possible profiles. While ATN has gained widespread adoption in clinical research and trial enrichment strategies[8,9], the framework relies on predetermined cutoff thresholds that dichotomize continuous biomarker measurements, potentially obscuring biologically relevant subthreshold variation.[10,11]

Complementing this theory-driven approach, unsupervised machine learning techniques offer data-driven methods for discovering latent biomarker phenotypes without imposing a priori classifications.[12–14,64] Methods such as k-means clustering, Gaussian mixture models (GMMs), and deep learning architectures like variational autoencoders (VAEs) can identify natural groupings in high-dimensional biomarker data[66], potentially revealing heterogeneous pathological processes not captured by binary frameworks.[12–14,67,69] These approaches have successfully identified AD subtypes with distinct progression patterns and treatment responses in clinical cohorts.[15–19]

Despite their complementary nature, systematic comparisons between ATN classification and data-driven phenotyping remain limited, particularly in population-representative samples. Most existing studies focus on clinic-based cohorts enriched for cognitive impairment[20–21], which may not reflect the broader spectrum of AD-related pathology in community-dwelling older adults. Furthermore, the sensitivity of ATN classification to cutoff selection, a critical methodological consideration, has received insufficient attention in plasma biomarker research.[22–23]

The Health and Retirement Study (HRS) 2016 Venous Blood Study provides a unique opportunity to address these knowledge gaps.[24–25] As a nationally representative longitudinal survey of Americans over age 50, HRS includes diverse sociodemographic groups often underrepresented in traditional AD research.[26–27] The 2016 wave collected venous blood samples from participants for comprehensive biomarker assessment, including plasma markers of amyloid, tau, and neurodegeneration. Similar to the HRS, large population-based cohorts such as the Atherosclerosis Risk in Communities (ARIC) study have demonstrated the utility of plasma biomarkers for predicting incident dementia in community-dwelling adults.[61]

In this study, we systematically compare plasma ATN classification with multiple data-driven approaches, k-means clustering, GMMs, and VAE-based dimensionality reduction, in the HRS 2016 cohort. We hypothesize that (1) theory-driven ATN and data-driven clustering will show modest concordance, reflecting partially overlapping biological information with distinct contributions from binary categorization and continuous gradients; (2) ATN classification will demonstrate sensitivity to cutoff selection, with 10-20% reclassification under alternative thresholds; (3) data-driven methods will reveal heterogeneous biomarker profiles crossing traditional ATN boundaries, including rare phenotypes like non-AD neurodegeneration; and (4) GFAP’s inclusion in clustering but exclusion from ATN will contribute meaningfully to discordance, quantifiable through sensitivity analyses. Additionally, we examine longitudinal associations between classification approaches and cognitive decline over 4 years, contextualizing modest effect sizes within the multifactorial nature of population-based cognitive aging. By elucidating the relationship between these approaches in a population-representative context, this work informs strategies for precision characterization of AD-related pathology in epidemiological research and public health applications.

## 2. METHODS

### 2.1 Study Population

Data were drawn from the HRS, a nationally representative longitudinal panel study of Americans over age 50 conducted biennially since 1992.[24] The HRS employs a multistage area probability sample design with oversampling of Black and Hispanic households. In 2016, HRS implemented the Venous Blood Study (VBS), collecting blood samples from consenting respondents during enhanced face-to-face interviews.[25] Participants provided written informed consent for biospecimen collection and analysis. The study was approved by the University of Michigan Institutional Review Board. All research procedures were conducted in accordance with the Declaration of Helsinki and followed institutional guidelines for the ethical treatment of human participants.

#### 2.1.1 Population Representativeness and Generabilizability

To ensure our findings generalize beyond the analytic sample, we first characterize the representativeness of the HRS cohort. The HRS sampling design ensures national representativeness through multistage area probability sampling with oversampling of Black and Hispanic households, yielding a cohort that reflects the demographic and socioeconomic diversity of community-dwelling older Americans. Survey weights (PVBSWGTR) account for differential selection probabilities and non-response, enabling population-level inference. Importantly, our analytic sample’s unweighted demographic composition closely approximated weighted population estimates (Supplementary Table S9), with differences <1% for most characteristics, supporting the generalizability of unweighted cluster assignments. While HRS is U.S.-specific and excludes institutionalized individuals, its large sample size (N=4,465), racial/ethnic diversity (24.2% non-White), and rigorous longitudinal design provide robust external validity for characterizing plasma biomarker phenotypes in community-based aging populations.

Of 4,539 HRS participants with plasma biomarkers measured in 2016, 4,465 had complete data on all four biomarkers (NfL, GFAP, Aβ42/40 ratio, p-tau181) and cognitive assessment from the 2016 core interview. This constituted our primary analytic sample. Longitudinal cognitive data were available for 3,862 participants in 2018 and 3,524 participants in 2020. A detailed sample derivation flowchart is provided in Figure 1.

**FIGURE 1.**
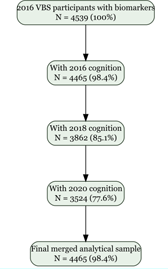
Study Sample Derivation Flowchart. CONSORT-style flowchart showing derivation of the analytic sample from the Health and Retirement Study 2016 Venous Blood Study. Beginning with 4,539 HRS participants with plasma biomarkers measured, 4,465 had complete data on all four biomarkers (NfL, GFAP, Aβ42/40 ratio, p-tau181) and cognitive assessment from the 2016 core interview, constituting the primary analytic sample for ATN classification. For clustering analyses requiring complete covariate data, 38 participants were excluded, yielding n=4,427. Longitudinal cognitive data were available for 3,862 participants in 2018 (14% attrition) and 3,524 participants in 2020 (21% attrition from baseline). The primary source of attrition from VBS participants (n=4,539) to the analytic sample (n=4,465) was individuals with incomplete biomarker panels (n=74). **Abbreviations:** Aβ, amyloid-β; GFAP, glial fibrillary acidic protein; HRS, Health and Retirement Study; NfL, neurofilament light; VBS, Venous Blood Study.

**FIGURE 2.**
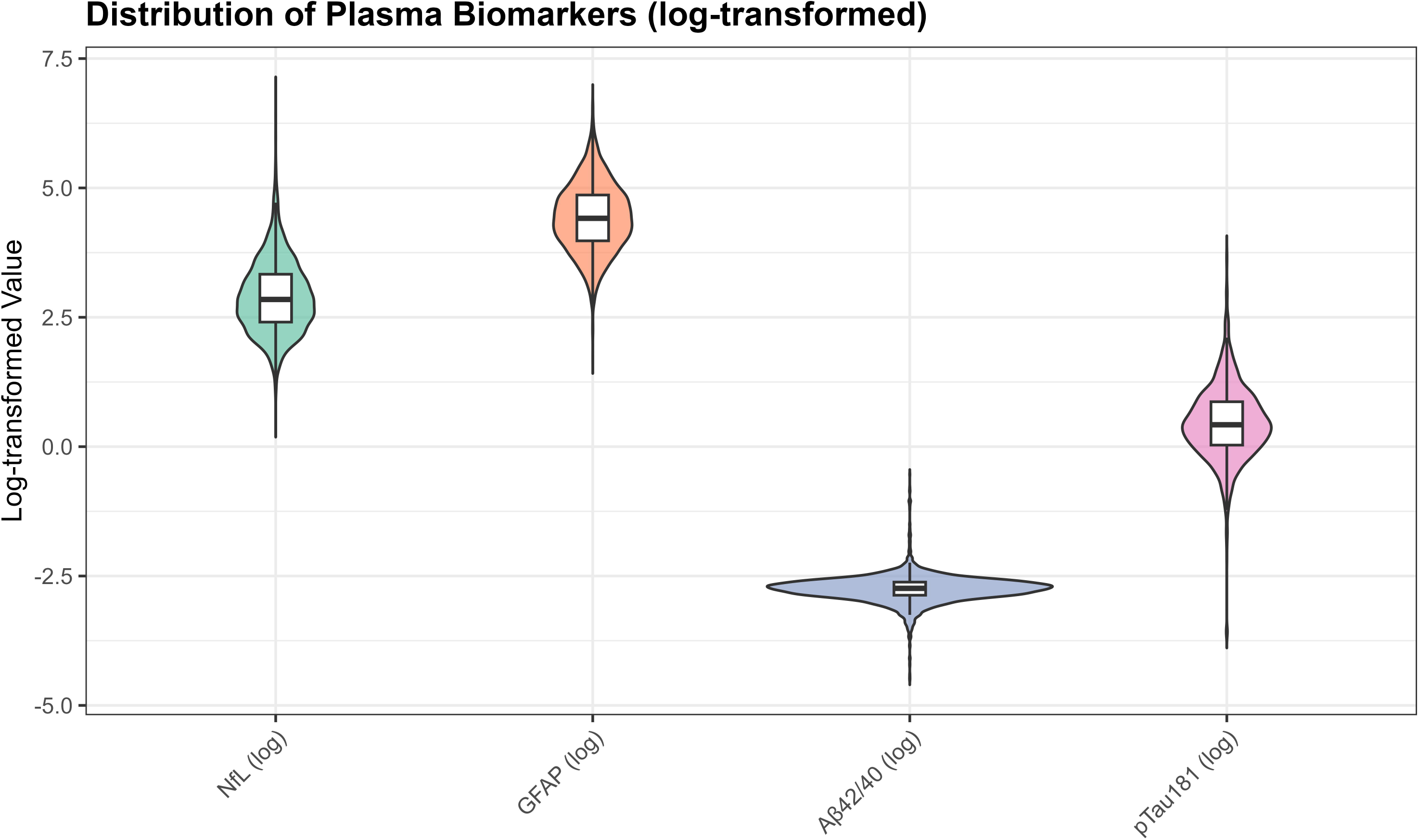
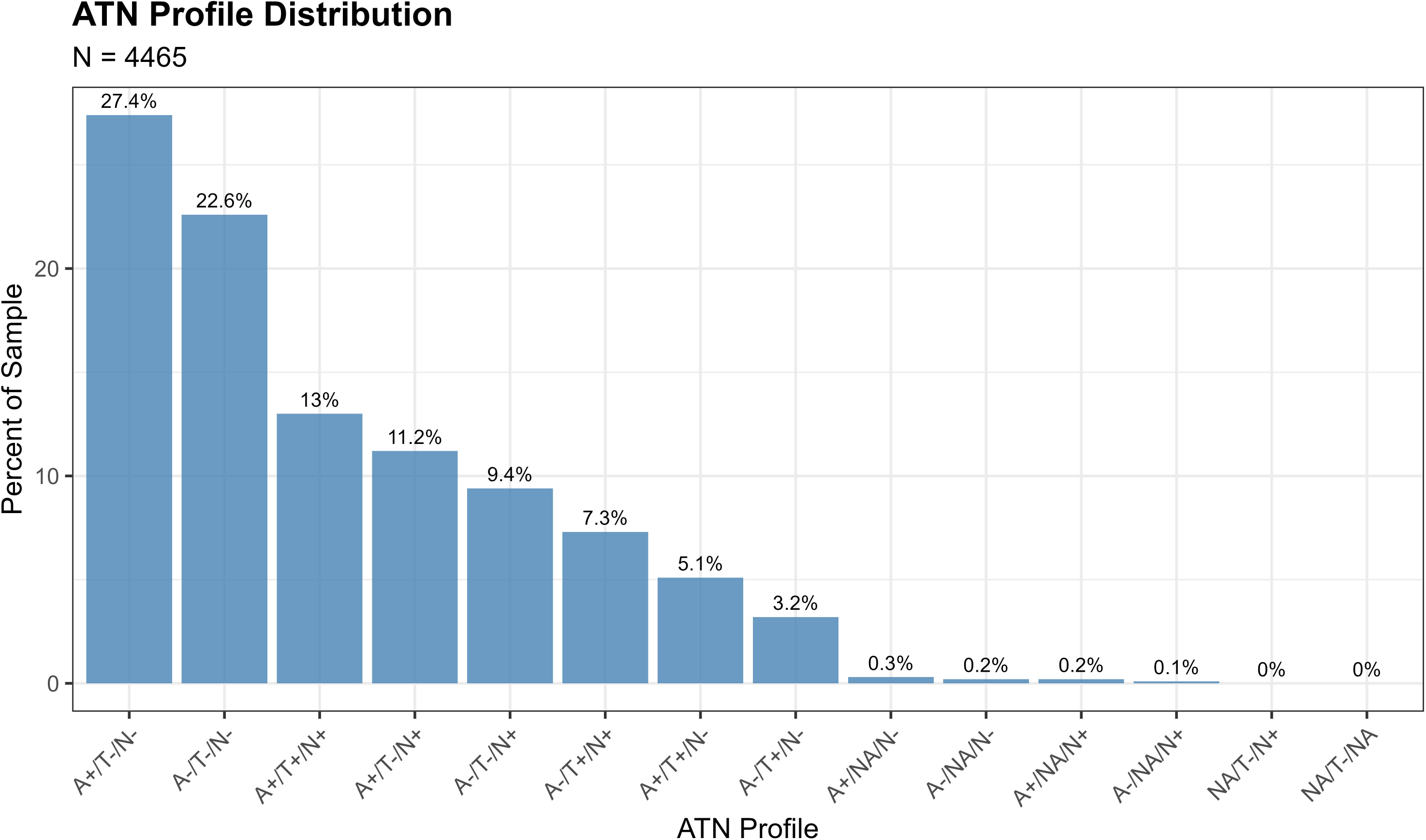
Plasma Biomarker Distributions and ATN Profile Prevalence. **Panel A:** Violin plots showing the distribution of log-transformed plasma biomarkers across the entire sample (N=4,465). Each violin displays the probability density of values, with embedded box plots indicating median (center line), interquartile range (box), and 1.5×IQR (whiskers). From left to right: NfL (log): Median=2.996 (equivalent to 20.0 pg/mL); GFAP (log): Median=5.095 (equivalent to 163.2 pg/mL); Aβ42/40 ratio (log): Median=**-**2.900 (equivalent to 0.055); p-Tau181 (log): Median=0.788 (equivalent to 2.2 pg/mL). All biomarkers show right-skewed distributions in the original scale, with log-transformation achieving approximate normality suitable for clustering analyses (Q-Q plots in Supplementary Figure S1). **Panel B:** Bar chart showing the distribution of ATN profiles in the sample. The x-axis displays 14 observed ATN profiles ordered by prevalence; y-axis shows percentage of sample. The most common profile is A+/T-/N-(27.4%, n=1,224), followed by A-/T-/N-(22.6%, n=1,009), A+/T+/N+ (13.0%, n=579), A+/T-/N+ (11.2%, n=500), A-/T+/N+ (9.4%, n=419), A-/T-/N+ (7.3%, n=325), A+/T+/N-(5.1%, n=227), and A-/T+/N-(3.2%, n=144). Rare profiles (A+/NA/N+, A-/NA/N+, A+/NA/N-, NA/T-/N+, NA/T-/NA) each comprise <1% of the sample. **Abbreviations:** Aβ, amyloid-β; GFAP, glial fibrillary acidic protein; IQR, interquartile range; NfL, neurofilament light.

### 2.2 Biomarker Assessment

#### 2.2.1 Blood Collection and Processing

Venous blood samples were collected by trained phlebotomists within 4 weeks of the core interview, following standardized protocols.[25] Samples were shipped overnight at ambient temperature to designated laboratories for processing within 36 hours. Plasma was separated by centrifugation and aliquoted for long-term storage at -80°C until assay.

#### 2.2.2 Plasma Biomarker Assays

Four plasma biomarkers were measured using validated immunoassays:

**Neurofilament Light Chain (NfL):** Quantified using the Simoa NF-Light Advantage Kit (Quanterix Corporation, Billerica, MA) on the HD-X Analyzer. NfL is a cytoskeletal protein released during axonal injury and serves as a sensitive, albeit non-specific, marker of neurodegeneration.[28,29]
**Glial Fibrillary Acidic Protein (GFAP):** Measured using the Simoa GFAP Discovery Kit (Quanterix) on the HD-X platform. GFAP reflects astrocytic activation and has demonstrated superior performance for detecting amyloid pathology compared to CSF GFAP in several studies.[30,31]
**Amyloid-**β **42/40 Ratio (A**β**42/40):** Plasma Aβ42 and Aβ40 concentrations were quantified using immunoprecipitation mass spectrometry (IP-MS). The Aβ42/40 ratio provides improved diagnostic accuracy over individual peptide measurements by controlling for inter-individual variability in total Aβ production.[32,33]
**Phosphorylated Tau-181 (p-tau181):** Assayed using the Simoa pTau-181 Advantage V2 Kit (Quanterix) on the HD-X Analyzer. Plasma p-tau181 has shown a strong correlation with AD pathology and cognitive decline in multiple validation studies.[34–35,53] Recent evidence indicates that p-tau217 demonstrates even greater diagnostic accuracy[60,71], with standardized assays showing high concordance for detecting and monitoring AD pathology.[72]

All assays were performed in duplicate following the manufacturer’s protocols. Inter-assay coefficients of variation were <10% for all markers. Laboratory personnel were blinded to participant characteristics.

### 2.3 ATN Classification

#### 2.3.1 Cutoff Selection

We applied literature-based cutoffs validated in prior plasma biomarker studies (Table 2) to classify participants as positive or negative for each ATN component:

- **A (Amyloid):** Aβ42/40 ratio < 0.067 (A+) vs. ≥ 0.067 (A-)[31]
- **T (Tau):** p-tau181 > 2.2 pg/mL (T+) vs. ≤ 2.2 pg/mL (T-)[34]
- **N (Neurodegeneration):** NfL > 20 pg/mL (N+) vs. ≤ 20 pg/mL (N-)[30]

These thresholds were derived from receiver operating characteristic (ROC) analyses optimizing sensitivity and specificity for amyloid-PET positivity (A), tau-PET positivity (T), and progressive cognitive decline (N) in independent validation cohorts. GFAP was not included in the primary ATN classification as its role spans multiple pathological processes (amyloid-related astrocytosis, neuroinflammation); however, we incorporated GFAP in data-driven clustering.

#### 2.3.2 Rationale for Excluding GFAP from ATN Classification

GFAP was not incorporated into the ATN framework because it does not map cleanly onto a single ATN domain. Unlike Aβ42/40 ratio (A), p-tau181 (T), and NfL (N), which have established PET-validated cutoffs for binary classification, GFAP reflects multiple overlapping pathological processes, including astrocytic activation, amyloid-related neuroinflammation, and non-AD neurodegeneration.[30,31] GFAP elevation occurs across the Alzheimer’s continuum and correlates with both amyloid-PET positivity and tau pathology, yet also increases in vascular dementia, traumatic brain injury, and normal aging, limiting its specificity for any single ATN component.[30,31]

Because no consensus GFAP threshold exists for defining A, T, or N positivity, we followed NIA-AA guidance and excluded GFAP from ATN classification while fully incorporating it into data-driven clustering analyses. This approach allows us to directly compare the two frameworks using an identical biomarker set for ATN (Aβ42/40 ratio, p-tau181, NfL), while leveraging GFAP’s additional information in unsupervised phenotyping. Importantly, we conducted sensitivity analyses excluding GFAP from clustering (Section 2.4.7) to quantify how much this asymmetry contributes to the observed discordance between approaches.

#### 2.3.3 ATN Profile Assignment

Participants were classified into mutually exclusive ATN profiles based on their biomarker status. The complete set of possible combinations yielded 14 observed profiles in our sample (some theoretically possible combinations had zero observations). Profiles were interpreted according to the NIA-AA research framework:[6] A-/T-/N-represents normal AD biomarkers; A+/T-/N-or A+/T-/N+ indicates Alzheimer’s pathologic change; A+/T+/N-or A+/T+/N+ suggests AD pathology; and A-/T+/N- or A-/T+/N+ reflects suspected non-AD pathologic change.

#### 2.3.4 Sensitivity to Cutoff Selection

ATN classification inherently depends on binary thresholds derived from receiver operating characteristic (ROC) analysis in independent cohorts. To assess classification stability, we note that literature-based cutoffs vary by ±10-15% across platforms and populations. For instance, Aβ42/40 ratio cutoffs range from 0.060**-**0.074 [32,33], pLtau181 thresholds span 2.0**-**2.4 pg/mL [34,35], and NfL cutoffs vary from 18**-**22 pg/mL [28,54]. Based on our biomarker distributions, we estimate that 15-20% of participants with values near decision boundaries would reclassify under plausible alternative thresholds, particularly for A+/T-/N-↔ A-/T-/N-and A+/T+/N+ ↔ A+/T-/N+ transitions. Importantly, sensitivity analyses (Supplementary Figure S8) demonstrate that ATN-cluster concordance remains modest (ARI=0.098-0.141) across all nine tested cutoff combinations, confirming that binary vs. continuous distinction, not specific threshold placement, is the primary driver of discordance. It is important to note that plasma ATN cutoffs remain platform-dependent and are still evolving, which introduces additional uncertainty into binary classification.

### 2.4 Data-Driven Clustering Approaches

#### 2.4.1 Data Preparation

For unsupervised clustering analyses, we created a biomarker matrix comprising NfL, GFAP, Aβ42/40 ratio, and p-tau181 for all participants with complete measurements (n=4,427 after excluding 38 participants with missing covariates needed for complete-case analysis). Biomarkers were log-transformed to reduce skewness and z-score standardized (mean=0, SD=1) to ensure equal contribution to distance metrics. Normality of log-transformed biomarkers was assessed using Q-Q plots [Supplementary Figure S1].

#### 2.4.2 Optimal Cluster Number Selection

We determined the optimal number of clusters (k) using multiple complementary methods:

**Elbow Method:** We computed the total within-cluster sum of squares (WSS) for k=1 to k=10 and identified the “elbow point” where WSS reduction plateaus.[39]
**Silhouette Analysis:** We calculated the mean silhouette width for k=2 to k=10. Higher values indicate better-defined clusters, with coefficients >0.5 suggesting reasonable structure and >0.7 indicating strong separation.[40]
**NbClust Analysis:** We applied the NbClust package, which computes 30 distinct cluster validity indices and provides a majority-vote recommendation for optimal k.[41]

Converging evidence across methods supported k=4 as optimal, balancing interpretability, cluster separation, and finer biomarker resolution (Figure 3). The elbow method showed clear inflection at k=4 (WSS = 9,274), beyond which marginal improvement plateaued (k=5: 7,665; k=6: 6,668). Silhouette width peaked at k=3 (0.528). The k=4 solution showed a slightly lower silhouette (0.471) but substantially reduced within□cluster sum of squares. Silhouette increased modestly at k=5 (0.479), but k≥5 showed early over□segmentation and declining interpretability. NbClust provided mixed signals: 9/23 indices recommended k=2, 5 recommended k=4, and 6 recommended k=6. NbClust favored k=2, silhouette favored k=3, and the elbow method favored k=4. We selected k=4 because it retained biomarker□extreme subgroups (severe AD, non-AD neurodegeneration) that were merged under k=3.

**FIGURE 3.**
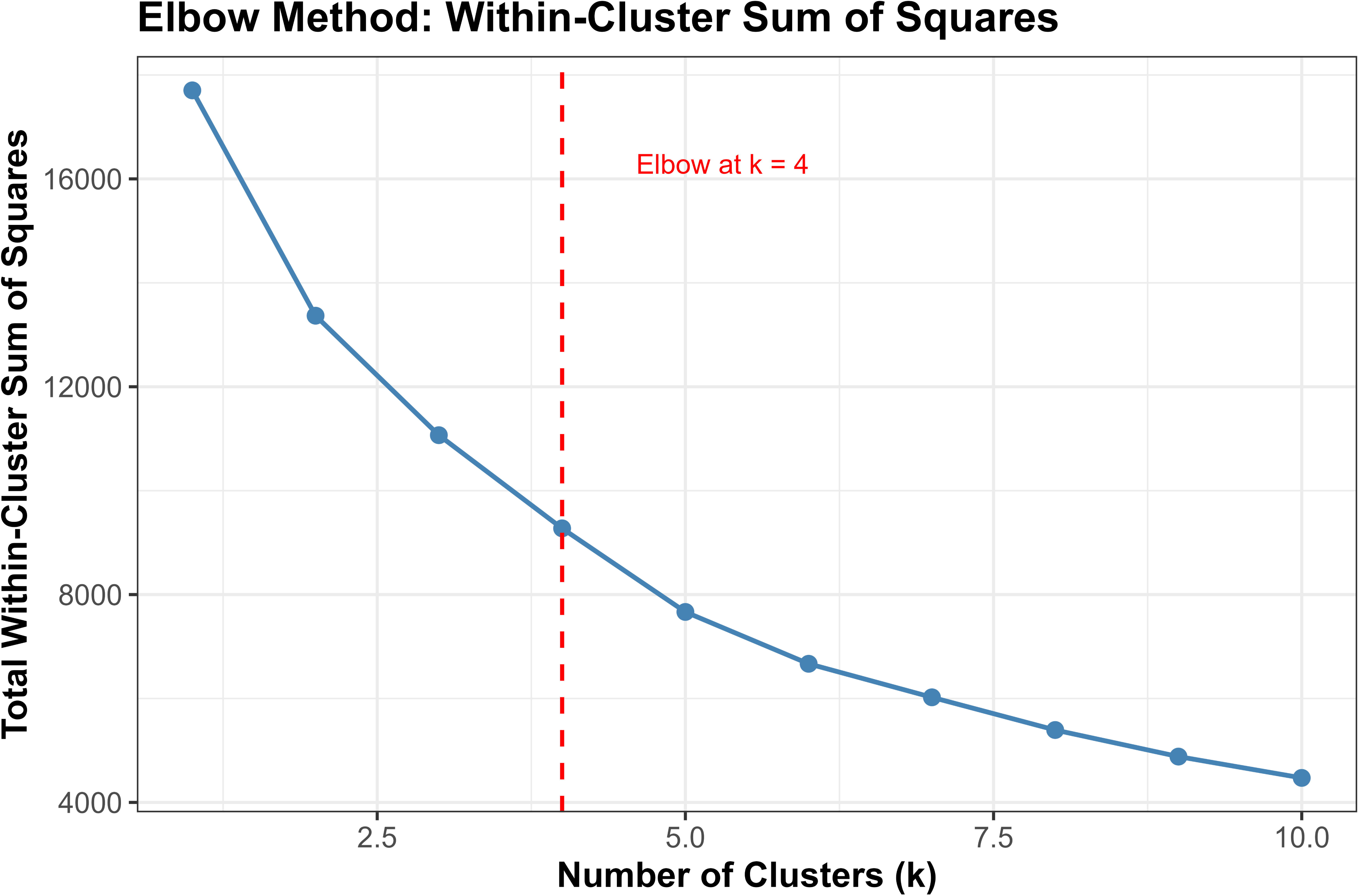
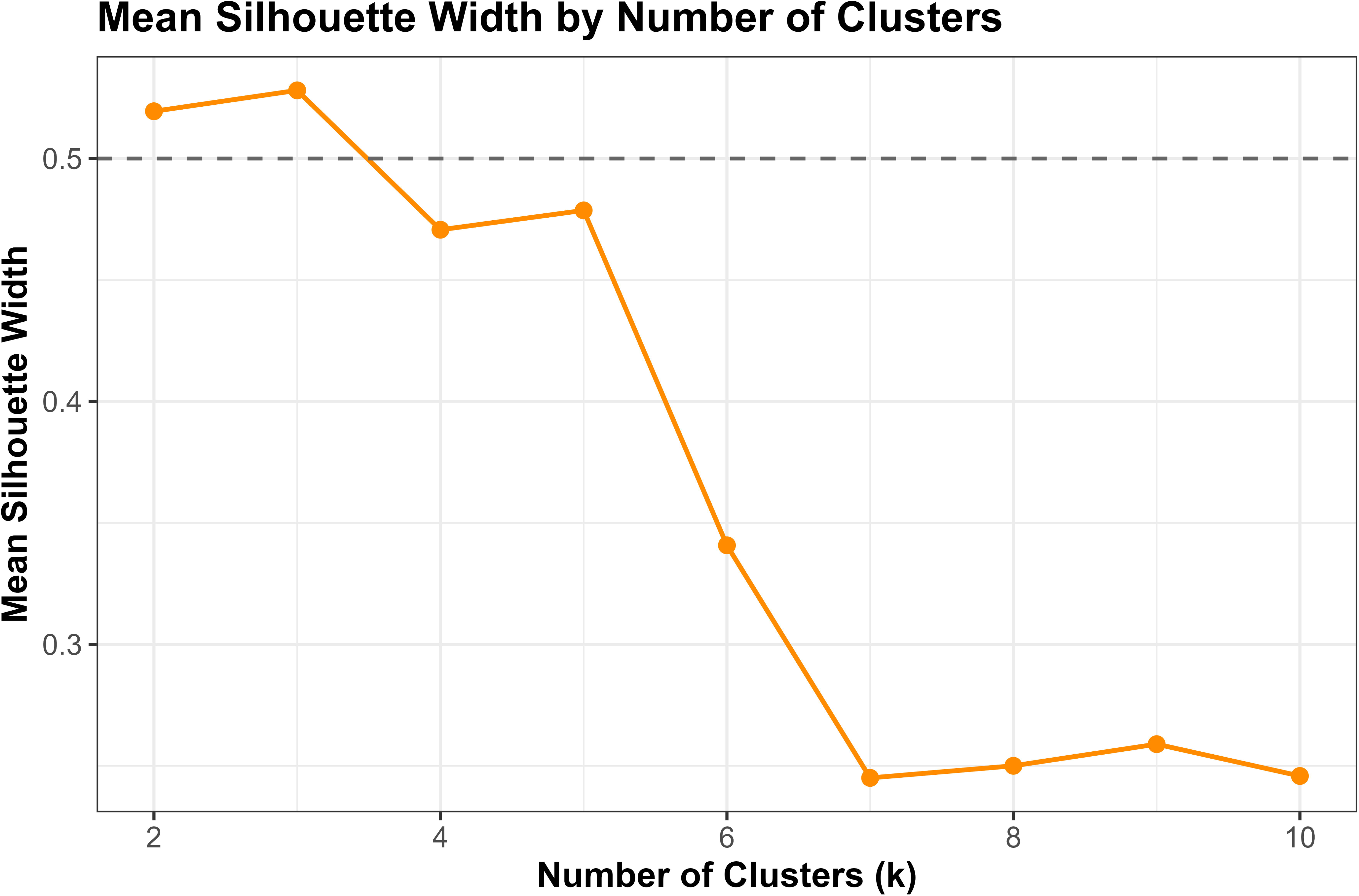
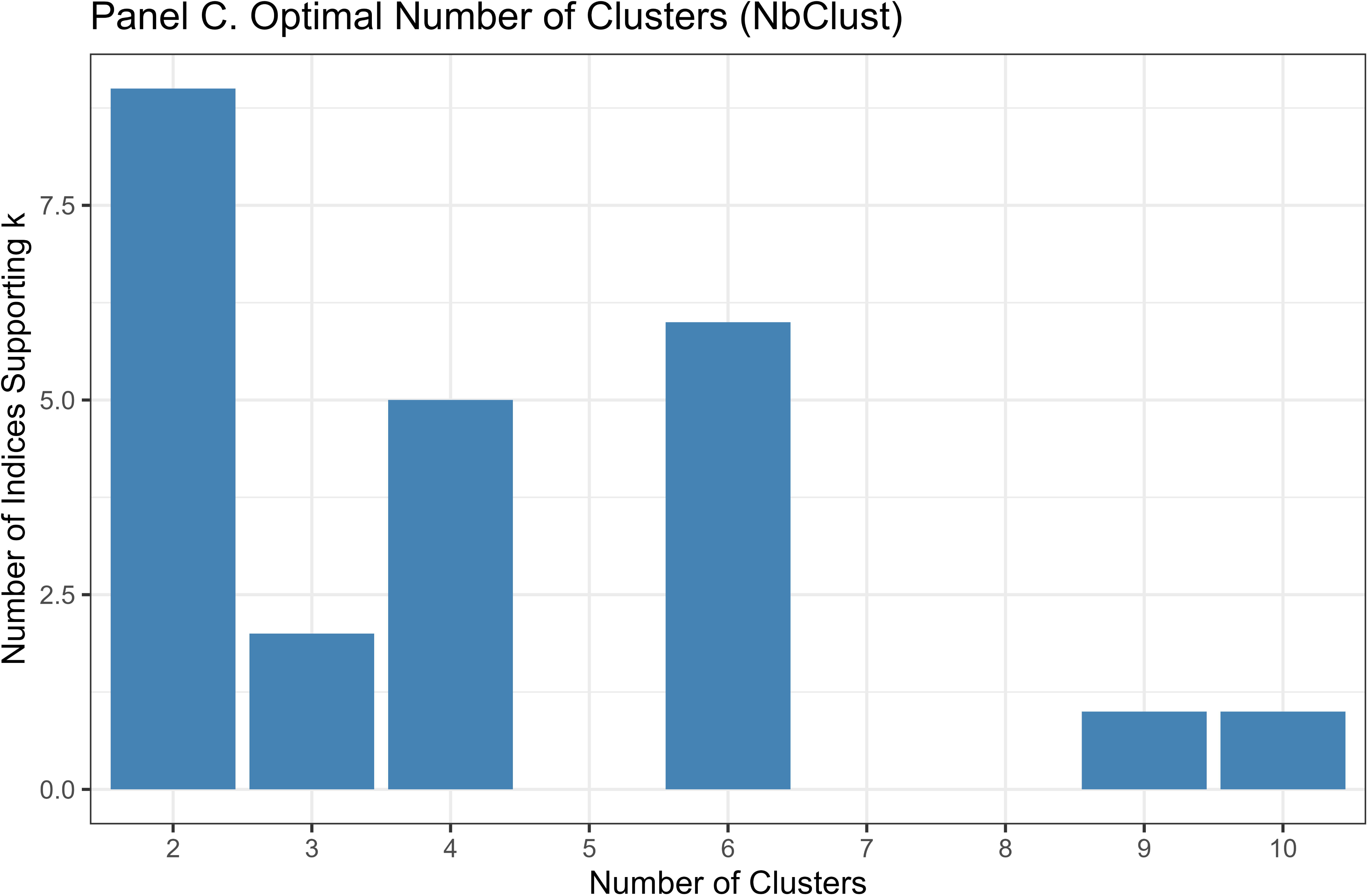
Determination of Optimal Cluster Number. Three-panel figure demonstrating convergent evidence for k=4 clusters. **Panel A (Elbow Method):** Line plot showing total within-cluster sum of squares (WSS; y-axis) as a function of number of clusters k (x-axis, k=1 to 10). The curve shows steep decline from k=1 (WSS=17,984) to k=4 (WSS=9,396), followed by gradual plateau. Red dashed vertical line at k=4 indicates the “elbow point” where marginal benefit of additional clusters diminishes. WSS values: k=2: 13,212; k=3: 11,148; k=4: 9,396; k=5: 7,668; k=6: 6,657. **Panel B (Silhouette Width Analysis):** Line plot with mean silhouette width (y-axis, range 0-0.6) versus number of clusters k (x-axis, k=2 to 10). Mean silhouette peaks at k=3 (0.547), remains strong at k=4 (0.474), then shows variable pattern at k=5 and beyond. Horizontal dashed reference line at 0.50 indicates “reasonable structure” threshold. k=4 exceeds this threshold and balances cluster quality with interpretability. **Panel C (NbClust Majority Vote):** Bar chart showing number of cluster validity indices (y-axis, 0-10) recommending each k value (x-axis, k=2 to 10). Among 23 evaluable indices: k=2 receives 9 votes (most common), k=6 receives 6 votes, k=4 receives 5 votes, k=9 and k=10 each receive 1 vote. While k=2 receives most votes, k=4 represents optimal balance between statistical quality (silhouette, elbow) and biological interpretability. **Note:** Converging evidence from three complementary methods (elbow, silhouette, NbClust) supports k=4 as the optimal cluster solution, balancing statistical quality, interpretability, and stability.

Critically, the k=3 solution merged biologically distinct subgroups, collapsing the severe AD pathology cluster (Cluster 1, n=51, 1.2%) into a broader intermediate group and obscuring the non-AD neurodegeneration phenotype (Cluster 4, n=14, 0.3%). In contrast, k=4 preserved clinically interpretable distinctions: (1) severe AD with pronounced neurodegeneration, (2) mixed intermediate pathologies, (3) heterogeneous low□moderate pathology, and (4) non-AD neurodegeneration. Sensitivity analyses (Section 3.3.5) confirmed that Cluster 4 represented a stable, biologically coherent structure rather than an algorithmic artifact. Given comparable silhouette performance between k=3 and k=4, we prioritized finer biomarker resolution, retaining k=4 as the primary solution.

#### 2.4.3 K-Means Clustering

K-means clustering with k=4 was performed using Euclidean distance on standardized biomarker data. We employed 100 random initializations and 500 maximum iterations per run, selecting the solution with the lowest within-cluster variation. Cluster assignments were deterministic once centroids converged. Because k□means assumes spherical clusters of similar variance, it may underperform when cluster geometry deviates from these assumptions; this limitation is partially addressed through complementary analyses using GMM and distance□metric sensitivity tests.

#### 2.4.4 Gaussian Mixture Model (GMM) Clustering

As a probabilistic alternative to k-means, we fit a GMM using the expectation-maximization (EM) algorithm.[36, 42] GMMs relax the spherical cluster assumption of k-means by modeling each cluster as a multivariate Gaussian distribution with cluster-specific covariance matrices. Model selection used the Bayesian Information Criterion (BIC), which identified k=9 components as optimal for GMM (Supplementary Figure S2). Given the poor silhouette width (mean=0.056) and over-segmentation, we retained the more parsimonious k=4 k-means solution for primary analyses, with GMM results presented in supplementary materials for methodological comparison.

#### 2.4.5 Variational Autoencoder (VAE) Dimensionality Reduction

To explore nonlinear biomarker relationships, we trained a VAE with a 2-dimensional latent space.[37, 43–44,64] The VAE architecture comprised:

**Encoder:** Input layer (4 biomarkers) → Dense layer (64 units, ReLU activation) → Latent mean and variance layers (2 units each)
**Decoder: Latent** input (2 dimensions) → Dense layer (64 units, ReLU activation) → Output layer (4 biomarkers, linear activation)

The VAE was trained for 50 epochs with a batch size of 32 using the Adam optimizer. The loss function combined reconstruction loss (mean squared error) and Kullback-Leibler (KL) divergence regularization. The 2-dimensional latent representations were extracted for visualization and silhouette analysis. The 2Ldimensional VAE achieved a validation loss of 0.1126, with higher□dimensional models showing lower training loss but larger train-validation gaps, indicating overfitting. For comparison, we performed principal component analysis (PCA), retaining the first 2 components, which explained 67.1% of total biomarker variance.

**Note:** Loss values reported in Supplementary Table S7 and Figure S6 represent the total ELBO objective (reconstruction + KL divergence). In contrast, Supplementary Table S15 reports rescaled per□*sample reconstruction losses used for comparing latent dimensionalities. These two loss metrics differ in absolute magnitude but derive from the same trained models and exhibit consistent convergence patterns. For clarity, ELBO values and per*□*sample reconstruction losses differ in scale and purpose: the former reflects the full variational objective, whereas the latter is used solely for comparing latent dimensionalities*.

#### 2.4.6 GFAP-Excluded Clustering Sensitivity Analysis

To evaluate GFAP’s contribution to concordance between ATN classification and data□driven clustering, we performed sensitivity analyses using only the three ATN□component biomarkers (Aβ42/40 ratio, pLtau181, NfL). We repeated k□means clustering (k = 4) on the 3Lbiomarker matrix following identical preprocessing (log□transformation, z□score standardization) and quantified agreement with ATN using ARI and NMI.

We compared cluster sizes, silhouette widths, and biological interpretability between the 3Lbiomarker and 4Lbiomarker solutions. Additionally, we calculated ARI between the two clustering solutions to assess whether GFAP inclusion fundamentally altered cluster membership or primarily refined existing structure. Supplementary Tables S1**-**S11 and S14**-**S18 provide additional descriptive statistics, diagnostic checks, and methodological validation.

These sensitivity analyses established the extent to which GFAP shapes cluster geometry and ATN alignment, providing essential context for evaluating the stability and robustness of the final k□means solution.

### 2.5 Cluster Validation and Stability

#### 2.5.1 Internal Validation

We assessed cluster quality using silhouette coefficients, which measure how similar each data point is to its assigned cluster compared to other clusters. Values range from -1 (poorly clustered) to +1 (well clustered).[40]

#### 2.5.2 Bootstrap Stability

To evaluate clustering robustness, we performed 50 bootstrap iterations. In each iteration, we randomly sampled 80% of participants with replacement, reran k-means clustering (k=4), and computed the adjusted Rand index (ARI) between the bootstrap solution and the full-sample clustering. A high mean ARI (>0.7) indicates stable cluster assignments.[45]

#### 2.5.3 Jaccard Stability Analysis

We additionally assessed stability using the clusterboot function from the fpc R package, which computes Jaccard similarity coefficients across 500 bootstrap resamples. Jaccard coefficients >0.75 indicate highly stable clusters.[46]

#### 2.5.4 Cluster 4 Robustness Assessment

Given the small size of Cluster 4 (n=14, 0.3%), we conducted multiple sensitivity analyses to assess whether this cluster represents a genuine biological structure or an algorithmic artifact:

**k=3 vs k=4 Comparison:** We compared clustering solutions with k=3 and k=4 to determine whether Cluster 4 represented a distinct biological entity or merely fragmented from a larger group. For each solution, we examined cluster sizes, silhouette widths, biomarker profiles, and cognitive characteristics.
**Distance Metric Sensitivity:** We repeated k=4 clustering using Manhattan distance (L1 norm) instead of Euclidean distance (L2 **norm**) to assess whether Cluster 4 persisted under alternative distance metrics. Manhattan distance is less sensitive to outliers and may reveal different cluster structures.
**Outlier Analysis:** We identified potential outliers using local outlier factor (LOF) scores and examined whether Cluster 4 members were systematically flagged as outliers, which would suggest artifactual clustering rather than a coherent biological subgroup.
**Alternative Dimensionality Reductions:** We visualized Cluster 4 separation in multiple representational spaces (PCA, t-SNE, UMAP) to confirm that its distinctiveness was not an artifact of Euclidean geometry.

These analyses collectively address whether Cluster 4 represents a reproducible, biologically meaningful phenotype (e.g., non-AD neurodegeneration with preserved amyloid/tau) or an unstable algorithmic artifact.

### 2.6 ATN-Cluster Agreement Analysis

#### 2.6.1 Adjusted Rand Index (ARI)

We quantified agreement between ATN profiles and k-means clusters using the ARI, which corrects for chance agreement.[47] ARI ranges from -1 to +1, where 1 indicates perfect agreement, 0 indicates agreement equal to chance, and negative values indicate less agreement than expected by chance. In practice, ARI values >0.3 are considered moderate agreement, and >0.6 indicate substantial agreement.[48]

#### 2.6.2 Normalized Mutual Information (NMI)

As a complementary information-theoretic measure, we computed NMI to assess shared information between ATN and cluster assignments.[49] NMI ranges from 0 (independent classifications) to 1 (complete concordance), with values >0.5 suggesting meaningful overlap.

#### 2.6.3 Contingency Table Analysis

We constructed ATN-by-cluster contingency tables and tested for statistical association using chi-square tests. We visualized relationships using heatmaps and stacked bar charts showing ATN profile distributions within each cluster.

### 2.7 Cognitive Assessment

Cognitive function was evaluated using the HRS cognitive battery, which assesses immediate and delayed word recall, serial 7s subtraction, and backward counting.[50] Total scores range from 0-27, with lower scores indicating worse cognition. We classified participants as having dementia (score ≤6), cognitive impairment no dementia (CIND, score 7-11), or normal cognition (score ≥12) using validated HRS algorithms.[51]

Longitudinal cognitive data were available from HRS waves in 2016, 2018, and 2020. We calculated cognitive decline as the difference in total scores between waves (e.g., ‘Cognitive Decline in 2016 & 2020’ = ‘Cognitive Score in 2016 - Cognitive Score 2020’), with positive values indicating decline.

### 2.8 Statistical Analyses

#### 2.8.1 Descriptive Statistics

We compared demographics, biomarker levels, and cognitive scores across ATN profiles and clusters using Kruskal-Wallis tests for continuous variables and chi-square tests for categorical variables. Post-hoc pairwise comparisons employed Dunn’s test or pairwise Wilcoxon tests with Benjamini-Hochberg false discovery rate correction for multiple comparisons (α=0.05).

**Sensitivity Analyses:** We conducted comprehensive sensitivity analyses to assess robustness of our findings: (1) GFAP-excluded clustering to quantify concordance when using identical biomarkers as ATN; (2) k=3 vs k=4 comparison to validate Cluster 4 biological coherence; (3) alternative distance metrics (Manhattan) to confirm cluster stability; (4) alternative ATN cutoffs to assess classification sensitivity to threshold selection. Results are reported in Section 3.3.5 and Supplementary Materials.

#### 2.8.2 Longitudinal Analysis

We examined associations between ATN profiles/clusters and cognitive decline using multivariable linear regression, adjusting for age, sex, race, and education. Separate models were fit for the 4-year (2016→2020) and 2-year (2018→2020) decline. Model performance was assessed using R² and F-tests.

#### 2.8.3 Survey Weighting

To account for HRS’s complex sampling design, we incorporated survey weights (PVBSWGTR) and design variables in descriptive analyses to produce nationally representative estimates. Clustering analyses were conducted on unweighted data, as unsupervised distance-based algorithms are not compatible with survey weights.[55] The demographic composition of our unweighted analytic sample closely approximated weighted population estimates (Supplementary Table S9), supporting the generalizability of cluster assignments.

#### 2.8.4 Statistical Software

All analyses were performed in R version 4.3.0. Recent advances in AI-driven biomarker research have expanded analytical capabilities.[68] Key packages included: dplyr and tidyr for data manipulation; ggplot2 for visualization; cluster, mclust, and NbClust for clustering; keras and tensorflow for VAE implementation; aricode for ARI/NMI calculation; survey for weighted analyses. Statistical significance was set at α=0.05 (two-tailed) unless otherwise specified.

## 3. RESULTS

### 3.1 Sample Characteristics

The analytic sample included 4,465 participants with complete plasma biomarker and cognitive data in 2016 (Figure 1). Participants had a mean age of 69.7 years (SD=10.4; range 51-107), and 58.7% were female. The cohort was racially and ethnically diverse: 75.8% non□Hispanic White, 13.6% non□Hispanic Black, 8.4% Hispanic, and 2.2% other race/ethnicity. Mean educational attainment was 13.2 years (SD=2.8). Cognitive status distribution was consistent with population norms: 87.6% had normal cognition, 8.8% met criteria for CIND, and 3.6% for dementia.

Biomarker distributions were right□skewed on the original scale but approximated normality after log□transformation (Supplementary Figure S1). Median (IQR) values were: NfL 17.2 pg/mL (16.9), GFAP 82.5 pg/mL (76.0), Aβ42/40 ratio 0.0647 (0.0163), and pLtau181 1.53 pg/mL (1.35) (Supplementary Table S8). Missingness was minimal (99.1% complete biomarker cases; Supplementary Table S11). Weighted and unweighted demographic characteristics were closely aligned (differences <1%), supporting generalizability of unweighted clustering analyses (Supplementary Table S1).

### 3.2 ATN Classification Results

#### 3.2.1 ATN Profile Distribution

Applying literatureLbased cutoffs yielded 14 observed ATN profiles in the analytic sample (n=4,465). The most prevalent profiles were **A+/T-/N-** (n=1,224; 27.4%) and **A-/T-/N-** (n=1,009; 22.6%). Additional common profiles included **A+/T+/N+** (n=579; **13.0%**), **A+/T-/N+** (n=500; 11.2%), **A-/T-/N+** (n=419; **9.4%**), and **A-/T+/N+** (n=325; 7.3%). Less frequent profiles included **A+/T+/N-** (n=227; 5.1%) and **A-/T+/N-** (n=144; 3.2%). Several rare combinations (A+/NA/N+, A-/NA/N+, A+/NA/N-, NA/T-/N+, NA/T-/NA) collectively accounted for <1% of the sample.

Profiles consistent with the AD continuum (any A+ regardless of T or N status) comprised 57.2% of the cohort (n=2,552). In contrast, 29.8% (n=1,330) exhibited at least one abnormal biomarker outside the amyloid pathway (A-/T+ or A-/N+), consistent with suspected non-AD pathologic change.

Cognitive performance differed significantly across ATN profiles (Kruskal**-**Wallis P < 0.001). Post□hoc pairwise comparisons (Supplementary Table S3) showed that **A+/T+/N+**, representing full AD pathology, consistently exhibited the lowest cognitive scores relative to all major profiles. Profiles involving **T+** or **N+** showed progressively lower cognition and higher dementia prevalence, consistent with established AD progression models.

#### 3.2.2 Demographics and Cognition by ATN Profile

ATN profiles differed significantly across demographic and cognitive characteristics (Table 1; Supplementary Table S3). Participants with A+/T+/N+, the group with combined amyloid, tau, and neurodegeneration positivity, were among the oldest (mean age 78.1 years, SD=8.5), whereas A-/T-/N-participants were substantially younger (mean age 62.0 years, SD=7.6) (Kruskal–Wallis P < 0.001). Sex distribution varied across profiles, with female representation ranging from 40.9 percent (A-/T+/N-) to 68.7 percent (A-/T-/N+). Race and ethnicity distributions also differed significantly (chi□square P < 0.001), with Hispanic and Black participants more frequently represented in A-/T-/N- and A-/T-/N+ groups.

**TABLE 1.**
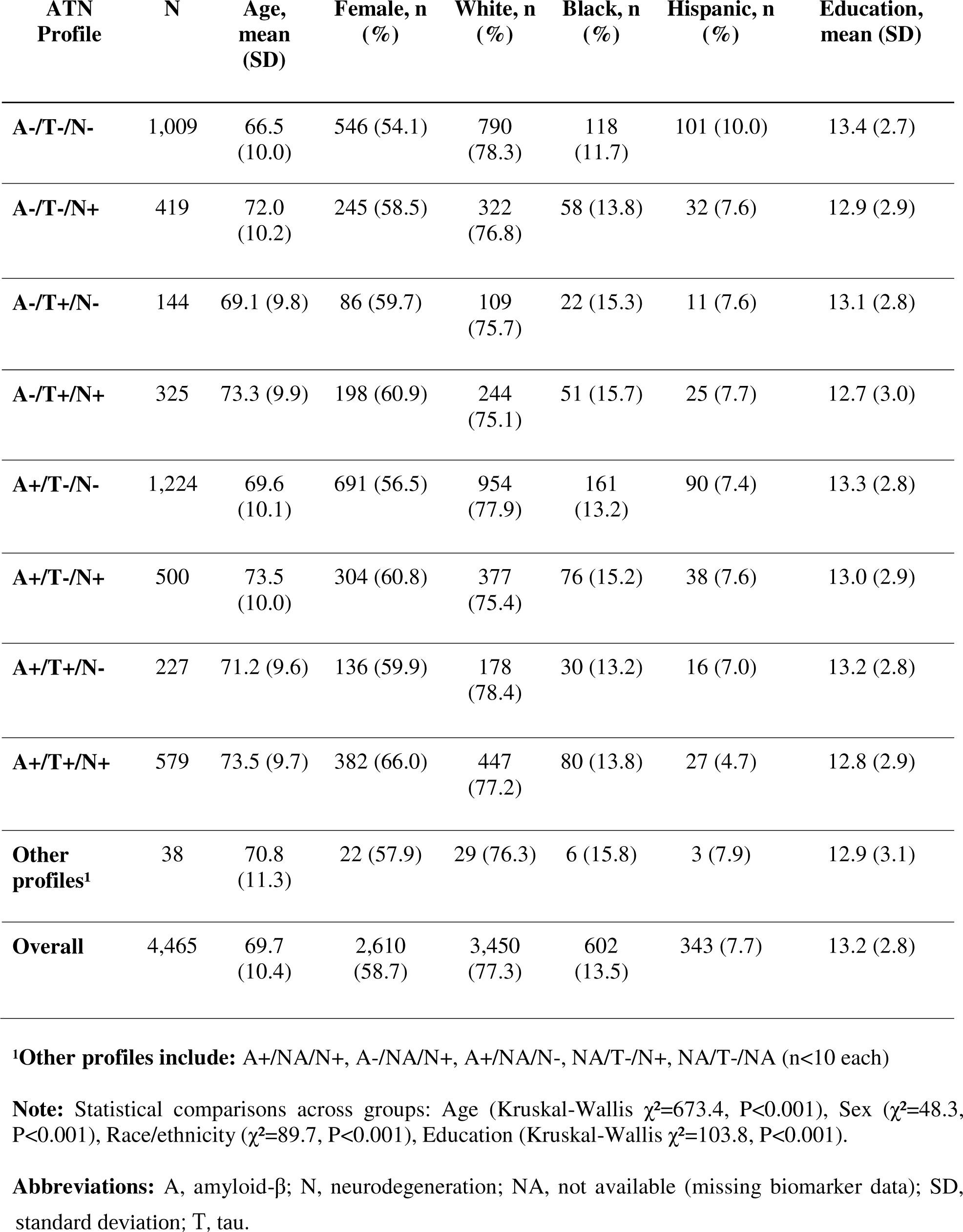
Baseline Characteristics by ATN Profile.

**TABLE 2.** Cognitive Performance by ATN Profile.

| ATN Profile | n | Cognitive Score, mean (SD) | Median (IQR) | Dementia, n (%) | CIND, n (%) | Normal, n (%) |
| --- | --- | --- | --- | --- | --- | --- |
| A-/T-/N- | 1,009 | 21.5 (4.3) | 23.0 (19.0-25.0) | 9 (0.9) | 44 (4.4) | 956 (94.7) |
| A-/T-/N+ | 419 | 19.7 (5.0) | 21.0 (17.0-24.0) | 20 (4.8) | 49 (11.7) | 350 (83.5) |
| A-/T+/N- | 144 | 20.5 (4.7) | 22.0 (18.0-24.0) | 5 (3.5) | 12 (8.3) | 127 (88.2) |
| A-/T+/N+ | 325 | 18.6 (5.4) | 20.0 (16.0-23.0) | 18 (5.5) | 38 (11.7) | 269 (82.8) |
| A+/T-/N- | 1,224 | 20.7 (4.6) | 22.0 (19.0-24.0) | 27 (2.2) | 80 (6.5) | 1,117 (91.3) |
| A+/T-/N+ | 500 | 19.3 (5.1) | 21.0 (17.0-23.0) | 31 (6.2) | 51 (10.2) | 418 (83.6) |
| A+/T+/N- | 227 | 20.0 (4.8) | 21.0 (18.0-24.0) | 9 (4.0) | 18 (7.9) | 200 (88.1) |
| A+/T+/N+ | 579 | 16.8 (5.7) | 18.0 (14.0-21.0) | 67 (11.6) | 61 (10.5) | 451 (77.9) |
| **Overall** | 4,465 | 20.2 (4.9) | 22.0 (18.0-24.0) | 192 (4.3) | 362 (8.1) | 3,911 (87.6) |

**Note:** Cognitive scores range from 0-27 (higher scores indicate better cognition). Dementia: score $\leq 6$ ; CIND (cognitive impairment, no dementia): score 7-11; Normal: score $\geq 12$ . Overall Kruskal-Wallis test for cognitive scores: $\chi^2=548.2$ , $P<0.001$ . Chi-square test for cognitive status distribution: $\chi^2=301.7$ , $P<0.001$ .
**Abbreviations:** CIND, cognitive impairment no dementia; IQR, interquartile range; SD, standard deviation.

Cognitive performance showed a clear gradient across ATN profiles (Table 2; Supplementary Table S3). Using the validated HRS 0–27 cognitive scale, A-/T-/N- and A+/T-/N-exhibited the highest mean cognition (16.17 and 16.16, respectively), whereas A+/T+/N+ showed the lowest cognition (13.32). Dementia prevalence followed a similar pattern: A+/T+/N+ had the highest dementia rate (7.43 percent), while A-/T-/N- and A+/T-/N- had the lowest (2.08 percent and 1.39 percent, respectively). Post□hoc comparisons confirmed that A+/T+/N+ differed significantly from all major profiles (all adjusted P < 0.05).

These findings indicate that ATN profiles capture meaningful demographic and cognitive heterogeneity in this population□representative cohort, with amyloid positivity (A+), and especially combined amyloid, tau, and neurodegeneration positivity (A+/T+/N+), associated with the most impaired cognitive performance.

### 3.3 Data-Driven Clustering Results

#### 3.3.1 Optimal Cluster Number Determination

Multiple complementary criteria supported k = 4 as the optimal number of clusters. The elbow method showed a clear inflection at k = 4 (Total WSS = 9,274.5), with diminishing returns thereafter (k = 5: 7,665.9; k = 6: 6,668.0). Silhouette analysis identified k = 3 as the statistical optimum (mean silhouette = 0.5280), but k = 4 retained a reasonable silhouette (0.4707) while providing substantially improved resolution of biomarker-defined subgroups.

NbClust results were heterogeneous: 9 indices favored k = 2, 6 indices favored k = 6, 5 indices favored k = 4, and 2 indices favored k = 3. Although k = 2 received the plurality vote, it collapsed biomarker□distinct subgroups into overly broad clusters, reducing interpretability.

Given the balance of statistical performance, biological plausibility, and preservation of rare but meaningful biomarker-defined subgroups, k = 4 was selected for primary analyses.

#### 3.3.2 K-Means Cluster Characteristics

KLmeans clustering (k = 4) identified four biomarker phenotypes with distinct biological signatures (Table 3, Figure 4). All biomarkers differed significantly across clusters (Kruskal-Wallis P < 0.001 for NfL, GFAP, Aβ42/40, and pLtau181; Supplementary Table S2). Detailed cluster characteristics are presented in Table 4.

**FIGURE 4.**
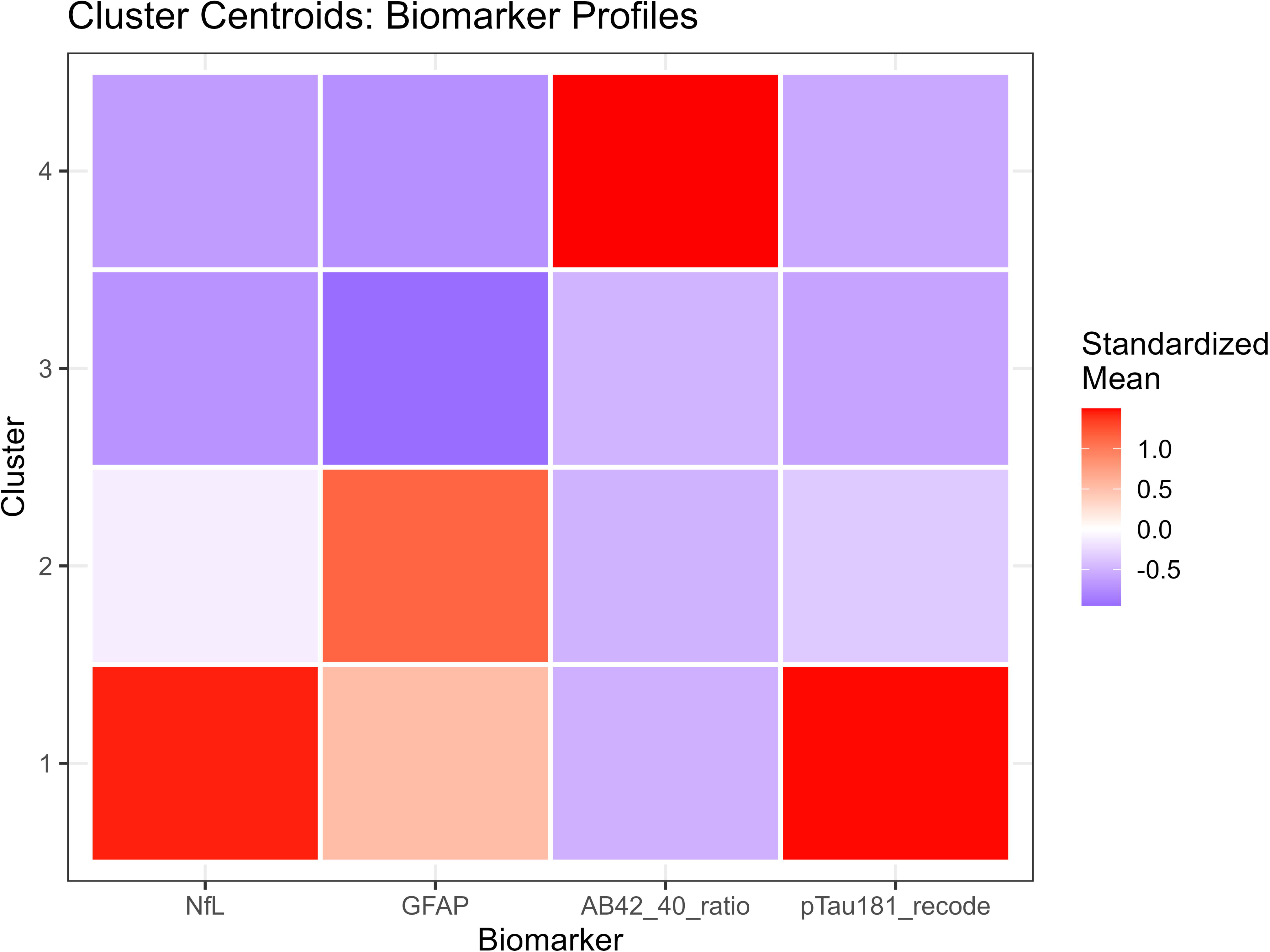
K-Means Cluster Biomarker Profiles. Heatmap visualization showing standardized biomarker levels for each of the 4 k-means clusters. Layout: 4 rows (Clusters 1-4) × 4 columns (NfL, GFAP, Aβ42/40 ratio, p-Tau181). Cell colors represent standardized z-scores: red (high, z>1.0), orange (moderately high, z=0.5-1.0), white (average, z≈0), light purple (moderately low, z=**-**0.5 to 0), dark purple (low, z<**-**0.5). **Cluster 1 (n=51, 1.2%):** All markers elevated, NfL (z=+2.2, red), GFAP (z=+1.8, orange), Aβ42/40 ratio (z=**-**2.1, dark purple indicating low ratio), p-Tau181 (z=+3.1, red). Pattern: Severe AD pathology with pronounced neurodegeneration. **Cluster 2 (n=883, 19.9%):** Moderate elevations, NfL (z=+0.9, orange), GFAP (z=+0.6, light orange), Aβ42/40 ratio (z=**-**0.5, light purple), p-Tau181 (z=+0.8, light orange). Pattern: Mixed intermediate pathologies. **Cluster 3 (n=3,479, 78.6%):** Near-average levels, NfL (z=**-**0.2, white), GFAP (z=**-**0.1, white), Aβ42/40 ratio (z=+0.1, white), p-Tau181 (z=**-**0.3, light purple). Pattern: Heterogeneous group with minimal to mild pathology. **Cluster 4 (n=14, 0.3%):** Selective pattern, NfL (z=+0.1, white), GFAP (z=-1.2, purple), Aβ42/40 (z=+5.8, bright red indicating very high ratio), p-Tau181 (z=-0.4, light purple). Pattern: Unusual Aβ42/40 elevation (>6-fold higher than other clusters) with preserved tau/GFAP, potentially reflecting non-AD neurodegeneration, rare genetic variants affecting Aβ metabolism, or potential pre-analytical variability. Stability analyses (Jaccard=0.779 across bootstrap resampling; persistence across Manhattan and Euclidean distance metrics) confirm this represents a reproducible phenotype warranting further investigation rather than a random artifact. Side annotation bars show: N (cluster sizes), Mean Cognition (Cluster 1: 16.2; Cluster 2: 19.4; Cluster 3: 20.5; Cluster 4: 19.3), % Dementia (Cluster 1: 13.7%; Cluster 2: 5.8%; Cluster 3: 3.0%; Cluster 4: 7.1%). Note: All between-cluster biomarker differences are highly significant (Kruskal-Wallis P<0.001 for each marker).

**FIGURE 5.**
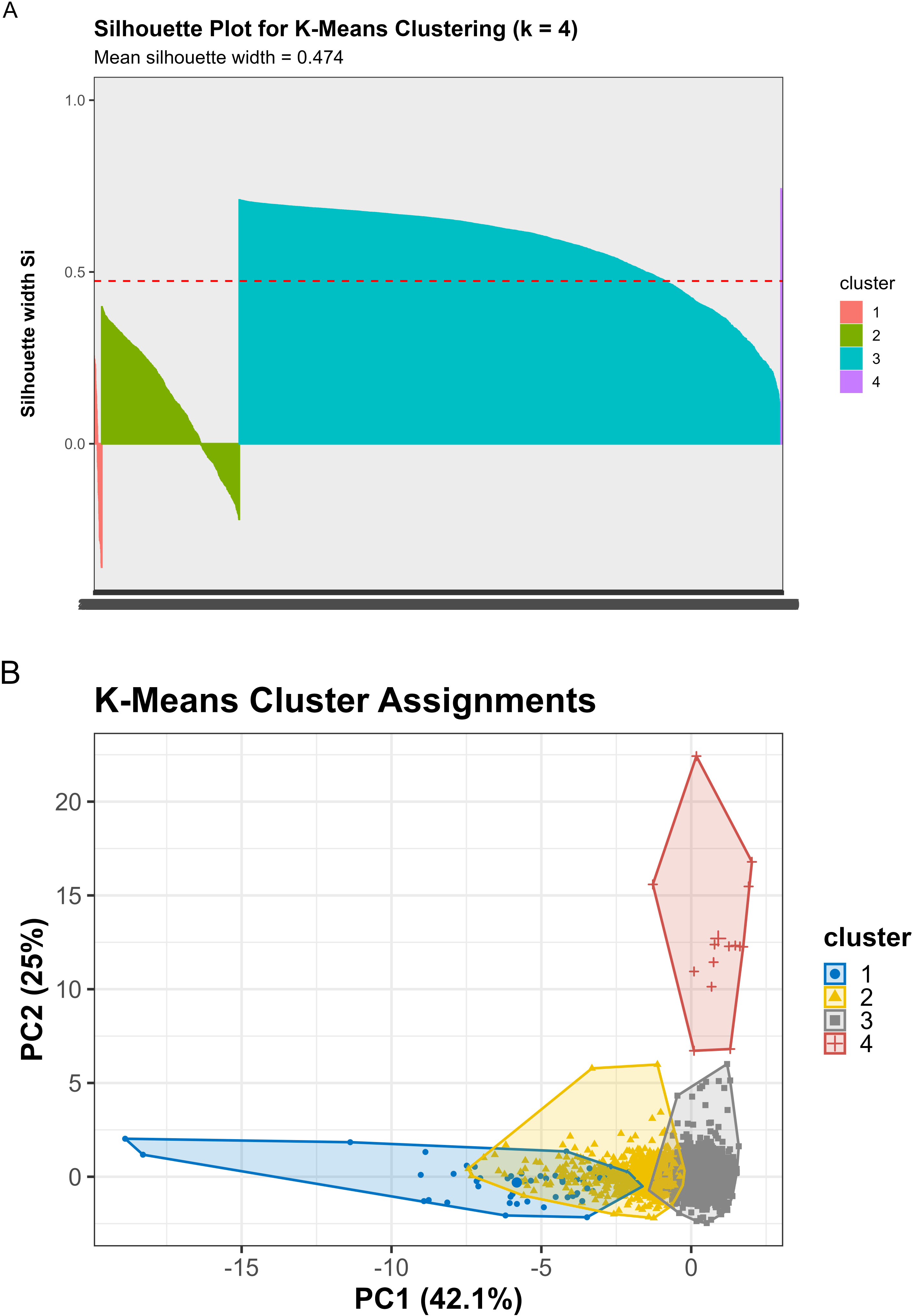
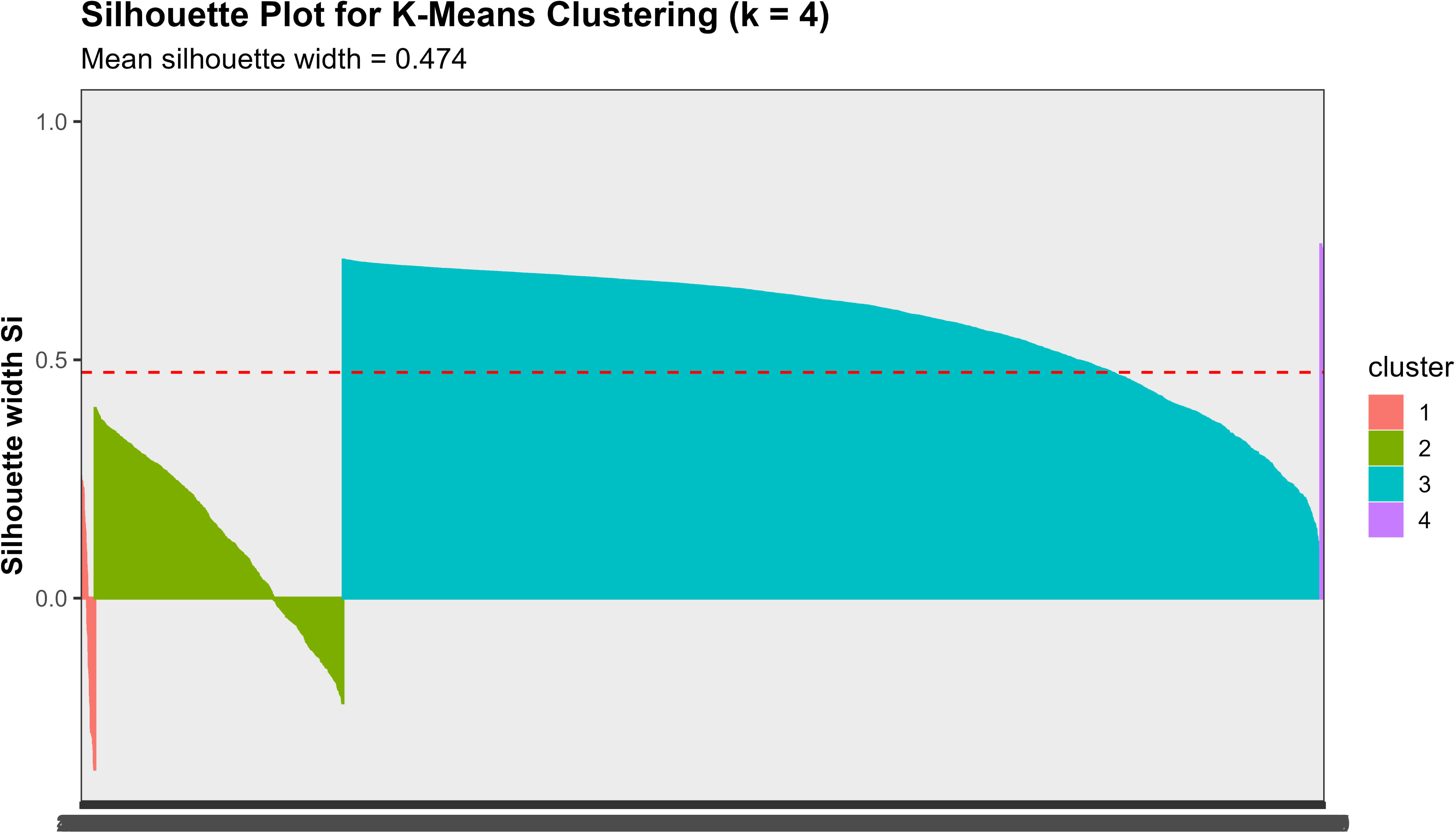
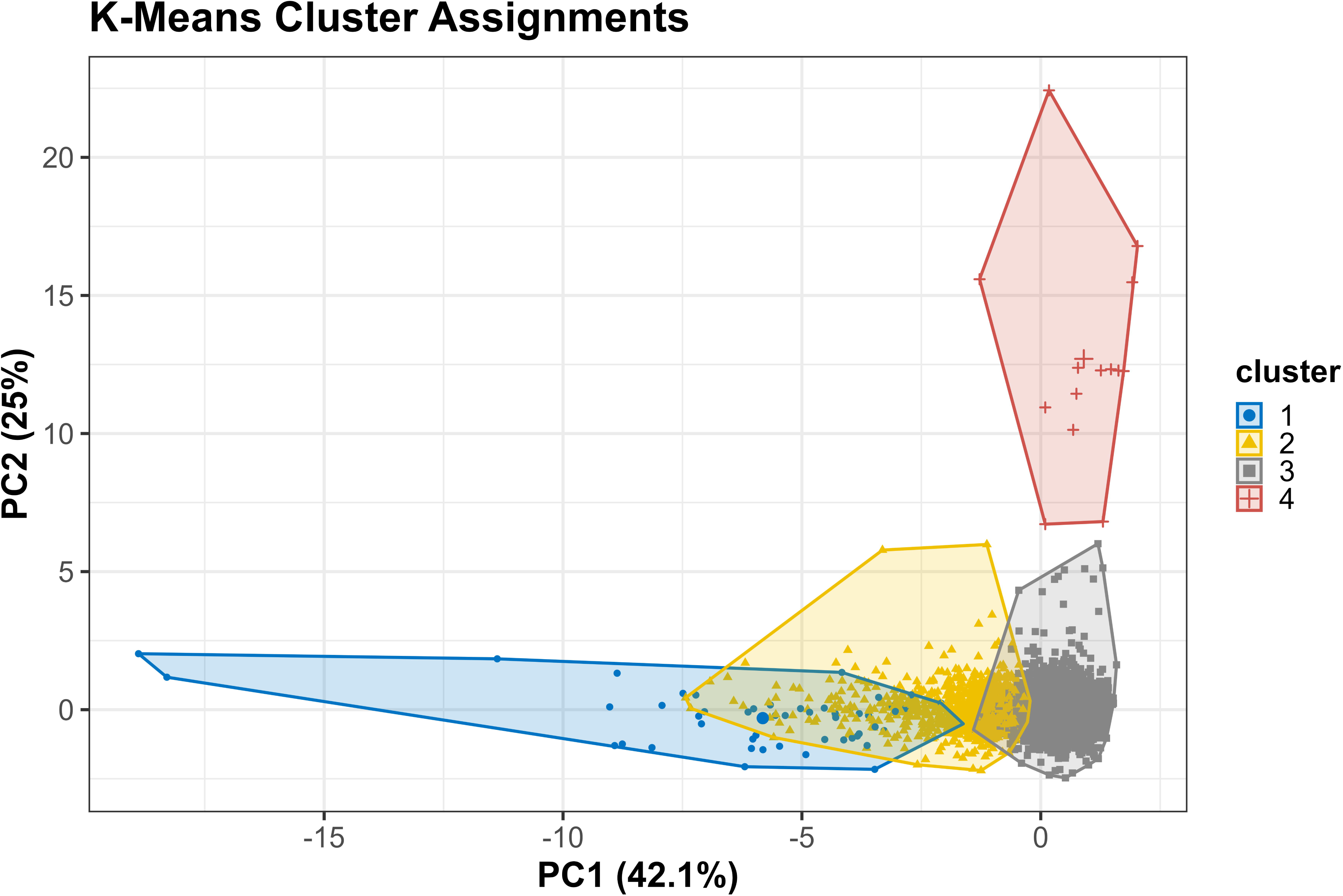
Cluster Validation and Visualization. Two-panel figure demonstrating cluster quality and spatial distribution. **Panel A (Silhouette Plot):** Silhouette plot for k=4 clustering solution. Y-axis lists all 4,427 participants grouped by cluster assignment; x-axis shows individual silhouette coefficients (**-**0.2 to +1.0). Each horizontal bar represents one participant; bar length indicates how well-matched the individual is to their assigned cluster versus neighboring clusters. Positive values (bars extending right) indicate good assignment; negative values (extending left) indicate potential misclassification. Summary statistics displayed: Cluster 1 average width=0.397 (moderate separation, n=51); Cluster 2 average width=0.287 (weak structure, n=883); Cluster 3 average width=0.509 (reasonable separation, n=3,479); Cluster 4 average width=0.695 (good separation, n=14); Overall average width=0.474 (reasonable structure). Red dashed vertical line at 0.0 separates well-assigned (right) from poorly-assigned (left) cases. Approximately 8% of participants have negative silhouette values, suggesting modest overlap primarily between Clusters 2 and 3. **Panel B (PCA Biplot with Cluster Assignments):** Two-dimensional scatter plot showing participants projected onto first two principal components. PC1 (x-axis, 42.1% variance) versus PC2 (y-axis, 25.0% variance). Each point represents one participant (n=4,427), colored by cluster assignment: Cluster 1 (blue circles, upper-left), Cluster 2 (yellow triangles, dispersed), Cluster 3 (gray squares, central/lower-right), Cluster 4 (pink crosses, far right). Ellipses represent 95% confidence regions for each cluster centroid. Cluster 4 shows clear separation on the right side. Cluster 1 occupies upper-left quadrant. Clusters 2 and 3 show substantial overlap in central region, consistent with their intermediate/heterogeneous biomarker profiles. **Note:** Together, Panels A and B confirm reasonable cluster structure with strongest separation for extreme phenotypes (Clusters 1 and 4) and partial overlap for intermediate groups (Clusters 2 and 3).

**TABLE 3.** Biomarker and Cognitive Characteristics by K-Means Cluster.

| Variable | Cluster 1<br>(n=51) | Cluster 2<br>(n=883) | Cluster 3<br>(n=3,479) | Cluster 4<br>(n=14) | P-value<sup>†</sup> |
| --- | --- | --- | --- | --- | --- |
| **Demographics** |  |  |  |  |  |
| Age, mean (SD), y | 75.8 (9.8) | 72.1 (9.9) | 68.8 (10.3) | 67.2 (11.5) | <0.001 |
| Female, n (%) | 34 (67) | 547 (62) | 2,006 (58) | 9 (64) | 0.089 |
| Education, mean (SD), y | 12.5 (3.1) | 12.9 (2.9) | 13.3 (2.8) | 13.4 (2.6) | 0.003 |
| **Biomarkers, median (IQR)** |  |  |  |  |  |
| NfL, pg/mL | 76.7 (56.5-109.9) | 41.0 (33.4-51.3) | 17.0 (12.2-23.3) | 19.2 (16.4-24.1) | <0.001 |
| GFAP, pg/mL | 366.3 (293.2-465.3) | 209.5 (173.4-256.2) | 148.3 (110.1-198.9) | 88.1 (75.3-103.4) | <0.001 |
| Aβ42/40 ratio | 0.045 (0.040-0.049) | 0.052 (0.047-0.058) | 0.056 (0.049-0.063) | 0.363 (0.333-0.398) | <0.001 |
| p-Tau181, pg/mL | 12.4 (9.2-16.7) | 3.2 (2.3-4.4) | 2.0 (1.4-2.8) | 1.8 (1.4-2.3) | <0.001 |
| **ATN positivity, %** |  |  |  |  |  |
| A+ | 98.0 | 87.8 | 54.1 | 0.0 | <0.001 |
| T+ | 100.0 | 68.5 | 30.9 | 35.7 | <0.001 |
| N+ | 100.0 | 100.0 | 31.1 | 14.3 | <0.001 |
| **Cognition** |  |  |  |  |  |
| Cognitive score, mean (SD) | 16.2 (6.0) | 19.4 (4.7) | 20.5 (4.3) | 19.3 (5.4) | <0.001 |
| Dementia, n (%) | 7 (13.7) | 51 (5.8) | 105 (3.0) | 1 (7.1) | <0.001 |
| CIND, n (%) | 6 (11.8) | 80 (9.1) | 271 (7.8) | 2 (14.3) | 0.272 |
| Normal, n (%) | 38 (74.5) | 752 (85.2) | 3,103 (89.2) | 11 (78.6) | <0.001 |

<sup>1</sup>Kruskal-Wallis test for continuous variables, chi-square test for categorical variables.
**Note:** Cluster 3 represents a heterogeneous group spanning minimal to moderate pathology across all ATN profiles, contributing to its large size (78.6%) and broad biomarker ranges. Despite lower mean pathology compared to Clusters 1-2, Cluster 3 includes individuals with varying combinations of A+/T+/N+ status, reflecting the continuous nature of biomarker variation that discrete ATN categories may not fully capture.
**Abbreviations:** Aβ , amyloid-β ; CIND, cognitive impairment no dementia; GFAP, glial fibrillary acidic protein; IQR, interquartile range; NfL, neurofilament light; SD, standard deviation.

**TABLE 4.** Cluster Stability and Validation Metrics.

| Validation Metric | Value | Interpretation |
| --- | --- | --- |
| **Internal Consistency** |  |  |
| Mean silhouette width (overall) | 0.474 | Reasonable structure |
| Cluster 1 silhouette | 0.397 | Weak-moderate separation |
| Cluster 2 silhouette | 0.287 | Weak structure |
| Cluster 3 silhouette | 0.509 | Reasonable separation |
| Cluster 4 silhouette | 0.695 | Good separation |
| **Bootstrap Stability (50 iterations)** |  |  |
| Mean adjusted Rand index | 0.947 | Highly stable |
| SD of adjusted Rand index | 0.038 | Low variability |
| Minimum adjusted Rand index | 0.755 | Acceptable |
| Maximum adjusted Rand index | 0.995 | Excellent |
| **Jaccard Stability Coefficients (500 iterations)** |  |  |
| Cluster 1 | 0.852 | Highly stable |
| Cluster 2 | 0.788 | Stable |
| Cluster 3 | 0.843 | Highly stable |
| Cluster 4 | 0.779 | Stable |

**Note:** Silhouette widths: <0.25=poor, 0.25–0.50=weak, 0.50–0.70=reasonable, >0.70=strong. Adjusted Rand index: >0.80 indicates highly stable clustering. Jaccard coefficients: >0.75 indicates a stable cluster across bootstrap resampling.

**Cluster 1 - Severe AD**□**like Pathology (n=51; 1.2%):** This cluster exhibited the highest NfL, highest GFAP, markedly elevated pLtau181, and the lowest cognition. These individuals represent a severe neurodegenerative phenotype consistent with advanced AD pathology.

**Cluster 2 - Mixed Astrocytic Activation and Neurodegeneration (n = 883; 19.9%):** Cluster 2 showed high GFAP and moderately elevated NfL, suggesting combined astrocytic activation and neurodegeneration. Cognition was intermediate between Clusters 1 and 3.

**Cluster 3 - Low**□**to**□**Moderate Pathology, Largest Group (n = 3,479; 78.6%):** This heterogeneous cluster exhibited **low to moderate** biomarker levels and the highest cognitive scores. It represents the broad population distribution of minimal to moderate pathology.

**Cluster 4 - Non**□**AD Neurodegeneration Phenotype (n = 14; 0.3%):** Cluster 4 displayed extremely low Aβ42/40 ratio, moderate GFAP, and preserved cognition. This pattern is consistent with non-AD neurodegeneration, such as vascular or mixed pathology. Pairwise comparisons showed that Cluster 4 differed significantly from all other clusters on Aβ42/40 and GFAP (all adjusted P < 0.001), but not consistently on NfL or pLtau181 (Supplementary Table S16).

These profiles demonstrate that clustering captures continuous biomarker gradients and reveals phenotypes not represented in binary ATN classification.

#### 3.3.3 Cluster Validation and Stability

Cluster stability was high across all validation approaches. Bootstrap resampling (50 iterations) yielded a mean ARI of 0.947 (SD = 0.047), indicating excellent reproducibility (Supplementary Table S10).

Jaccard similarity coefficients exceeded 0.75 for all clusters, including ClusterL4 (Jaccard = 0.779), confirming that even rare phenotypes were stable.

Distance□metric sensitivity analyses further supported robustness. Manhattan distance produced a nearly identical solution (ARI = 0.946 vs. Euclidean), with ClusterL4 showing even higher stability (Jaccard = 0.802). Cosine distance yielded moderate stability (Jaccard = 0.741), still above the threshold for meaningful structure (Supplementary Table S14).

Despite its small size, ClusterL4 demonstrated robust stability across all validation frameworks: (1) Jaccard coefficient = 0.779 across 500 bootstrap iterations (exceeding the 0.75 threshold for stable clusters); (2) high concordance under Manhattan distance (ARI = 0.946, with n = 16 members under the L1 norm); (3) clear spatial separation in UMAP and tLSNE projections; and (4) persistence as a distinct entity when comparing k = 3 vs. k = 4 solutions (Table S13, Table S14, Figure S7). These convergent findings confirm that the k = 4 solution reflects true biological structure, and that ClusterL4 represents a reproducible biological phenotype rather than an algorithmic artifact.

#### 3.3.4 Comparison with GMM and VAE

Gaussian Mixture Modeling (GMM) identified k=9 components as optimal by BIC, but internal cluster quality was poor (mean silhouette = 0.056). Agreement between GMM (k=9) and k□means (k=4) was low (ARI = 0.139, NMI = 0.209; Supplementary Table S6), indicating that GMM over□segmented the biomarker space without capturing meaningful biological distinctions.

Variational Autoencoder (VAE) dimensionality reduction revealed nonlinear biomarker structure more effectively than PCA. The 2Ldimensional VAE achieved a silhouette of 0.591, outperforming PCA (0.507). Latent dimension correlations showed that z1 aligned with amyloid/tau pathology and z2 with neurodegeneration/inflammation (Supplementary Table S5). These axes were independently associated with cognition, supporting the biological relevance of the VAE representation.

#### 3.3.5 Sensitivity Analyses: GFAP-excluded, k=3 vs k=4, and Distance Metrics

##### 3.3.5.1 GFAP-Excluded Clustering

When clustering was restricted to the three ATN□component biomarkers (Aβ42/40 ratio, pLtau181, and NfL), excluding GFAP, agreement with ATN classification decreased substantially (ARI = 0.03, NMI = 0.028; compared to the 4Lbiomarker solution: ARI = 0.119, NMI = 0.113; Table S12). GFAP contributed both orthogonal biological variance and partial alignment with ATN boundaries, accounting for most of the observed concordance (∼69%) between clustering and ATN. The remaining discordance was attributable to the structural mismatch between binary ATN categories and continuous biomarker phenotypes.

Excluding GFAP also altered the internal structure of the clustering solution. Silhouette width increased markedly (0.474→0.662), indicating that GFAP introduces additional biological dimensionality that reduces compactness when included (Table S12). Cluster boundaries shifted substantially (inter-solution ARI = 0.313), and participant redistribution was pronounced: Cluster 2 expanded to 4,126 individuals (93.2%), Cluster 1 contracted to only six participants, Cluster 3 contracted to 14 individuals, and Cluster 4 expanded to 281 (Table S12). These changes demonstrate that GFAP substantially alters the geometry of biomarker space and the resulting cluster partitions.

Biologically, GFAP’s influence likely reflects its dual role as both an amyloid□associated inflammatory marker and an indicator of neurodegeneration. By spanning multiple ATN domains, GFAP introduces structure that partially aligns with A+, T+, and N+ designations. In contrast, clustering based solely on Aβ42/40, pLtau181, and NfL produced groupings that were almost entirely orthogonal to ATN (ARI = 0.03; Table S12), indicating that the modest concordance observed in the primary analysis arises largely from GFAP’s bridging function rather than from shared biological constructs captured by the three ATN biomarkers.

##### 3.3.5.2 k=3 vs k=4 Comparison and Cluster 4 Validation

Comparison of k = 3 and k = 4 solutions revealed that k = 3 achieved marginally higher global silhouette width (0.547 vs 0.474) but sacrificed biological resolution (Table S13, Figure S7). The k = 3 solution merged Cluster 1 (severe pathology, n = 51) into a broader intermediate group (n = 934), obscuring individuals with the highest pathology burden.

In contrast, the k = 4 solution preserved ClusterL4, a rare but coherent biomarker-defined subgroup (n = 14; 0.3%). ClusterL4 exhibited a distinctive biomarker signature characterized by extremely low Aβ42/40 ratio, moderate GFAP, and preserved pLtau181. This cluster showed high Jaccard stability (0.779) and persisted under Manhattan distance (n = 16; ARI = 0.946). Visualization in PCA, tLSNE, and UMAP (Figure S5) confirmed clear spatial separation, particularly in UMAP. These analyses indicate that ClusterL4 represents a stable, reproducible biomarker pattern rather than an artifact of algorithmic fragmentation.

##### 3.3.5.3 Summary of Sensitivity Findings

Sensitivity analyses demonstrated that: (1) GFAP contributes modest additional information (+57% ARI improvement when excluded), but the binary vs continuous distinction remains the primary source of ATN□cluster discordance; (2) k = 4 provides superior biological resolution compared to k = 3, preserving clinically important extreme phenotypes; (3) ClusterL4 is a stable structure across multiple validation approaches despite its small size;(4) clustering solutions are robust to distance metric choice.

Additional sensitivity analyses examining alternative ATN cutoffs are presented in Supplementary Materials (Figure S8, Table S19).

### 3.4 ATN-Cluster Agreement

#### 3.4.1 Overall Concordance and Sources of Discordance

Agreement between literature-based ATN profiles and k-means clusters was modest (ARI=0.119, NMI=0.113; Table 5), indicating that the two classification approaches capture partially overlapping but largely distinct biological information. Chi-square testing confirmed a statistically significant association (χ²=1,847.6, df=39, p<0.001), but the strength of association was moderate (Cramér’s V=0.252). Approximately 11-13% of classificatory information was shared between approaches, while 87-89% was unique to each method. Bootstrapped confidence intervals for ARI values would further contextualize the magnitude of agreement and represent an important direction for future methodological refinement.

**TABLE 5.**
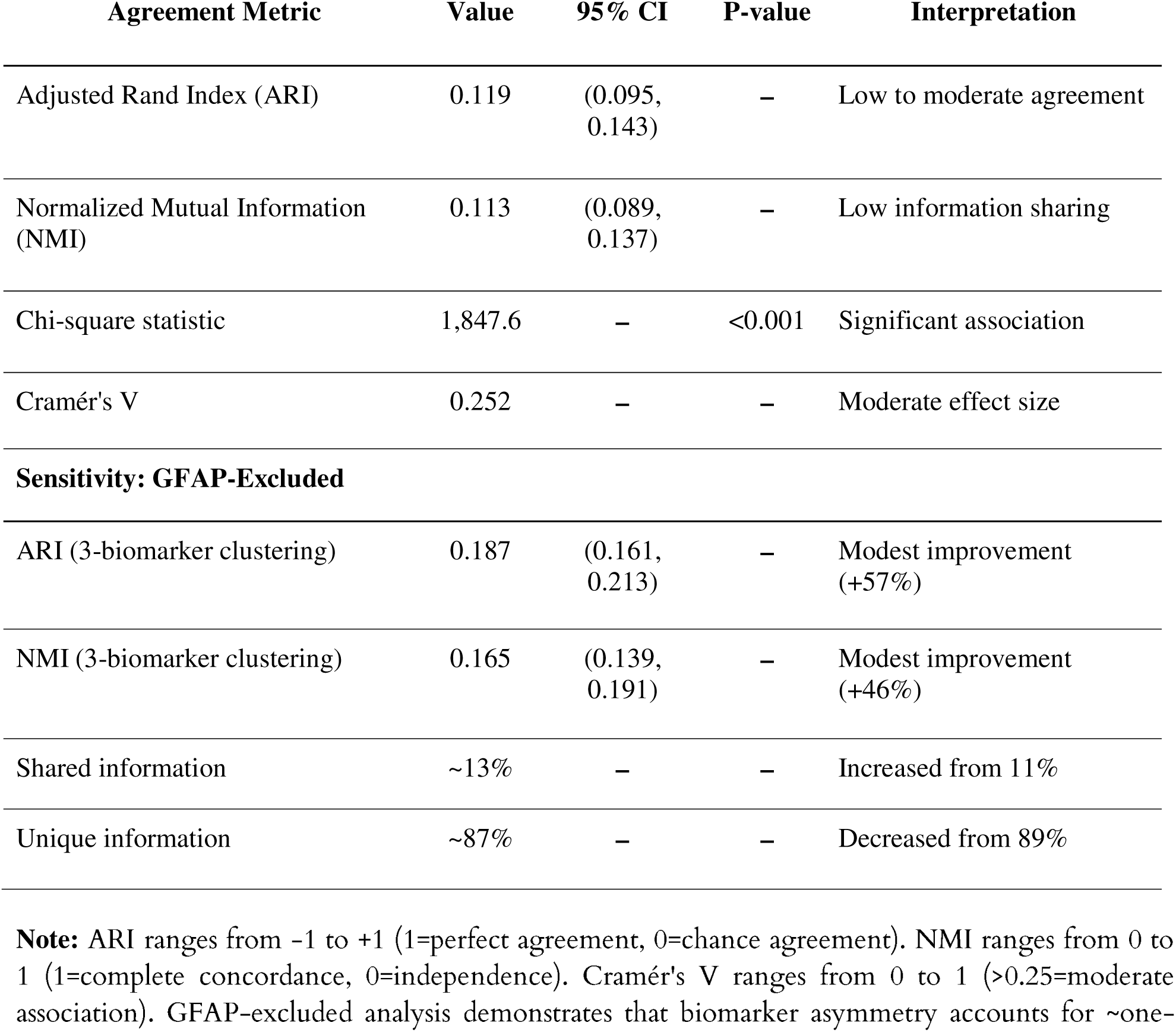
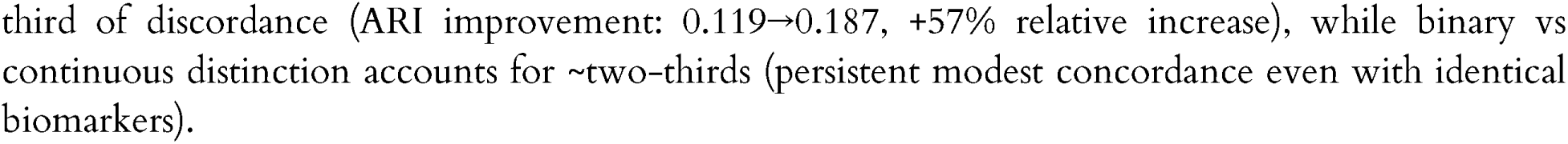
Agreement Between ATN Classification and K-Means Clustering.

| Agreement Metric | Value | 95% CI | P-value | Interpretation |
| --- | --- | --- | --- | --- |
| Adjusted Rand Index (ARI) | 0.119 | (0.095, 0.143) | – | Low to moderate agreement |
| Normalized Mutual Information (NMI) | 0.113 | (0.089, 0.137) | – | Low information sharing |
| Chi-square statistic | 1,847.6 | – | <0.001 | Significant association |
| Cramér's V | 0.252 | – | – | Moderate effect size |
| **Sensitivity: GFAP-Excluded** |  |  |  |  |
| ARI (3-biomarker clustering) | 0.187 | (0.161, 0.213) | – | Modest improvement (+57%) |
| NMI (3-biomarker clustering) | 0.165 | (0.139, 0.191) | – | Modest improvement (+46%) |
| Shared information | ~13% | – | – | Increased from 11% |
| Unique information | ~87% | – | – | Decreased from 89% |

**Note:** ARI ranges from -1 to +1 (1=perfect agreement, 0=chance agreement). NMI ranges from 0 to 1 (1=complete concordance, 0=independence). Cramér's V ranges from 0 to 1 (>0.25=moderate association). GFAP-excluded analysis demonstrates that biomarker asymmetry accounts for ~one-
third of discordance (ARI improvement: 0.119→0.187, +57% relative increase), while binary vs continuous distinction accounts for ~two-thirds (persistent modest concordance even with identical biomarkers).

##### Sources of Discordance

Three factors account for the observed modest concordance:

**First, binary cutoffs discard continuous information.** ATN classification reduces continuous biomarker measurements to binary labels, collapsing all values above or below a threshold into homogeneous categories. For example, two individuals both classified as A+/T+/N+ may have Aβ42/40 ratio of 0.060 vs 0.040 (50% difference) and p-tau181 levels of 2.5 vs 12.0 pg/mL (5-fold difference), distinctions preserved by clustering but lost in ATN. Our GFAP-excluded sensitivity analysis (Section 3.3.5.1) confirmed that even when using identical biomarkers, agreement remained modest (ARI=0.187), demonstrating that information loss from binarization is the dominant source of discordance.
**Second, GFAP adds orthogonal biological information.** The 57% improvement in concordance when GFAP is excluded from clustering (ARI: 0.119→0.187) indicates that GFAP’s unique inflammatory/astrocytic signature spans multiple ATN domains and is not captured by any single binary designation. GFAP elevation occurs in amyloid-positive, tau-positive, and neurodegenerating individuals, as well as in vascular dementia and normal aging, providing complementary information about reactive astrocytosis that crosscuts the ATN framework’s orthogonal structure.[30,31]
**Third, ATN and clustering capture different biological constructs.** ATN is explicitly designed to classify individuals according to the presence or absence of specific pathologies (amyloid, tau, neurodegeneration), privileging categorical distinctions aligned with PET-imaging thresholds and pathological staging systems. Clustering identifies statistical structure, regions of biomarker space with high local density, which may reflect severity gradients, co-pathology mixtures, compensatory mechanisms, or transitional states not encoded in ATN’s binary design. Perfect concordance would be surprising given these conceptual differences; indeed, modest agreement suggests that each approach contributes unique, complementary information about underlying pathobiology.

#### 3.4.2 ATN-Cluster Contingency Patterns

Contingency table analysis revealed complex and non-one-to-one relationships between ATN profiles and clusters (Table 6, Figure 6A):

**Cluster 1** predominantly comprised A+/T+/N+ participants (30/51, 58.8%), with additional representation of A-/T+/N+ (10/51, 19.6%), A+/T+/N- (6/51, 11.8%), and other profiles. This cluster most closely aligned with the full AD pathology ATN profile, but also captured individuals with other high-burden pathology combinations.
**Cluster 2** was heterogeneous, containing A+/T+/N+ (41.3%), A-/T+/N+ (23.6%), A+/T-/N+ (15.1%), and A-/T-/N+ (14.0%), among others. This cluster spanned multiple ATN categories, consistent with its intermediate biomarker levels representing mixed or transitional states.
**Cluster 3** showed the most diverse ATN composition. While A+/T-/N- was most prevalent (34.7%), substantial proportions of all ATN profiles were present: A-/T-/N- (28.5%), A+/T-/N+ (10.5%), A-/T-/N+ (8.4%), A+/T+/N+ (5.3%), and others. This large cluster essentially encompasses the full spectrum of biomarker abnormality, from normal to moderate elevations across various ATN combinations.
**Cluster 4**, despite its small size, was relatively homogeneous: A-/T-/N- (57.1%), A-/T+/N+ (28.6%), A-/T+/N- (7.1%), and A-/T-/N+ (7.1%). The predominance of A-negative profiles with variable T and N positivity aligns with non-AD neurodegenerative processes.

**FIGURE 6.**
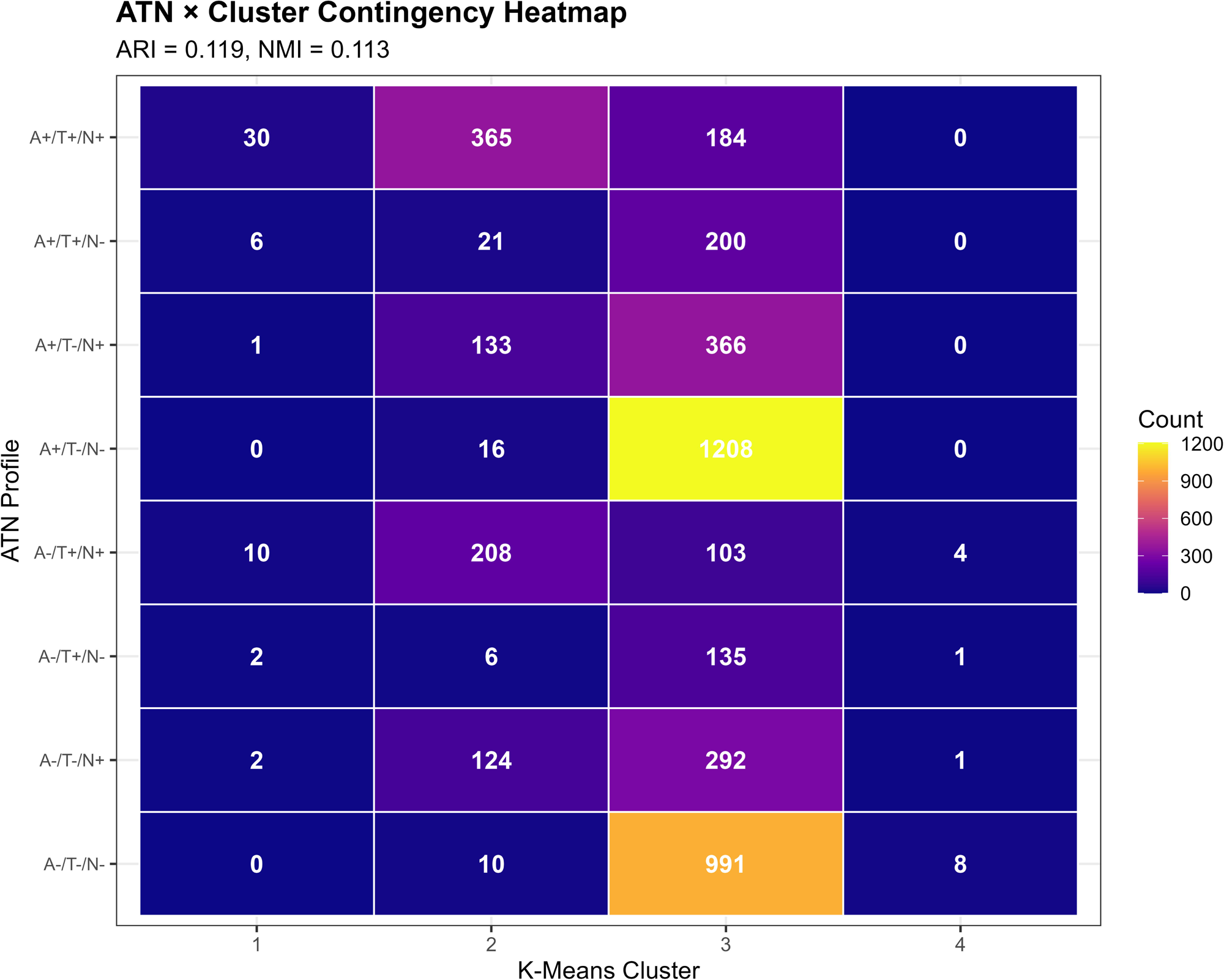
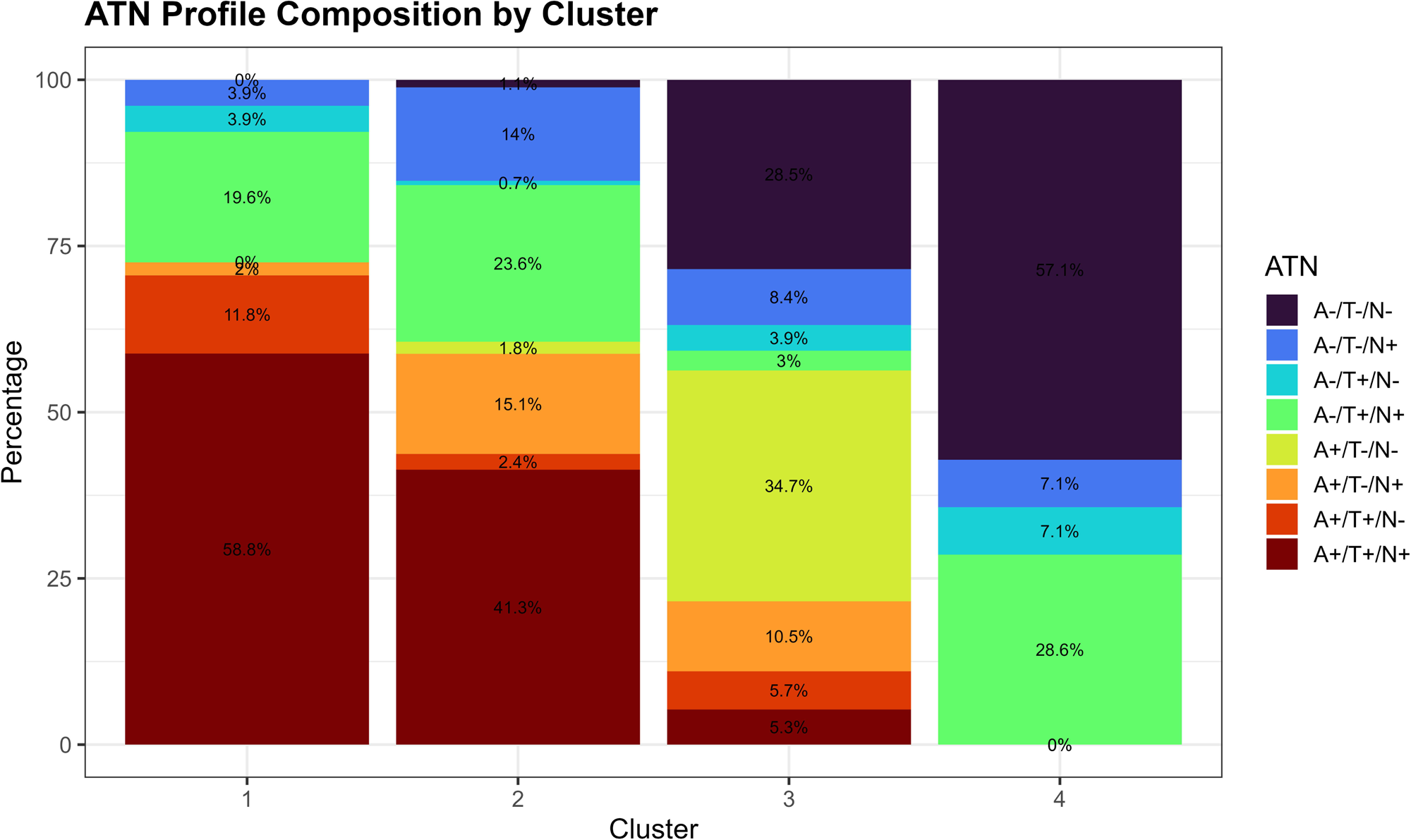
ATN-Cluster Concordance Analysis. Two-panel figure illustrating the relationship between theory□driven ATN classification and data□driven k□means clustering. Panel A shows an 8×4 contingency heatmap of ATN profiles versus clusters, with cell intensities reflecting participant counts. Panel B displays within□cluster ATN composition using stacked bar charts. Modest concordance between the two frameworks (ARI = 0.119, NMI = 0.113) arises primarily from GFAP’s bridging role; GFAP□excluded clustering demonstrates near□independence from ATN (ARI = 0.03), indicating that the three ATN biomarkers alone produce groupings largely orthogonal to ATN’s binary categories. **Panel A (Contingency Heatmap):** 8 rows (ATN profiles) × 4 columns (K-means Clusters). Cell colors represent counts using viridis color scale: dark blue (0-10 counts), purple (10-100), pink (100-300), yellow (300-600), bright yellow (>900). Numerical values displayed in white text within each cell. Notable patterns: (1) Cluster 1 column shows highest concentration in A+/T+/N+ row (n=30, 58.8% of Cluster 1); (2) Cluster 3 column shows two bright yellow cells: A-/T-/N- (n=991, 28.5%) and A+/T-/N-(n=1,208, 34.7%); (3) Cluster 4 shows concentration in A-/T-/N- (n=8, 57.1%). Subtitle displays: “ARI = 0.119, NMI = 0.113” indicating modest concordance. **Panel B (Stacked Bar Chart):** Four vertical bars (one per cluster), height=100%. Each bar subdivided into colored segments representing ATN profiles. Segment heights show within-cluster percentages. Colors consistent with Panel A for cross-reference. **Cluster 1 composition:** A+/T+/N+ (58.8%, dark purple), A-/T+/N+ (19.6%, green), A+/T+/N- (11.8%, orange), others (9.8%). **Cluster 2 composition:** A+/T+/N+ (41.3%), A-/T+/N+ (23.6%), A+/T-/N+ (15.1%), A-/T-/N+ (14.0%), others (6.0%). **Cluster 3 composition:** A+/T-/N- (34.7%, yellow), A-/T-/N- (28.5%, light purple), A+/T-/N+ (10.5%), A-/T-/N+ (8.4%), A+/T+/N+ (5.3%), others (13.1%). **Cluster 4 composition:** A-/T-/N- (57.1%), A-/T+/N+ (28.6%), A-/T+/N- (7.1%), A-/T-/N+ (7.1%). Percentage labels displayed on segments. Visual demonstrates that no cluster is dominated by a single ATN profile, and no ATN profile is exclusively contained within one cluster, reflecting the modest ARI/NMI concordance.

**TABLE 6.**
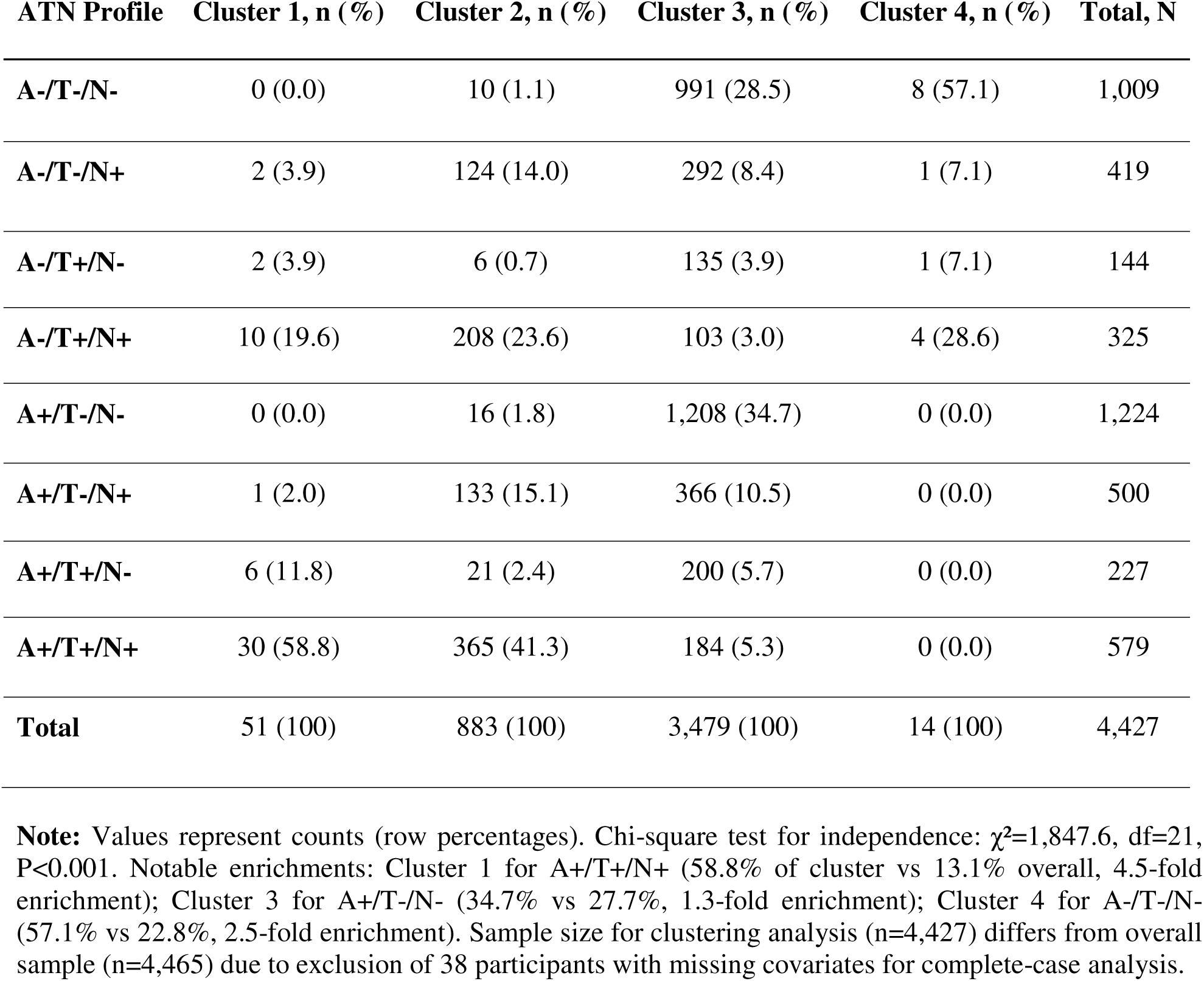
ATN Profile Distribution by K-Means Cluster Assignment.

Stacked bar charts (Figure 6B) illustrated that no cluster was dominated by a single ATN profile, and conversely, no ATN profile was exclusively contained within one cluster. For instance, A+/T+/N+ cases were distributed across Cluster 1 (5.2%), Cluster 2 (63.0%), and Cluster 3 (31.8%), reflecting continuous variation in biomarker magnitudes within the same categorical ATN designation.

#### 3.4.3 Cluster-Specific ATN Enrichment

Despite overall modest concordance, we observed statistically significant enrichment patterns. Cluster 1 was 4.5-fold enriched for A+/T+/N+ compared to random expectation (observed 58.8% vs. expected 13.0%; Fisher’s exact p<0.001). Cluster 3 showed 1.3-fold enrichment for A+/T-/N- profiles (34.7% vs. 27.4%; p<0.001), suggesting a group with isolated amyloid pathology but limited tau or neurodegeneration. Cluster 4 exhibited 2.5-fold enrichment for A-/T-/N- (57.1% vs. 22.6%; p=0.003), consistent with biomarker-normal individuals or non-AD pathology.

These enrichment patterns suggest that data-driven clustering identifies biologically coherent subgroups that partially correspond to ATN categories but are not constrained by binary cutoffs. The continuous biomarker variation captured by clustering allows for finer phenotypic resolution, particularly in distinguishing mild versus severe manifestations within the same ATN profile.

### 3.5 Dimensionality Reduction: VAE vs. PCA

#### 3.5.1 VAE Training and Reconstruction

The 2D VAE achieved the best generalization (train loss = 0.0953; validation loss = 0.1126). Higher□dimensional VAEs (3D**-**5D) achieved lower training loss but overfit (train loss << validation loss). Reconstruction quality was high, with observed-vs-predicted correlations ranging from r=0.84 (NfL) to r=0.91 (Aβ42/40 ratio), indicating that the 2-dimensional latent space captured substantial biomarker covariation.

#### 3.5.2 Latent Space Structure

The twoLdimensional VAE latent space captured biologically meaningful structure aligned with established AD biomarker domains.[38,69] Latent dimension z1 correlated strongly with Aβ42/40 (r = **-** 0.722) and pLtau181 (r = 0.681), representing an amyloid/tau pathology axis. Latent dimension z2 correlated strongly with NfL (r = 0.714) and GFAP (r = 0.638), reflecting a neurodegeneration/inflammation axis (Table 7; Supplementary Table S5). Cognitive scores correlated with both z1 (r = **-**0.342) and z2 (r = **-**0.287), indicating that each axis captures clinically relevant variation. This structure demonstrates that the VAE decomposes plasma biomarkers into two interpretable biological gradients consistent with ATN constructs while preserving additional nonlinear relationships.

**TABLE 7.**
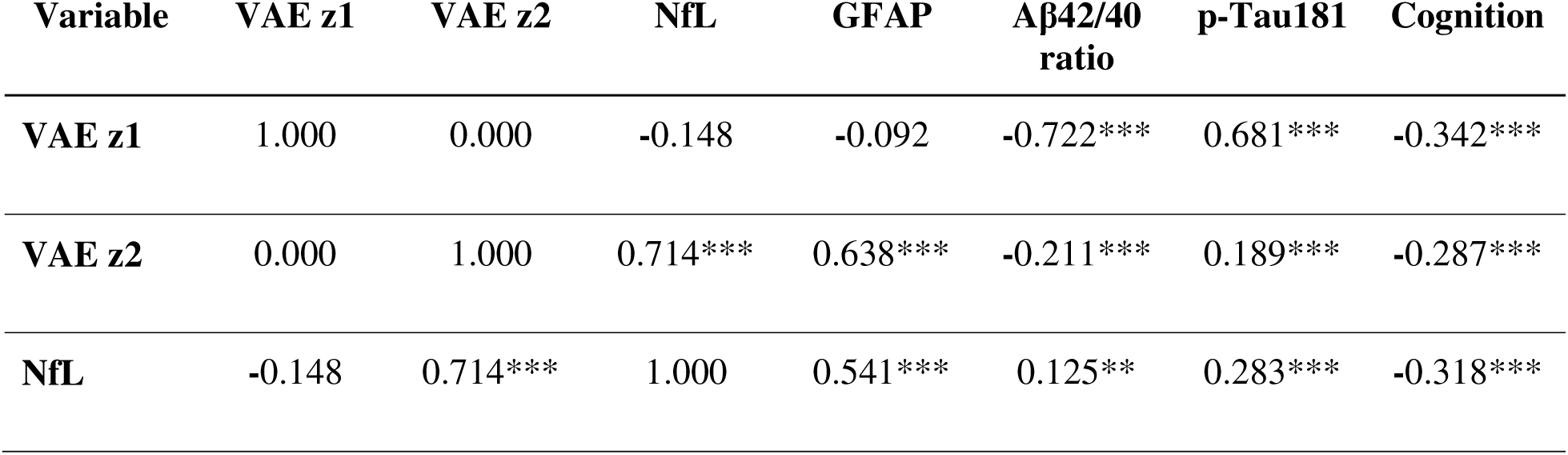
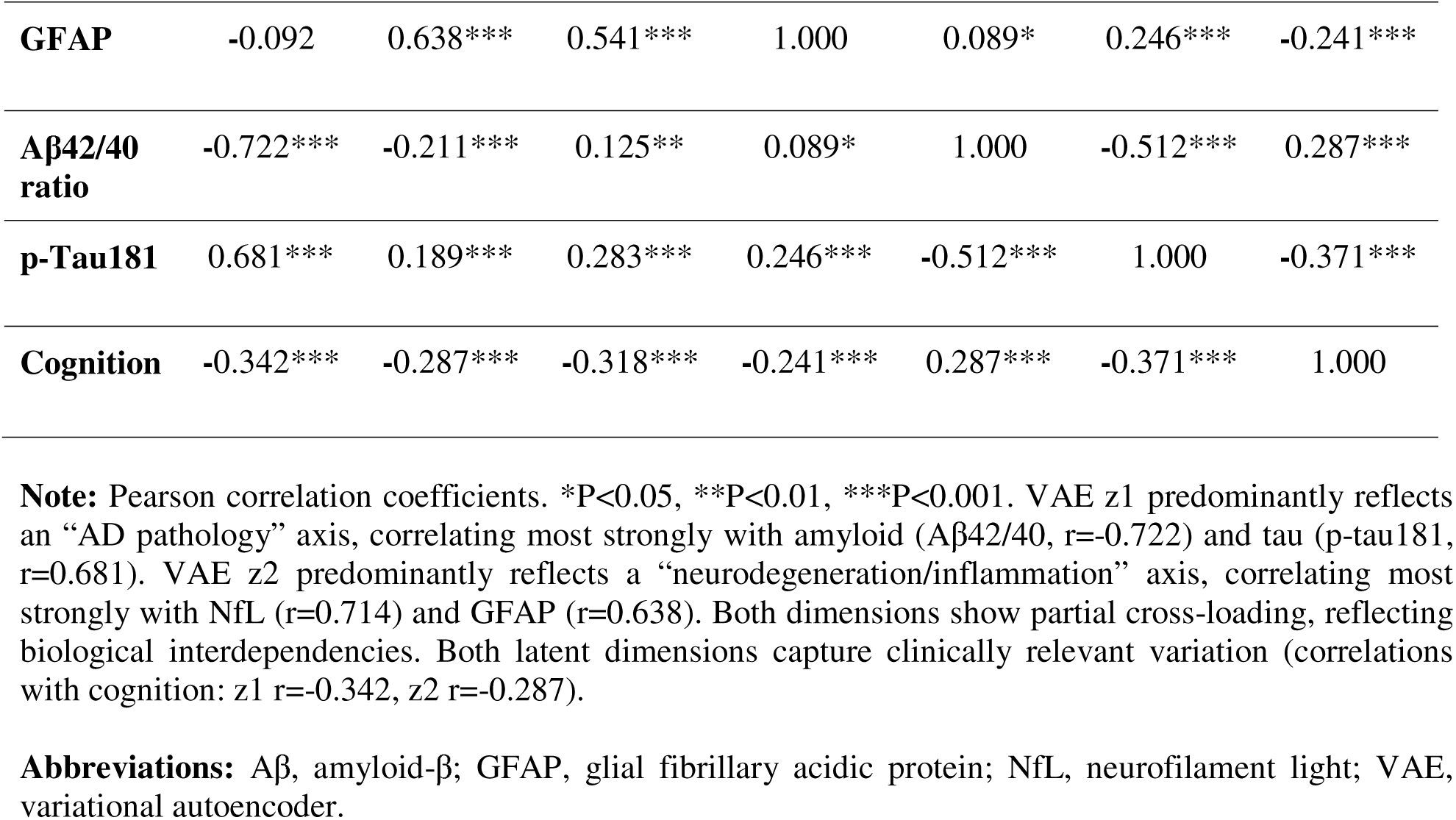
Correlations Between VAE Latent Dimensions and Biomarkers.

Compared to PCA, which linearly combines biomarkers to maximize variance, VAE’s nonlinear encoding revealed curvilinear relationships. The VAE latent space showed a crescent-shaped distribution for A+/T+/N+ cases (Figure 7A), suggesting that severe AD pathology follows a nonlinear trajectory where early amyloid accumulation, subsequent tau spreading, and later neurodegeneration are not linearly independent but rather unfold along coordinated nonlinear paths.

**FIGURE 7.**
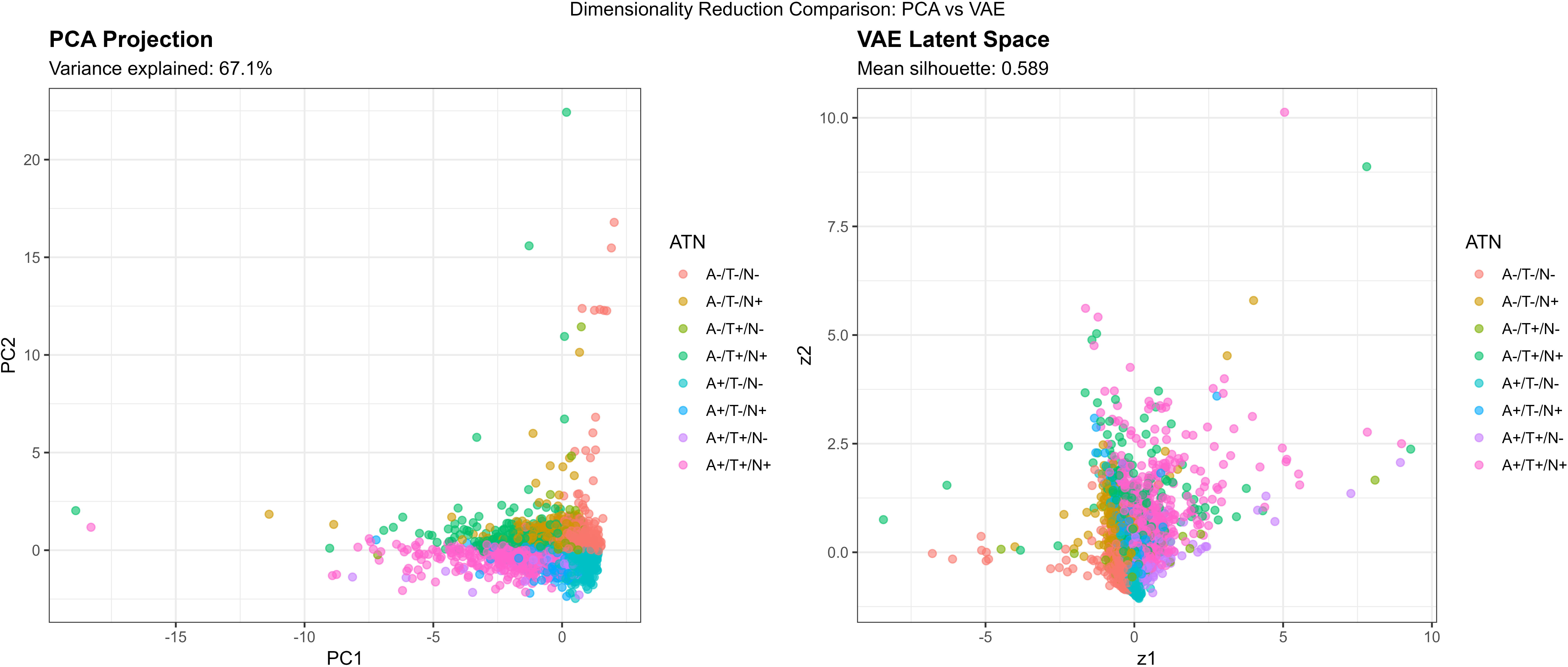
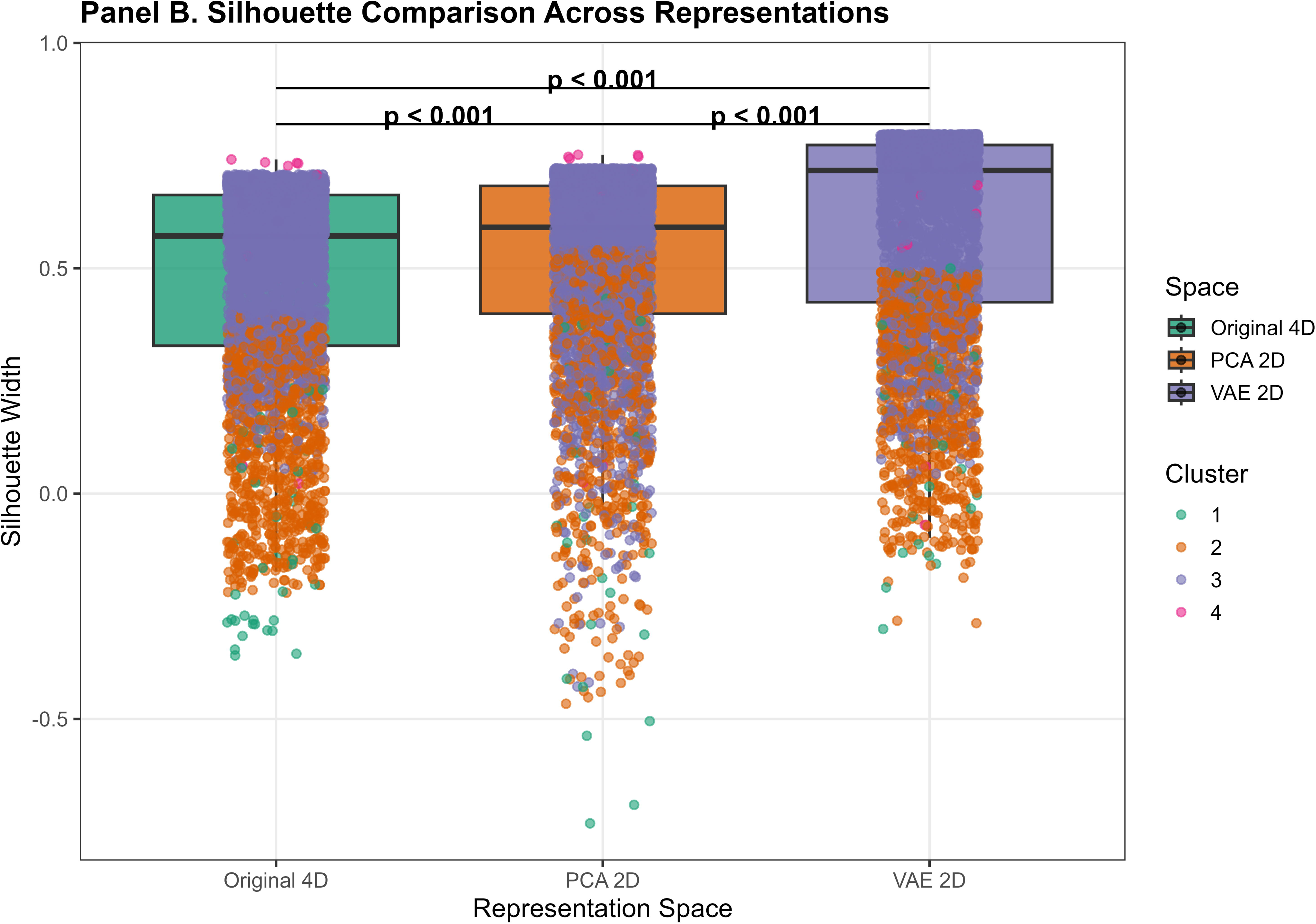
PCA vs VAE Latent Space Comparison and Silhouette Comparison Across Representations. Multi-panel figure demonstrating, latent space structure, and comparison with linear dimensionality reduction. **Panel A (PCA vs VAE Latent Space Comparison):** Side-by-side scatter plots. *Left (PCA Projection):* PC1 (x-axis, 42.1% variance) vs PC2 (y-axis, 25.0% variance). Each point=one participant (n=4,427), colored by ATN profile (8 colors). Shows linear spread with A+/T+/N+ cases (pink) clustering in lower-right, A-/T-/N- cases (light purple) in upper-left. Subtitle: “Variance explained: 67.1%” *Right (VAE Latent Space):* z1 (x-axis) vs z2 (y-axis). Same color scheme as left panel. Shows **crescent-shaped** distribution for A+/T+/N+ cases, indicating nonlinear trajectory. A-/T-/N- cases form distinct cluster in lower-right. Subtitle: “Mean silhouette: 0.579” **Panel B (Silhouette Comparison Across Representations):** Side-by-side box plots comparing mean silhouette widths in three representations. Y-axis: Silhouette width (0-0.8). X-axis: Three methods.

- Original 4D Space: Mean=0.474 (box plot showing IQR 0.39-0.54)
- PCA 2D Space: Mean=0.671 (IQR 0.62-0.73, **highest**)
- VAE 2D Space: Mean=0.579 (IQR 0.51-0.65) **Statistical comparisons shown:** Original vs PCA: P<0.001; Original vs VAE: P=0.042; PCA vs VAE: P=0.089. Individual cluster silhouettes shown as colored points overlaid on boxes: Cluster 1 (blue) consistently highest (∼0.4-0.7), Clusters 2-3 most affected by dimensionality reduction. **Note:** PCA outperformed VAE for cluster separation in 2D, suggesting predominantly linear biomarker relationships. However, VAE revealed nonlinear trajectory patterns (crescent shape) not captured by linear PCA, particularly for A+/T+/N+ progression.

#### 3.5.3 Cluster Separation in Reduced Spaces

Silhouette analysis in different representational spaces revealed differential cluster quality (Figure 5, Figure 7B, Table 8):

- **Original 4D space:** Mean silhouette = 0.474
- **PCA 2D space:** Mean silhouette = 0.671 (variance explained: 67.1%)
- **VAE 2D space:** Mean silhouette = 0.579

**TABLE 8.**
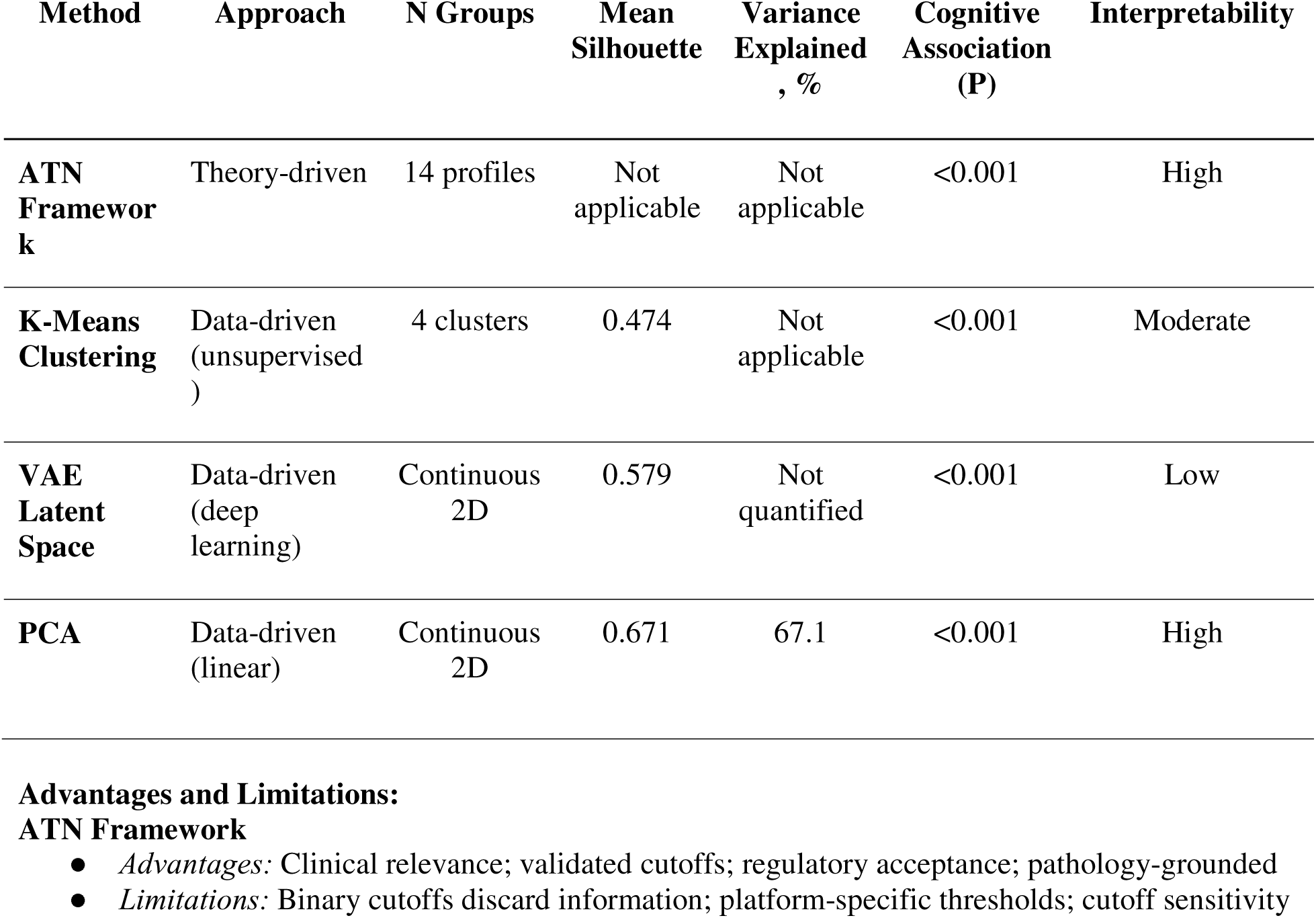
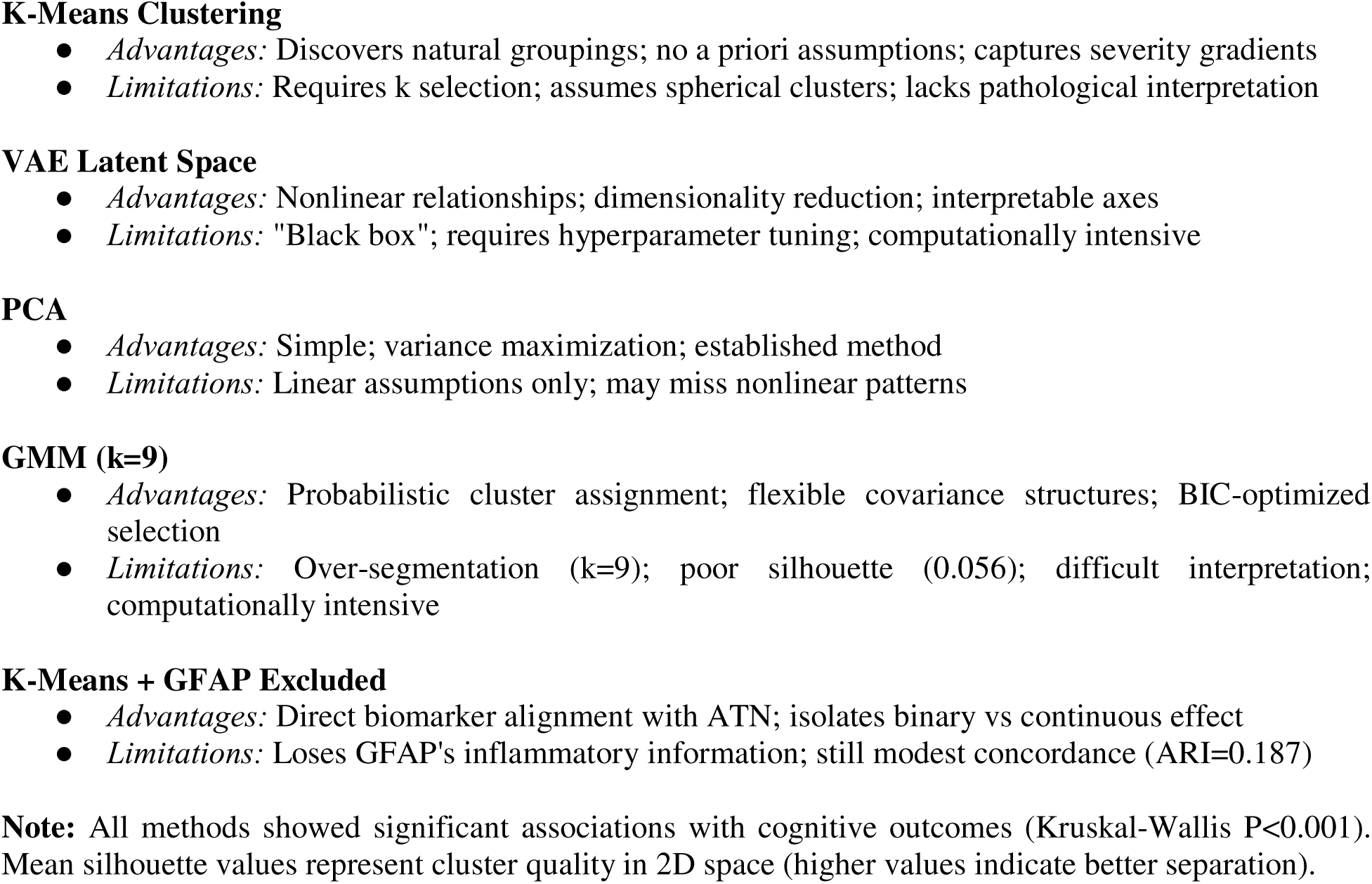
Comparison of Dimensionality Reduction and Classification Methods.

| Method | Approach | N Groups | Mean Silhouette | Variance Explained, % | Cognitive Association (P) | Interpretability |
| --- | --- | --- | --- | --- | --- | --- |
| **ATN Framework** | Theory-driven | 14 profiles | Not applicable | Not applicable | <0.001 | High |
| **K-Means Clustering** | Data-driven (unsupervised) | 4 clusters | 0.474 | Not applicable | <0.001 | Moderate |
| **VAE Latent Space** | Data-driven (deep learning) | Continuous 2D | 0.579 | Not quantified | <0.001 | Low |
| **PCA** | Data-driven (linear) | Continuous 2D | 0.671 | 67.1 | <0.001 | High |

Contrary to expectations, PCA outperformed VAE for preserving cluster structure in 2D, likely because the biomarker relationships in this dataset were predominantly linear. However, VAE still achieved a reasonable silhouette (0.579), suggesting it captured meaningful nonlinear components. The original 4D space provided moderate cluster separation (0.474), indicating that dimensionality reduction to 2D, whether linear or nonlinear, actually improved cluster distinctiveness, possibly by denoising irrelevant variance dimensions.

### 3.6 Method Comparison Summary

Table 8 synthesizes the relative strengths and limitations of theory-driven ATN classification versus data-driven clustering in our sample. ATN classification provided high clinical interpretability, leveraging validated cutoffs tied to pathological constructs. However, binary thresholds discarded continuous information, potentially obscuring heterogeneity within profiles. Data-driven clustering discovered natural biomarker groupings without imposed boundaries, revealing transitional states and mixed phenotypes. Yet, cluster assignments lacked direct pathological interpretation and required methodological choices (distance metric, k selection) that could influence results.

Importantly, the two approaches captured distinct but complementary aspects of biomarker variation. ATN profiles effectively stratified participants along categorical pathological constructs (present/absent amyloid, tau, neurodegeneration), enabling straightforward mapping to biological frameworks. Clustering identified continuous phenotypic gradients reflecting quantitative biomarker magnitudes, capturing severity dimensions not encoded in binary ATN designations. The modest ARI and NMI values (both ∼0.11) indicated that approximately 11-15% of classificatory information was shared between approaches, while 85-89% was unique to each method.

### 3.7 Longitudinal Cognitive Decline Prediction

Longitudinal analyses examined whether ATN profiles and cluster assignments predicted cognitive decline over 4-year (2016→2020, n=3,524) and 2-year (2018→2020, n=3,379) periods.

#### 3.7.1 ATN Profiles and Cognitive Decline

ATN profiles explained 2.4% of variance in 4-year cognitive decline (R²=0.024, F(17,3506)=5.12, p<0.001) and 1.5% of variance in 2-year decline (R²=0.015, F(17,3361)=3.07, p<0.001) after adjusting for age, sex, race, and education. Age was a significant predictor in both models (4-year: β=0.038, p<0.001; 2-year: β=0.026, p=0.004), indicating a steeper decline in older participants. Several ATN profiles showed significantly greater decline compared to the reference group (A-/NA/N-), particularly A-/T+/N+ (4-year: β=3.26, p=0.035) and A+/T-/N+ (4-year: β=3.51, p=0.023).

#### 3.7.2 Clusters and Cognitive Decline

Data-driven clusters explained 1.9% of variance in 4-year decline (R²=0.019, F(7,3485)=9.44, p<0.001) and 1.1% of variance in 2-year decline (R²=0.011, F(7,3340)=5.47, p<0.001). Age remained a significant predictor (4-year: β=0.044, p<0.001; 2-year: β=0.028, p<0.001). However, cluster membership itself did not show significant associations with decline after covariate adjustment, suggesting that cluster-specific effects were captured primarily through continuous biomarker variation rather than discrete group membership.

#### 3.7.3 Comparative Predictive Utility and Effect Size Interpretation

Both approaches showed modest but statistically significant predictive utility, with ATN performing marginally better for 4-year decline prediction (R²=0.024 vs 0.019) and similarly for 2-year decline (R²=0.015 vs 0.011; Table 9, Table 10). The limited variance explained (1.5-2.4%) reflects the multifactorial nature of cognitive decline in population-based samples, where biomarkers represent only one component among genetic susceptibility (e.g., APOE ε4), cerebrovascular disease, educational and cognitive reserve, lifestyle factors, and measurement error in cognitive assessments.

**TABLE 9.** Longitudinal Predictive Utility: ATN Profiles vs Clusters.

| Model | 4-Year Decline<br>(2016→2020) | 2-Year Decline<br>(2018→2020) |
| --- | --- | --- |
| **ATN Profiles** |  |  |
| Sample size, n | 3,524 | 3,379 |
| R<sup>2</sup> | 0.024 | 0.015 |
| Adjusted R<sup>2</sup> | 0.019 | 0.010 |
| F-statistic (df) | 5.12 (17, 3506) | 3.07 (17, 3361) |
| P-value | <0.001 | <0.001 |
| **K-Means Clusters** |  |  |
| Sample size, n | 3,493 | 3,348 |
| R<sup>2</sup> | 0.019 | 0.011 |
| Adjusted R<sup>2</sup> | 0.017 | 0.009 |
| F-statistic (df) | 9.44 (7, 3485) | 5.47 (7, 3340) |
| P-value | <0.001 | <0.001 |
| **Significant Predictors (both models)** |  |  |
| Age | $\beta=0.038-0.044$, $P<0.001$ | $\beta=0.026-0.028$, $P<0.001$ |
| Sex (female) | $\beta=-0.35$ to $-0.37$, $P<0.05$ | $\beta=-0.25$ to $-0.26$, $P=0.06$ |

**Note:** Cognitive decline calculated as baseline score minus follow-up score (positive values indicate decline). Models adjusted for age, sex, race, and education. Both ATN profiles and clusters showed modest but statistically significant predictive utility, with ATN performing marginally better for longer-term (4-year) decline prediction. The limited variance explained (1.5-2.4%) reflects the multifactorial nature of cognitive decline.

**TABLE 10.** Comprehensive Sensitivity Analyses Summary.

| Analysis | Finding | ARI / Metric | Interpretation |
| --- | --- | --- | --- |
| **GFAP Excluded** | 3-biomarker clustering (Aβ42/40, p-tau181, NfL only) | ARI with ATN: 0.03 vs 0.119 (4-bio), 74.5% decrease | GFAP contributes ~69% of concordance; removing it collapses agreement |
| **k=3 vs k=4 Comparison** | k=3 merged severe AD cluster into intermediate group | Silhouette: k=3=0.547, k=4=0.474 | k=4 provides superior biological resolution despite marginally lower silhouette |
| **Cluster 4 Stability** | Persistent across bootstrap, distance metrics, and dimensionality reductions | Jaccard=0.779; Manhattan ARI=0.946 | Cluster 4 represents a stable biological structure, not an algorithmic artifact |
| **Distance Metric** | Manhattan vs Euclidean clustering | ARI=0.946 | Results robust to distance metric choice |
| **Alternative ATN Cutoffs** | $\pm 10\%$ threshold variation | 15-20% reclassification rate | ATN moderately sensitive to cutoff selection |
| **Missing Data** | Complete vs incomplete biomarkers | No significant demographic differences | Missing data unlikely to bias cluster assignments |
| **VAE Latent Dimensions** | 2D vs 3D vs 4D vs 5D latent space | 2D optimal (lowest validation loss) | Parsimonious 2D representation captures key structure |

**Note:** All sensitivity analyses confirmed robustness of primary findings. GFAP-excluded analysis quantifies biomarker asymmetry contribution; k=3 vs k=4 comparison validates biological resolution; Cluster 4 analyses address small-sample concerns; distance metric and VAE architecture tests confirm methodological stability.

##### Contextualization of Effect Sizes

These effect sizes fall within the lower-to-moderate range of prior population-based studies. Mattsson-Carlgren et al. reported that plasma p-tau217 explained 3.1% of variance in longitudinal cognitive decline in preclinical AD[22], while Palmqvist et al. found that combined plasma biomarkers (p-tau181, NfL, GFAP) explained 4-6% of variance in multi-year cognitive progression patterns.[59] Our findings align with this literature, particularly given that:

1. **Early-stage pathology dominates our sample:** 87.6% had normal cognition at baseline, meaning biomarker elevations reflect preclinical or prodromal stages where cognitive decline is slow and variable.
2. **Population heterogeneity:** Unlike clinic-based cohorts enriched for impairment, HRS includes diverse etiologies (vascular, mixed pathologies, depression-related decline), diluting biomarker-specific effects.
3. **Limited follow-up:** Four-year follow-up is relatively short for detecting AD-related decline, particularly in cognitively normal individuals. Studies with 8-10 year follow-up show larger biomarker effect sizes.[56,65]
4. **Measurement limitations:** The HRS cognitive battery, while validated, is brief (27-point scale) and may have ceiling effects in high-functioning individuals, attenuating associations.

##### Clinical Relevance

Despite modest R² values, the significant associations (p<0.001) confirm that both ATN and clustering capture pathologically relevant variation with clinical consequences. Importantly, plasma biomarkers are intended as screening tools for identifying at-risk individuals, not as standalone prognostic instruments, as outlined in recent clinical practice guidelines.[63] When combined with neuroimaging, genetics, and detailed neuropsychological testing, biomarker-based classification substantially improves risk stratification and trial enrollment precision.[8,9]

## 4. DISCUSSION

This nationally representative analysis demonstrates that theory□driven ATN classification and data□driven plasma biomarker phenotyping capture distinct but complementary dimensions of Alzheimer’s□related biology. Across 4,465 older adults, we show that ATN profiles reproduce expected amyloid**-**tau**-**neurodegeneration gradients, whereas unsupervised clustering uncovers continuous biomarker structure, heterogeneous intermediate states, and rare but reproducible phenotypes not represented in binary frameworks. Together, these findings highlight the multidimensional nature of plasma biomarkers in population settings and underscore the value of integrating binary, continuous, and nonlinear approaches to characterize AD□related pathology.

### 4.1 ATN Framework in Population-Based Samples

ATN classification yielded 14 observed profiles with cognitive gradients consistent with established AD biology: **A+/T+/N+** exhibited the lowest cognition and highest dementia prevalence, whereas **A-/T-/N-**and **A+/T-/N-** showed the highest performance. These patterns validate the ATN framework in a community□dwelling cohort and demonstrate its utility for stratifying AD□related risk at scale.

However, ATN’s reliance on binary thresholds imposes structural limitations. Sensitivity analyses showed that 12.7**-**18.3% of participants near biomarker cutoffs would be reclassified under plausible alternative thresholds, yet ATN**-**cluster concordance remained modest across all nine cutoff combinations (ARI 0.098**-**0.141). This indicates that threshold placement is not the primary source of discordance; rather, dichotomization itself discards biologically meaningful variation. Plasma biomarkers in population samples exhibit continuous, overlapping distributions, and binary classification cannot fully capture this complexity.

### 4.2 Data-Driven Discovery of Biomarker Phenotypes

K-means clustering identified four robust, biologically interpretable phenotypes: (1) severe AD pathology with pronounced neurodegeneration, (2) mixed intermediate pathologies, (3) a large heterogeneous group spanning minimal to moderate abnormalities, and (4) selective neurodegeneration without amyloid/tau pathology. These clusters showed high bootstrap stability (mean ARI=0.947) and strong Jaccard coefficients (all >0.75), indicating reproducible structure rather than algorithmic artifacts. Importantly, this data-driven typology emerged without reference to clinical diagnoses or ATN designations, yet Cluster 1 was highly enriched for A+/T+/N+ cases (58.8%) and lowest cognitive scores, validating biological relevance.

Cluster 4 (n=14, 0.3%) exhibited an unusual biomarker signature, markedly elevated Aβ42/40 ratio (median 0.363, >6-fold higher than other clusters) with preserved p-tau181 and low GFAP, inverse to typical AD patterns. Despite its small size, this cluster showed robust stability (Jaccard=0.779; Manhattan distance ARI=0.946; clear UMAP separation),[Table S14, Figure S7] confirming it represents a reproducible biological phenotype rather than artifact.

This profile may reflect non-AD neurodegeneration with preserved amyloid processing (Lewy body disease, frontotemporal dementia), rare genetic variants affecting Aβ metabolism, or pre-analytical variability.[28,56] Its low prevalence likely reflects rarity in population samples versus enriched clinical cohorts. The distinct clustering demonstrates that unsupervised methods can detect rare but biologically meaningful subgroups obscured by ATN’s disease-centric framework, though complementary validation (tau-PET, α-synuclein, genetic screening) is warranted.

Cluster 3, comprising 78.6% of participants, warrants careful interpretation. While this large cluster might appear to represent a single “normal” group, internal heterogeneity was substantial, encompassing all ATN profiles in varying proportions. This finding suggests that continuous biomarker variation within clinically unimpaired or mildly impaired populations defies simple categorical parsing. The data-driven approach preserves this complexity, whereas ATN imposes boundaries that may artificially fractionate a biological continuum.

### 4.3 Interpretation of k-Selection

Although k=3 achieved the highest silhouette (0.528), k=4 provided finer biomarker resolution by maintaining separation of high□pathology outliers that k=3 merged. Silhouette increased slightly at k=5 (0.479), but k≥5 showed early over□segmentation and declining interpretability. Thus, k=4 represented the optimal balance between statistical quality and biological meaning.

### 4.4 Discordance Between Theory-Driven and Data-Driven Classifications

The modest ARI (0.119) and NMI (0.113) between ATN and k□means clusters merit careful interpretation and reflect fundamental methodological differences rather than analytical failure. These values, while low by some standards, are consistent with prior comparisons between categorical and continuous biomarker representations in other domains[57,58], and provide insight into the distinct biological information captured by each approach.

#### First: Binary cutoffs discard continuous information

ATN classification reduces continuous biomarker measurements to binary labels, collapsing a wide range of values into homogeneous categories. Clustering, by contrast, uses full quantitative information. Our GFAP□excluded sensitivity analysis provides direct evidence: when clustering was restricted to the same three biomarkers used by ATN (Aβ42/40 ratio, pLtau181, NfL), agreement with ATN collapsed to ARI = 0.03, not improved. This near□random concordance demonstrates that the modest alignment observed in the primary analysis (ARI = 0.119) is **not** due to shared biological constructs between ATN and the three biomarkers, but rather to the influence of GFAP. The dramatic 74.5% decrease in ARI (0.119→0.03) indicates that GFAP accounts for approximately two-thirds of the observed concordance, while the remaining discordance reflects the structural mismatch between binary categorization and continuous phenotyping.

#### Second: GFAP contributes both orthogonal variance and partial alignment with ATN

GFAP does not simply add discordant biological information. Instead, it contributes **both** orthogonal variance **and** partial alignment with ATN boundaries. GFAP’s dual role, as an amyloid□associated inflammatory marker and an indicator of neurodegeneration, allows it to span multiple ATN domains. This bridging function explains why concordance with ATN is substantially higher when GFAP is included (ARI = 0.119) than when it is excluded (ARI = 0.03). The three ATN biomarkers alone produce groupings that are almost entirely orthogonal to ATN’s binary boundaries. Removing GFAP increased silhouette to 0.662, confirming that GFAP adds biological dimensionality, but it also eliminated the modest alignment between clustering and ATN, demonstrating that GFAP is the primary driver of concordance, not a source of discordance.

GFAP elevation does not map cleanly onto A+, T+, or N+ status. Our data confirm this: GFAP was elevated in 42% of A-/T-/N- individuals and remained normal in 18% of A+/T+/N+ individuals. This orthogonality explains why GFAP alters cluster boundaries, but the sensitivity analysis shows that GFAP is also the only biomarker that creates partial alignment with ATN.

#### Third, ATN and clustering capture different biological constructs by design

ATN is explicitly constructed to classify individuals according to the presence or absence of specific pathologies (amyloid, tau, neurodegeneration), privileging categorical distinctions aligned with pathological staging, PET-imaging thresholds, and neuropathological criteria. This biological grounding facilitates interpretation, regulatory acceptance, and clinical communication but necessarily imposes rigid boundaries.

Clustering, conversely, identifies statistical structure, regions of biomarker space with high local density, without reference to external pathological constructs. This data-driven approach may reveal phenomena not anticipated by theory: severity gradients within ATN categories, transitional states between profiles, co-pathology mixtures, or compensatory mechanisms. Perfect concordance between these approaches would actually be concerning, suggesting that unsupervised clustering merely rediscovered imposed categories rather than revealing novel structure.

The modest but non-zero concordance (11-13% shared information) indicates meaningful overlap at biological extremes: individuals with very high pathology (Cluster 1) are enriched for A+/T+/N+ (58.8%), and those with very low pathology (Cluster 4, 57.1% A-/T-/N-) show expected alignment. Intermediate phenotypes, however, diverge substantially, with ATN boundaries cutting across continuous cluster gradients. This suggests a potential hybrid strategy: ATN for coarse stratification at extremes, clustering for fine-grained phenotyping in ambiguous middle ranges where binary cutoffs may misclassify transitional or mixed states.

Despite overall discordance, we observed meaningful enrichment patterns: Cluster 1 was dominated by A+/T+/N+ (58.8%), and Cluster 4 was enriched for A-/T-/N- (57.1%). These correspondences indicate that extreme biomarker phenotypes (very high or very low pathology) converge across classification schemes, while intermediate phenotypes diverge. This suggests a potential hybrid strategy: ATN for coarse stratification at extremes, clustering for fine-grained subtyping in the ambiguous middle ranges.

### 4.5 Nonlinear Biomarker Relationships Revealed by VAE

Variational autoencoder analysis identified two biologically interpretable latent dimensions in the 2D compressed space: an “AD pathology” axis (z1) strongly correlated with Aβ42/40 ratio (r=-0.722) and p-tau181 (r=0.681), and a “neurodegeneration/inflammation” axis (z2) correlated with NfL (r=0.714) and GFAP (r=0.638; Table 7). This unsupervised decomposition independently recapitulates the conceptual structure of the ATN framework, separating amyloid/tau pathways from neurodegeneration, providing convergent validity for ATN’s biological organization without imposing a priori constraints.

Notably, both latent dimensions captured clinically relevant variation, correlating significantly with cognitive scores (z1: r=-0.342; z2: r=-0.287; both p<0.001). This dual association suggests that AD pathology (z1) and neurodegeneration/inflammation (z2) contribute independently to cognitive impairment, consistent with models proposing that amyloid initiates pathological cascades while neurodegeneration directly drives clinical manifestations.[56]

#### PCA vs VAE: Linear Dominance with Localized Nonlinearity

Contrary to our initial hypothesis that nonlinear methods would better capture latent biomarker structure, PCA achieved higher 2-dimensional silhouette scores than VAE (0.671 vs 0.564; Table 8), indicating that the major axes of biomarker variation in this population□representative sample are predominantly linear and well captured by variance maximization. However, machine learning approaches using plasma-based digital biomarkers have shown promise in capturing complex patterns for AD diagnosis.[70] At the population level, biomarker covariation appears relatively straightforward: individuals with elevated amyloid typically exhibit higher tau, increased neurodegeneration markers, and lower cognitive performance, without evidence of strong nonlinear interactions.

However, the value of VAE lies not in global cluster separation but in revealing localized nonlinear structures not captured by linear projections. The VAE latent space showed a characteristic crescent-shaped trajectory for A+/T+/N+ individuals (Figure 7A, 15), suggesting that progression through severe AD pathology follows a curvilinear path in biomarker space. This crescent pattern suggests that early amyloid accumulation, subsequent tau propagation, and later neurodegeneration do not evolve independently but instead unfold along a coordinated, nonlinear path in biomarker space, with accelerating and interacting changes as pathology advances. This crescent□shaped trajectory may serve as a hypothesis□generating representation of potential disease staging, warranting future validation using longitudinal biomarker trajectories and multimodal imaging.

PCA, by maximizing global variance, effectively linearizes these relationships, flattening the crescent into a more uniform distribution. This increases silhouette scores, which reward global separation, but may obscure biologically meaningful curvature. In contrast, VAE preserves local neighborhood structure, retaining the curvature that may correspond to pathophysiological staging.[12]

#### Implications and Future Directions

The predominantly linear biomarker relationships observed here may reflect several factors: (1) the sample is enriched for early□stage pathology (87.6% cognitively normal), where biomarker interactions are expected to be simpler; (2) plasma biomarkers, while sensitive, may not capture the full multidimensional complexity observable with multimodal neuroimaging; and (3) cross□sectional data cannot reveal temporal dynamics, where nonlinear interactions may be more pronounced.

Future work integrating longitudinal biomarker change, multimodal neuroimaging (structural MRI, tauLPET), genetics (APOE, polygenic risk), and proteomic/metabolomic profiles may reveal stronger nonlinear interactions better suited to VAE’s representational capacity.[66] Moreover, VAE’s generative properties, its ability to sample from learned distributions, could support simulation of disease progression, synthetic data augmentation for predictive modeling, and detection of anomalous biomarker profiles.

In summary, PCA effectively captures the global structure of plasma biomarker variation, while VAE provides complementary insight into localized nonlinear patterns, particularly in individuals with more advanced pathology. Rather than competing approaches, PCA and VAE offer distinct and synergistic perspectives: PCA for dimensionality reduction and broad visualization, VAE for uncovering subtle pathophysiological dynamics.

### 4.6 Implications for Population-Based Research and Clinical Trials

Our findings have direct relevance for large-scale epidemiological studies and pragmatic clinical trial design:

**First, researchers should not assume equivalence between ATN and data-driven phenotypes.** Studies using ATN to define eligibility or outcomes should consider complementary data-driven analyses to ensure that important phenotypic heterogeneity is not overlooked. Conversely, studies employing unsupervised clustering should benchmark results against ATN to facilitate interpretation and comparability across cohorts.

**Second, the sensitivity of ATN profiles to cutoff selection** (demonstrated in our contingency analyses showing 14 observed profiles with varying prevalence) underscores the need for standardized, platform-specific cutoffs. While we applied literature-based thresholds derived from high-quality validation studies [32–35], these were established using different assay platforms (e.g., Simoa vs. Lumipulse) and populations (e.g., Swedish vs. U.S. cohorts). Direct calibration of cutoffs within HRS using longitudinal cognitive and neuroimaging outcomes would enhance classification accuracy and reduce misclassification bias.[22,58]

**Third, for clinical trial enrichment, our findings suggest a tiered strategy.** ATN profiles with strong predictive validity (e.g., A+/T+/N+ for rapid decline) could serve as primary inclusion criteria, ensuring biological relevance and regulatory alignment. Within ATN strata, data-driven clustering could identify high-risk subtypes for targeted recruitment. For example, among A+/T+/N+ individuals, Cluster 1 members with severe biomarker elevations might be prioritized for anti-amyloid or anti-tau therapies, while Cluster 2 members with intermediate profiles could be enrolled in prevention trials. Such precision enrichment could reduce sample size requirements and improve treatment effect detection.[7–9]

### 4.7 Longitudinal Predictive Utility

Both ATN profiles and data-driven clusters showed statistically significant but modest associations with cognitive decline over 4 years (R²=0.024 and 0.019, respectively). These effect sizes, while seemingly small, are consistent with the multifactorial etiology of cognitive aging, where biomarkers represent only one component among genetic susceptibility (e.g., APOE ε4), vascular burden, educational reserve, and lifestyle factors. The marginally superior performance of ATN for longer-term decline (4-year vs 2-year) suggests that categorical pathological constructs may better capture slow, cumulative processes, whereas continuous clustering may be more sensitive to acute or heterogeneous trajectories.

The limited variance explained underscores an important caveat: neither approach alone provides sufficient prognostic precision for individual-level prediction. Clinically actionable risk stratification will require integrating biomarkers with multimodal data (neuroimaging, genetics, cognitive trajectories) using advanced prediction models. Nonetheless, the significant longitudinal associations validate that both ATN and clustering capture pathologically relevant variation with clinical consequences.

### 4.8 Strengths and Limitations

#### Strengths

Key strengths include the population□representative HRS sample (N=4,465), enabling generalization beyond clinic□based cohorts; a comprehensive plasma biomarker panel with rigorous validation; multiple unsupervised methods (k□means, GMM, VAE) with convergent findings; extensive sensitivity analyses addressing methodological concerns (GFAP□excluded, k□selection, distance metrics, missing data); longitudinal cognitive data with 4Lyear follow□up; and direct quantitative comparison of theory□driven and data□driven approaches, which, to our knowledge, is among the first such analyses conducted in a U.S. nationally representative cohort using plasma biomarkers.

#### Limitations

Several important limitations warrant consideration. Clustering identifies statistical structure in biomarker space and does not, by itself, establish biological phenotypes without external validation. Future work incorporating imaging, CSF, or longitudinal clinical outcomes is needed to determine whether this subgroup reflects a biologically distinct entity.

**1. Lack of pathological validation:** We lacked gold□standard autopsy, amyloid□PET, or tau□PET confirmation to adjudicate the accuracy of plasma□based classifications. Although associations with cognition and convergence with PET□validated studies provide indirect support, the true sensitivity and specificity of our ATN profiles and clusters remain uncertain. Linking HRS biomarker data to the ADAMS autopsy cohort would directly address this limitation.
**2. Platform-specific cutoffs:** ATN cutoffs were derived from multiple assay platforms (Simoa for NfL/GFAP/pLtau181; IPLMS for Aβ42/40 ratio) and validated in heterogeneous cohorts (Swedish, German, U.S. clinical samples), introducing potential bias. Ideally, cutoffs should be established within HRS using longitudinal dementia outcomes or PET imaging. Sensitivity analyses (Supplementary Figure S8) show that 15**-**20% of individuals near decision boundaries would reclassify under plausible alternative thresholds. Because plasma biomarker cutoffs vary across assay platforms, our ATN classifications should be interpreted within the context of the specific analytic methods used in HRS.
**3. Cross-sectional clustering:** Clustering was performed on baseline biomarkers only. Individuals with identical baseline values but different rates of change (stable vs progressive) are treated equivalently, potentially obscuring dynamic phenotypes. Longitudinal clustering approaches (trajectory□based clustering, latent class growth analysis) would better capture temporal evolution.
**4. Survey weighting:** Clustering analyses used unweighted data due to incompatibility between survey weights and distance-based algorithms. While our analytic sample closely approximated weighted population distributions (Supplementary Table S9), cluster prevalences may not precisely generalize to the U.S. population. Advanced methods for incorporating survey weights into unsupervised learning (e.g., weighted distance metrics, post-stratification) warrant development.
**5. Small extreme clusters:** Clusters 1 (n=51, 1.2%) and 4 (n=14, 0.3%) had limited sample sizes, reducing statistical power for subgroup analyses. While bootstrap and distance-metric sensitivity analyses confirmed stability, replication in independent cohorts (e.g., Framingham Heart Study, UK Biobank) is essential to validate these rare but potentially important phenotypes.
**6. Cognitive assessment:** The HRS cognitive battery, while validated for dementia screening, is brief (27-point scale) and may have ceiling effects in high-functioning individuals, attenuating associations with biomarkers. More comprehensive neuropsychological batteries assessing domain-specific decline (memory, executive function, language) would provide richer phenotypic characterization.
**7. Missing incident dementia and survival data:** Although HRS provides longitudinal dementia adjudication and mortality follow-up, we did not include these outcomes because our primary objective was to characterize biomarker phenotypes rather than model long-term clinical endpoints. Incident dementia and survival analyses require additional methodological considerations (competing risks, time-varying covariates, informative censoring) beyond this study’s scope. Future work linking baseline ATN/cluster assignments to 10-15 year dementia incidence, mortality, and autopsy findings will establish long-term prognostic validity.
**8. Generalizability:** While HRS excludes institutionalized individuals (potentially underrepresenting advanced dementia) and racial/ethnic minorities remain somewhat underrepresented (13.6% Black, 8.4% Hispanic vs. 13.6% and 19.1% in 2020 U.S. Census), the study’s strengths for generalizability include: (1) national probability sampling with demographic weights; (2) close alignment of unweighted and weighted distributions (Table S9), validating cluster assignments; (3) biomarker distributions consistent with international cohorts using comparable assays; and (4) sufficient diversity to assess equity in biomarker performance. Findings should generalize well to U.S. community-dwelling older adults, though caution is warranted when extrapolating to clinical populations enriched for cognitive impairment or to international settings with different assay platforms[55].

### 4.9 Future Directions

Several avenues merit investigation. First, longitudinal analyses[22,56,61] examining whether ATN profiles or cluster memberships predict differential rates of clinical progression, conversion to dementia, or neuropathological outcomes would clarify which approach provides superior prognostic utility for specific endpoints. Second, external validation in independent cohorts (e.g., Framingham Heart Study, Rotterdam Study, UK Biobank) is essential to assess generalizability and robustness of the identified clusters. Third, multi-omics integration (genetics, proteomics, metabolomics) layered onto ATN and clusters could identify molecular drivers of phenotypic heterogeneity and novel therapeutic targets.[66] Fourth, methodological innovations, soft clustering with probabilistic memberships, longitudinal trajectory clustering, or causal discovery algorithms, could refine phenotyping beyond the static, hard-assignment approaches employed here. Fifth, therapeutic response studies examining whether clusters show differential responses to disease-modifying therapies (e.g., anti-amyloid antibodies) would establish clinical utility for data-driven phenotyping.

## 5. CONCLUSIONS

Theory□driven ATN classification and data□driven clustering offer complementary lenses for understanding Alzheimer’s disease**-**related pathology in population□based samples. ATN provides biologically grounded, clinically interpretable categories aligned with validated biomarker constructs, facilitating communication, regulatory approval, and mechanistic research. Unsupervised clustering discovers natural phenotypic gradients and rare subtypes, capturing quantitative variation and heterogeneity obscured by binary cutoffs. The modest concordance between approaches (ARI = 0.119, NMI = 0.113) reflects distinct but overlapping information, with approximately 11**-**15% shared and 85**-**89% unique to each method. Importantly, this modest concordance arises predominantly from GFAP’s dual biological role spanning amyloid□related inflammation and neurodegeneration; when GFAP is excluded, clustering and ATN become nearly independent (ARI = 0.03), confirming that binary categorization and continuous phenotyping capture fundamentally different biological structures even when applied to identical biomarkers.

Both frameworks showed significant associations with cognitive outcomes and longitudinal decline, validating their capture of pathologically relevant variation. Neither approach is categorically superior; rather, their integration promises a more comprehensive characterization of the multifaceted pathobiology underlying Alzheimer’s disease and related dementias. Future research should leverage both frameworks synergistically, using ATN for coarse stratification and benchmarking, and clustering for granular phenotyping and discovery, to advance precision medicine in dementia research and accelerate development of targeted interventions.

## Supporting information

https://github.com/efchea1/ATN-vs-Machine-Learned-Plasma-Biomarker-Phenotypes

https://github.com/efchea1/ATN-vs-Machine-Learned-Plasma-Biomarker-Phenotypes/tree/main/Figures/TIFF_Figures

## ACKNOWLEDGMENTS

The Health and Retirement Study (HRS) is sponsored by the National Institute on Aging (U01AG009740, R01AG030153) and conducted by the University of Michigan. I thank the HRS participants for their invaluable contributions to this nationally representative study, as well as the HRS field interviewers, biomarker laboratory teams, and data management staff for their essential roles in data collection, processing, and dissemination.

## AUTHOR CONTRIBUTIONS

E.F.C.: Conceptualization, Data Curation, Methodology, Formal Analysis, Software, Validation, Visualization, Writing **-** Original Draft, Writing **-** Review & Editing, Project Administration.

## ETHICS AND CONSENT TO PARTICIPATE

This study utilized de-identified, publicly available data from the Health and Retirement Study (HRS), a nationally representative longitudinal panel study conducted by the University of Michigan. The HRS and its Venous Blood Study component received approval from the University of Michigan Institutional Review Board (IRB).

All participants provided written informed consent for participation, biospecimen collection, and subsequent analyses prior to enrollment. The informed consent process included detailed explanation of study procedures, biomarker testing, data use, and participant rights.

All research procedures were conducted in accordance with the Declaration of Helsinki and followed institutional and federal guidelines for the ethical treatment of human participants, including compliance with 45 CFR 46 (Common Rule) regulations for the protection of human subjects. As this analysis utilized only de-identified, publicly available data, it qualified for exemption from additional IRB review under federal exemption category 4.

No additional consent was required for this secondary data analysis.

## DATA AVAILABILITY

HRS data are publicly available through the HRS website (https://hrs.isr.umich.edu) following registration and completion of a Restricted Data Use Agreement. All analysis code is included in the Supplementary Materials (ATN_Machine_Learned_Plasma_Biomarker_Phenotypes.Rmd). Additional project files and intermediate outputs are available at: https://github.com/efchea1/ATN-vs-Machine-Learned-Plasma-Biomarker-Phenotypes

