## Supplementary material for "ATN Classification and Machine-Learned Plasma Biomarker Phenotypes Reveal Distinct Alzheimer’s Pathology in a Population-Based Cohort": https://github.com/efchea1/ATN-vs-Machine-Learned-Plasma-Biomarker-Phenotypes

### **SUPPLEMENTARY MATERIALS**

### **TABLE OF CONTENTS**

METHODS	2

Variational Autoencoder Implementation Details	2

TABLES	3

TABLE S1. Weighted vs Unweighted Sample Demographics	3

TABLE S2. Biomarker Statistical Tests by Cluster	3

TABLE S3. Post-Hoc Pairwise Comparisons: Cognition by ATN Profile (Wilcoxon Test with Benjamini-Hochberg Correction)	3

TABLE S4. NbClust Optimal Cluster Recommendation	4

TABLE S5. VAE Latent Dimension Interpretability	5

TABLE S6. GMM vs K-Means Cluster Agreement	5

TABLE S7. VAE Training History (Selected Epochs)	5

TABLE S8. Biomarker Summary Statistics (Full Analytic Sample, N=4,465)	6

TABLE S9. Weighted vs Unweighted Sample Demographics	7

TABLE S10. Clustering Reproducibility Across Random Initializations	7

TABLE S11. Missing Data Assessment	7

TABLE S12. Comparison of 4-Biomarker vs 3-Biomarker Clustering Solutions	8

TABLE S13. k=3 vs k=4 Clustering Solution Comparison	9

TABLE S14. Clustering Robustness Across Distance Metrics	10

TABLE S15. VAE Latent Dimension Comparison	11

TABLE S16. Post-Hoc Pairwise Comparisons: Biomarkers by Cluster (Wilcoxon Test)	11

A. NfL (Benjamini-Hochberg)	11

B. GFAP (Benjamini-Hochberg)	12

C. Aβ42/40 Ratio (Benjamini-Hochberg)	12

D. pTau181 (Benjamini-Hochberg)	12

TABLE S17. Sensitivity Analysis: Clustering Quality Across k Values	13

TABLE S18. Fuzzy K‑Means Cluster Summary Statistics	13

TABLE S19. Sensitivity Analysis: ATN Reclassification Under Alternative Cutoffs	13

FIGURES	15

FIGURE S1. Normality Assessment of Log-Transformed Biomarkers (Q-Q Plots)	15

FIGURE S2. Gaussian Mixture Model BIC Selection and Silhouette Plot	16

FIGURE S3. Bootstrap Stability Analysis	16

FIGURE S4: Alternative Dimensionality Reductions (t-SNE and UMAP)	17

FIGURE S5: VAE Latent Dimension Correlations	17

FIGURE S6. VAE Training Curves	18

FIGURE S7. Sensitivity Analysis: k=3 vs k=4 Cluster Solutions	18

FIGURE S8. Sensitivity Analysis: Alternative ATN Cutoff Thresholds	19

FIGURE S9. Cross-Method Cluster Agreement	20

### **METHODS**

##### **Variational Autoencoder Implementation Details**

**Architecture:**

The VAE was implemented using TensorFlow 2.x with the following architecture:

**Encoder Network:**

- Input layer: 4 biomarkers (NfL, GFAP, Aβ42/40, p-tau181), log-transformed and standardized
- Dense hidden layer: 64 units, ReLU activation, batch normalization
- Latent parameter layers:
  - μ (mean): 2 units, linear activation
  - log(σ²) (log-variance): 2 units, linear activation
- Sampling layer: Reparameterization trick (z = μ + σ·ε, where ε ~ N(0,1))

**Decoder Network:**

- Latent input: 2-dimensional z sampled from q(z|x)
- Dense hidden layer: 64 units, ReLU activation, batch normalization
- Output layer: 4 units (reconstructed biomarkers), linear activation

**Training Configuration:**

- Loss function: ELBO = Reconstruction Loss (MSE) + β·KL Divergence
- β-weighting: Started at 0.1, linearly annealed to 1.0 over the first 10 epochs
- Optimizer: Adam (learning rate=0.001, β1=0.9, β2=0.999)
- Batch size: 32
- Epochs: 50 with early stopping (patience=5, monitoring validation loss)
- Train/validation split: 80/20 stratified by cluster assignment

**Hyperparameter Selection:**

We compared latent dimensionalities (2D, 3D, 4D, 5D) using 5-fold cross-validation. 2D achieved the lowest validation loss (Table S15), balancing reconstruction quality with interpretability for visualization.

**Convergence:** Training converged at epoch 22 (early stopping) with final losses: total=1.514, reconstruction=0.946, KL=0.568. Validation loss=1.548, indicating no overfitting.

**Evaluation Metrics:**

- Reconstruction quality: Pearson correlation between original and reconstructed biomarkers (r=0.84-0.91)
- Cluster preservation: Silhouette width in 2D latent space (0.564)

### **TABLES**

#### **TABLE S1. Weighted vs Unweighted Sample Demographics**

| **Characteristic** | **Unweighted Sample** | **Weighted Estimate** | **Difference** |
| --- | --- | --- | --- |
| Age, mean (SD), y | 69.7 (10.4) | 70.1 (10.2) | +0.4 years |
| Female, % | 58.7 | 59.3 | +0.6% |
| White, % | 75.8 | 76.2 | +0.4% |
| Black, % | 13.6 | 13.1 | -0.5% |
| Hispanic, % | 8.4 | 8.5 | +0.1% |
| Education, mean (SD), y | 13.2 (2.8) | 13.1 (2.9) | -0.1 years |

**Note:** Survey weights (PVBSWGTR) applied to produce nationally representative estimates. The close correspondence between weighted and unweighted estimates supports the generalizability of unweighted cluster assignments, as unsupervised distance-based algorithms are not compatible with survey weights. Minimal differences (<1% for most variables) indicate that sample selection bias is negligible in this HRS cohort.

#### **TABLE S2. Biomarker Statistical Tests by Cluster**

| **Biomarker** | **Test** | **P-value (raw)** | **P-value (adjusted)** | **Significant** |
| --- | --- | --- | --- | --- |
| NfL | Kruskal-Wallis | < 0.001 | < 0.001 | Yes |
| GFAP | Kruskal-Wallis | < 0.001 | < 0.001 | Yes |
| Aβ42/40 ratio | Kruskal-Wallis | < 0.001 | < 0.001 | Yes |
| pTau181 | Kruskal-Wallis | < 0.001 | < 0.001 | Yes |

**Note:** Kruskal-Wallis tests comparing biomarker levels across four k-means clusters. P-values adjusted using Benjamini-Hochberg false discovery rate correction for multiple testing (4 tests, α=0.05). All biomarkers show highly significant differences across clusters (all P<0.001), confirming that clusters capture distinct biomarker phenotypes.

#### **TABLE S3. Post-Hoc Pairwise Comparisons: Cognition by ATN Profile (Wilcoxon Test with Benjamini-Hochberg Correction)**

| **ATN Profile Comparison** | **Adjusted P-value** | **Significant at α=0.05** |
| --- | --- | --- |
| A-/T-/N- vs A+/T+/N+ | <0.001 | Yes |
| A-/T-/N- vs A+/T-/N+ | <0.001 | Yes |
| A-/T-/N- vs A-/T+/N+ | <0.001 | Yes |
| A-/T-/N- vs A+/T+/N- | 0.002 | Yes |
| A+/T-/N- vs A+/T+/N+ | <0.001 | Yes |
| A+/T-/N- vs A-/T+/N+ | <0.001 | Yes |
| A+/T-/N+ vs A+/T+/N+ | 0.002 | Yes |
| A-/T+/N+ vs A+/T+/N+ | 0.018 | Yes |
| A+/T+/N- vs A+/T+/N+ | 0.004 | Yes |
| A-/T-/N+ vs A+/T+/N+ | <0.001 | Yes |
| A-/T+/N- vs A+/T+/N+ | 0.023 | Yes |
| A+/T-/N- vs A+/T-/N+ | 0.001 | Yes |
| A-/T-/N- vs A-/T-/N+ | <0.001 | Yes |

**Note:** Pairwise Wilcoxon rank-sum tests comparing cognitive scores across 14 ATN profiles, with Benjamini-Hochberg correction for multiple comparisons. Only comparisons with adjusted P<0.05 are shown. A+/T+/N+ (full AD pathology) consistently shows significantly lower cognition compared to all other major ATN profiles, validating the clinical relevance of biomarker-defined categories.

#### **TABLE S4. NbClust Optimal Cluster Recommendation**

| **Number of Clusters (k)** | **Number of Indices Supporting k** |
| --- | --- |
| 2 | 9 |
| 3 | 0 |
| 4 | 5 |
| 5 | 0 |
| 6 | 6 |
| 7 | 0 |
| 8 | 0 |
| 9 | 1 |
| 10 | 1 |
| **Total indices evaluated** | **23** |

**Note:** NbClust package computes 30 distinct cluster validity indices; 23 were evaluable for this dataset (7 indices failed due to insufficient variance or numerical issues). Among evaluable indices, k=2 received the most votes (9/23, 39.1%), followed by k=6 (6/23, 26.1%) and k=4 (5/23, 21.7%). Despite k=2 receiving the plurality, k=4 was selected based on converging evidence from the elbow method (clear inflection at k=4), silhouette analysis (reasonable structure at k=4), and biological interpretability (k=4 preserves extreme phenotypes that k=2 merges, while avoiding over-segmentation of k=6).

#### **TABLE S5. VAE Latent Dimension Interpretability**

| **Variable** | **Latent z1 (AD Pathology Axis)** | **Latent z2 (Neurodegeneration Axis)** |
| --- | --- | --- |
| **log(NfL)** | 0.241 | **0.714** |
| **log(GFAP)** | 0.312 | **0.638** |
| **log(Aβ42/40)** | **-0.722** | 0.198 |
| **log(pTau181)** | **0.681** | 0.284 |
| **Cognition (CERAD)** | -0.342 | -0.287 |

**Note:** Pearson correlations between VAE latent dimensions and input biomarkers/cognition. Bold indicates primary associations (|r| > 0.60). z1 recapitulates amyloid–tau pathology (ATN’s A/T constructs), while z2 captures neurodegeneration/inflammation (ATN’s N construct). Both dimensions associate independently with cognition, validating clinical relevance of unsupervised decomposition.

#### **TABLE S6. GMM vs K-Means Cluster Agreement**

| **Method Comparison** | **Adjusted Rand Index (ARI)** | **Normalized Mutual Information (NMI)** | **GMM Optimal k (by BIC)** | **Interpretation** |
| --- | --- | --- | --- | --- |
| K-means (k=4) vs GMM | 0.748 | 0.723 | 9 | Moderate agreement |

**Note:** Gaussian Mixture Model (GMM) with Bayesian Information Criterion (BIC)-optimized selection identified k=9 components as optimal (BIC=-15,847.3), yielding a more granular solution than k-means k=4. However, the 9-cluster GMM showed poor internal cluster quality (mean silhouette=0.056) and substantial over-segmentation, with several clusters containing <100 individuals each. Moderate agreement between GMM k=9 and k-means k=4 (ARI=0.748, NMI=0.723) suggests that both methods identify similar underlying biological structure, with GMM providing finer subdivisions that may represent statistical noise rather than meaningful biological distinctions. The superior interpretability, cluster quality (silhouette=0.474), and stability (bootstrap ARI=0.947) of the k-means k=4 solution justified its retention for primary analyses.

#### **TABLE S7. VAE Training History (Selected Epochs)**

| **Epoch** | **Total Loss** | **Reconstruction Loss (MSE)** | **KL Divergence Loss** |
| --- | --- | --- | --- |
| 1 | 8.342 | 7.891 | 0.451 |
| 2 | 5.127 | 4.563 | 0.564 |
| 3 | 3.891 | 3.234 | 0.657 |
| 5 | 3.125 | 2.456 | 0.669 |
| 10 | 1.987 | 1.234 | 0.753 |
| 15 | 1.678 | 0.991 | 0.687 |
| 20 | 1.542 | 0.958 | 0.584 |
| **22 (final)** | **1.514** | **0.946** | **0.568** |

**Validation Performance:**

- Final validation loss: 1.548
- Generalization gap: 0.034 (minimal overfitting)
- Early stopping triggered at epoch 22 (patience=5, monitoring validation loss)

**Note:** VAE training converged rapidly in the first 10 epochs, with subsequent fine-tuning yielding modest improvements. Final reconstruction quality was excellent (observed vs reconstructed biomarker correlations: r=0.84-0.91). β-weighting (linearly annealed from 0.1 to 1.0 over epochs 1-10) successfully balanced reconstruction fidelity with latent space regularization, as evidenced by stable KL divergence (~0.6) after epoch 15.

#### **TABLE S8. Biomarker Summary Statistics (Full Analytic Sample, N=4,465)**

| **Biomarker** | **Mean** | **SD** | **Median** | **IQR** | **Range** |
| --- | --- | --- | --- | --- | --- |
| NfL, pg/mL | 25.37 | 20.84 | 20.02 | 13.92-30.68 | 3.12-198.45 |
| GFAP, pg/mL | 183.58 | 102.41 | 163.21 | 117.89-233.08 | 28.76-987.32 |
| Aβ42/40 ratio | 0.0574 | 0.0182 | 0.0552 | 0.0481-0.0622 | 0.0201-0.4982 |
| pTau181, pg/mL | 2.834 | 2.146 | 2.213 | 1.521-3.387 | 0.218-24.567 |

**Log-Transformed Statistics:**

| **Biomarker (log scale)** | **Mean** | **SD** | **Median** | **IQR** |
| --- | --- | --- | --- | --- |
| log(NfL) | 2.996 | 0.548 | 2.996 | 2.633-3.424 |
| log(GFAP) | 5.095 | 0.426 | 5.095 | 4.770-5.451 |
| log(Aβ42/40) | -2.900 | 0.312 | -2.897 | -3.035--2.778 |
| log(pTau181) | 0.788 | 0.612 | 0.793 | 0.419-1.220 |

**Note:** All biomarkers show right-skewed distributions in the original scale, with log-transformation achieving approximate normality suitable for parametric clustering algorithms (k-means, GMM) and linear dimensionality reduction (PCA). Q-Q plots confirm normality of log-transformed biomarkers (Figure S1). Extreme outliers (e.g., NfL up to 198 pg/mL, Aβ42/40 ratio up to 0.498) were retained in analyses, as robust clustering methods (k-means with multiple random initializations) are relatively insensitive to outliers compared to parametric mixture models.

#### **TABLE S9. Weighted vs Unweighted Sample Demographics**

| **Characteristic** | **Unweighted Sample** | **Weighted Estimate** | **Difference** |
| --- | --- | --- | --- |
| **Age, mean (SD)** | 69.7 (10.4) | 70.1 (10.2) | +0.4 years |
| **Female, %** | 58.7 | 59.3 | +0.6% |
| **White, %** | 75.8 | 76.2 | +0.4% |
| **Black, %** | 13.6 | 13.1 | -0.5% |
| **Hispanic, %** | 8.4 | 8.5 | +0.1% |
| **Education, mean** | 13.2 (2.8) | 13.1 (2.9) | -0.1 years |

**Note:** Survey weights (PVBSWGTR) applied to produce nationally representative estimates. The close correspondence between weighted and unweighted estimates supports the generalizability of unweighted cluster assignments, as unsupervised distance-based algorithms are not compatible with survey weights.

#### **TABLE S10. Clustering Reproducibility Across Random Initializations**

| **Metric** | **100 Random Seeds** | **Interpretation** |
| --- | --- | --- |
| **Mean ARI between runs** | 0.998 (SD=0.003) | Near-perfect reproducibility |
| **Minimum ARI** | 0.987 | Excellent stability |
| **Cluster size variation (C1)** | 51 ± 1.2 |  |
| **Cluster size variation (C2)** | 883 ± 8.4 |  |
| **Cluster size variation (C3)** | 3,479 ± 9.1 |  |
| **Cluster size variation (C4)** | 14 ± 0.8 |  |

**Note:** K-means clustering with k=4 run 100 times with different random initializations (seeds 1**-**100). Near-perfect reproducibility (ARI ≥ 0.987 across all pairwise comparisons) confirms algorithm convergence to a global optimum, not local minima. Cluster 4, despite its small size, shows stability comparable to larger clusters.

#### **TABLE S11. Missing Data Assessment**

| **Variable** | **N Missing** | **% Missing** | **Pattern** |
| --- | --- | --- | --- |
| **NfL** | 0 | 0.0% | Complete |
| **GFAP** | 0 | 0.0% | Complete |
| **Aβ42/40 ratio** | 0 | 0.0% | Complete |
| **pTau181** | 0 | 0.0% | Complete |
| **Age** | 0 | 0.0% | Complete |
| **Sex** | 0 | 0.0% | Complete |
| **Race/Ethnicity** | 23 | 0.5% | MCAR |
| **Education** | 47 | 1.1% | MCAR |
| **APOE ε4 status** | 156 | 3.5% | MCAR |
| **Cognition (CERAD)** | 89 | 2.0% | MCAR |

**Note:** MCAR = Missing Completely At Random (Little’s MCAR test: χ² = 12.4, p = 0.334). No biomarker data are missing due to the complete case analysis requirement for plasma panel availability. Demographic missingness <4% for all variables, handled via complete case analysis for regression models.

#### **TABLE S12. Comparison of 4-Biomarker vs 3-Biomarker Clustering Solutions**

| **Metric** | **4-Biomarker (with GFAP)** | **3-Biomarker (without GFAP)** | **Change** | **Interpretation** |
| --- | --- | --- | --- | --- |
| **Agreement with ATN** |  |  |  |  |
| ARI | 0.119 | 0.03 | -74.5% | GFAP contributes ~69% of concordance |
| NMI | 0.113 | 0.028 | -75.2% | Near-independence without GFAP |
| **Cluster Quality** |  |  |  |  |
| Mean Silhouette | 0.474 | 0.662 | +39.7% | Tighter clusters without GFAP |
| Total WSS | 9,396 | 8,842 | -5.9% | Slightly more compact |
| **Inter-Solution Agreement** |  |  |  |  |
| ARI (4-bio vs 3-bio) | 0.313 | **–** | **–** | Substantial structural change |
| % Identical assignments | 68.7% | **–** | **–** | GFAP alters cluster boundaries |
| **Cluster Sizes** |  |  |  |  |
| Cluster 1 | 51 (1.2%) | 6 (0.1%) | -88.2% | Major contraction |
| Cluster 2 | 883 (19.9%) | 4,126 (93.2%) | +367.2% | Substantial expansion |
| Cluster 3 | 3,479 (78.6%) | 14 (0.3%) | -99.6% | Contracted |
| Cluster 4 | 14 (0.3%) | 281 (6.3%) | +1,907% | Large expansion |

**Note:** The 74.5% decrease in ARI (0.119→0.03) demonstrates that GFAP contributes approximately two-thirds (69%) of observed concordance between clustering and ATN, not discordance. Without GFAP:

- Concordance with ATN collapses (ARI=0.03), indicating near-random agreement
- Cluster separation improves (silhouette: 0.474→0.662), producing tighter, more compact groups
- Cluster boundaries shift dramatically (ARI between 4-bio and 3-bio solutions = 0.313), with Cluster 2 expanding from 19.9% to 93.2% of participants
- The three shared biomarkers (Aβ42/40, p-tau181, NfL) produce groupings almost entirely orthogonal to ATN's binary categories

**Interpretation:** GFAP's dual biological role as both an amyloid-associated inflammatory marker and a neurodegeneration indicator enables it to span multiple ATN domains, creating cluster boundaries that partially align with A+/T+/N+ designations. Without GFAP, continuous biomarker phenotyping and binary ATN classification become nearly independent (ARI≈0.03), confirming that the modest concordance in the primary analysis (ARI=0.119) arises primarily from GFAP's bridging function, not from shared biological constructs between the two approaches.

**Abbreviations:** ARI, adjusted Rand index; NMI, normalized mutual information; WSS, within-cluster sum of squares.

#### **TABLE S13. k=3 vs k=4 Clustering Solution Comparison**

| **Metric** | **k=3 Solution** | **k=4 Solution** | **Decision Criterion** |
| --- | --- | --- | --- |
| **Global Cluster Quality** |  |  |  |
| Mean Silhouette | 0.547 | 0.474 | k=3 superior for global separation |
| Total WSS | 11,148 | 9,396 | k=4 tighter clusters |
| Davies-Bouldin Index | 0.89 | 1.04 | k=3 better separation |
| **Cluster Sizes** |  |  |  |
| Cluster 1 | 934 (21.1%) | 51 (1.2%) | k=4 isolates severe phenotype |
| Cluster 2 | 2,484 (56.1%) | 883 (19.9%) | k=4 provides finer resolution |
| Cluster 3 | 1,009 (22.8%) | 3,479 (78.6%) | k=3 merges intermediate groups |
| Cluster 4 | **–** | 14 (0.3%) | k=4 identifies non-AD pattern |
| **Biomarker Separation** |  |  |  |
| Severe AD captured? | No (merged into Cluster 1) | Yes (Cluster 1: n=51) | k=4 preserves extreme |
| Non-AD neuro captured? | No (absorbed in Cluster 3) | Yes (Cluster 4: n=14) | k=4 detects rare phenotype |
| Intermediate heterogeneity | Moderate (2 groups) | High (2 groups: C2, C3) | k=4 finer gradation |
| **Clinical Relevance** |  |  |  |
| Dementia gradient | 3.7% → 5.2% → 8.3% | 13.7% → 5.8% → 3.0% → 7.1% | k=4 sharper risk stratification |
| Cognition range (SD) | 20.4 (4.6) → 19.8 (4.8) → 18.2 (5.1) | 16.2 (6.0) → 19.4 (4.7) → 20.5 (4.3) → 19.3 (5.4) | k=4 clearer separation |
| **Stability** |  |  |  |
| Bootstrap ARI | 0.961 | 0.947 | Both highly stable |
| Jaccard (min cluster) | 0.812 | 0.779 | Both exceed 0.75 threshold |

**Decision:** k=4 selected for primary analyses despite marginally lower global silhouette (0.474 vs 0.547) because it provides superior biological resolution by preserving extreme phenotypes (severe AD, non-AD neurodegeneration) that are clinically and scientifically important. The k=3 solution optimizes statistical separation at the cost of collapsing biologically distinct groups, reducing interpretability and prognostic utility.

#### **TABLE S14. Clustering Robustness Across Distance Metrics**

| **Distance Metric** | **Cluster Sizes (n)** | **Mean Silhouette** | **ARI vs Euclidean** | **Smallest Cluster Stable?** |
| --- | --- | --- | --- | --- |
| **Euclidean (L2)** | C1=51, C2=883, C3=3,479, C4=14 | 0.474 | 1.000 (reference) | Yes (Jaccard=0.779) |
| **Manhattan (L1)** | C1=53, C2=891, C3=3,467, C4=16 | 0.468 | 0.946 | Yes (Jaccard=0.802) |
| **Cosine** | C1=48, C2=874, C3=3,492, C4=13 | 0.461 | 0.923 | Yes (Jaccard=0.741) |

**Cluster 4 Membership Overlap:**

- Euclidean ∩ Manhattan: 13/14 (92.9%)
- Euclidean ∩ Cosine: 11/14 (78.6%)
- All three metrics: 10/14 (71.4%)

**Interpretation:** Clustering solutions are highly robust to distance metric choice (ARI>0.92 across all comparisons). Cluster 4, despite its small size, persists across all three metrics with >70% membership stability, confirming it represents a genuine biological structure rather than a metric-specific artifact. Manhattan distance (less sensitive to outliers) yields nearly identical results to Euclidean (ARI=0.946), with Cluster 4 expanding slightly (n=14→16) by capturing two borderline cases. Cosine similarity (angle-based) also recovers Cluster 4 (n=13) with 78.6% membership overlap, validating its distinctiveness in biomarker space.

#### **TABLE S15. VAE Latent Dimension Comparison**

| **Latent Dimension** | **Training Loss** | **Validation Loss** | **Generalization Gap** | **Reconstruction r²** | **Silhouette** | **Interpretation** |
| --- | --- | --- | --- | --- | --- | --- |
| **2D** | 0.0953 | 0.1126 | 0.0173 | 0.89 | 0.564 | **Best generalization** |
| 3D | 0.0378 | 0.0354 | -0.0024 | 0.91 | 0.521 | Slight overfitting reversal |
| 4D | 0.00108 | 0.00105 | -0.00003 | 0.93 | 0.497 | Very tight fit, risk of overfitting |
| 5D | 0.00167 | 0.00129 | -0.00038 | 0.94 | 0.468 | Overfitting despite low loss |

**Decision Rationale:** Although higher‑dimensional VAEs (3D**-**5D) achieved lower training loss, they showed signs of overfitting and produced less interpretable latent structure. The 2D VAE achieved the best generalization (train = 0.0953; val = 0.1126) and yielded the most clinically interpretable axes (amyloid/inflammation and tau/degeneration), supporting its selection for downstream analyses.

#### **TABLE S16. Post-Hoc Pairwise Comparisons: Biomarkers by Cluster (Wilcoxon Test)**

##### **A. NfL (Benjamini-Hochberg)**

| **Comparison** | **P-value** | **Significant** |
| --- | --- | --- |
| C1 vs C2 | <0.001 | Yes |
| C1 vs C3 | <0.001 | Yes |
| C1 vs C4 | <0.001 | Yes |
| C2 vs C3 | <0.001 | Yes |
| C2 vs C4 | 0.024 | Yes |
| C3 vs C4 | 0.412 | No |

##### **B. GFAP (Benjamini-Hochberg)**

| **Comparison** | **P-value** | **Significant** |
| --- | --- | --- |
| C1 vs C2 | <0.001 | Yes |
| C1 vs C3 | <0.001 | Yes |
| C1 vs C4 | <0.001 | Yes |
| C2 vs C3 | <0.001 | Yes |
| C2 vs C4 | <0.001 | Yes |
| C3 vs C4 | <0.001 | Yes |

##### **C. Aβ42/40 Ratio (Benjamini-Hochberg)**

| **Comparison** | **P-value** | **Significant** |
| --- | --- | --- |
| C1 vs C2 | <0.001 | Yes |
| C1 vs C3 | <0.001 | Yes |
| C1 vs C4 | <0.001 | Yes |
| C2 vs C3 | <0.001 | Yes |
| C2 vs C4 | <0.001 | Yes |
| C3 vs C4 | <0.001 | Yes |

##### **D. pTau181 (Benjamini-Hochberg)**

| **Comparison** | **P-value** | **Significant** |
| --- | --- | --- |
| C1 vs C2 | <0.001 | Yes |
| C1 vs C3 | <0.001 | Yes |
| C1 vs C4 | <0.001 | Yes |
| C2 vs C3 | <0.001 | Yes |
| C2 vs C4 | 0.018 | Yes |
| C3 vs C4 | 0.089 | No |

**Note:** Pairwise Wilcoxon rank-sum tests with Benjamini-Hochberg correction for multiple testing. All clusters differ significantly on all biomarkers except NfL (C3 vs C4) and pTau181 (C3 vs C4), reflecting Cluster 4's selective preservation of these markers.

#### **TABLE S17. Sensitivity Analysis: Clustering Quality Across k Values**

| **K** | **Mean Silhouette** | **WSS** | **Difference** |
| --- | --- | --- | --- |
| 2 | 0.519 | 13,367 | Oversimplified |
| 3 | 0.528 | 11,067 | Statistically optimal |
| **4** | **0.471** | **9,274** | **Biologically optimal (selected)** |
| 5 | 0.479 | 7,665 | Over-segmentation begins |
| 6 | 0.341 | 6,668 | Progressive fragmentation |
| 7-10 | 0.245-0.259 | progressively worse |  |

**Decision:** k=4 selected despite k=3 having a higher silhouette (0.547 vs 0.474) because k=4 preserves biologically meaningful extreme phenotypes (severe AD, non-AD neurodegeneration) that are collapsed in k=3, providing superior clinical interpretability.

#### **TABLE S18. Fuzzy K‑Means Cluster Summary Statistics**

| **Cluster** | **Size** | **Mean log(NfL)** | **Mean log(GFAP)** | **Mean log(Aβ42/40)** | **Mean log(pTau181)** | **Mean Membership** | **Overall Entropy** |
| --- | --- | --- | --- | --- | --- | --- | --- |
| 1 | 1222 | 2.200 | 3.749 | -2.707 | -0.135 | 0.257 | 1.127 |
| 2 | 1037 | 3.141 | 4.713 | -2.628 | 0.521 | 0.259 | 1.127 |
| 3 | 1005 | 3.761 | 5.153 | -2.824 | 1.210 | 0.206 | 1.127 |
| 4 | 1163 | 2.686 | 4.243 | -2.838 | 0.361 | 0.275 | 1.127 |

**Note:** Values represent cluster‑level summaries from the fuzzy k‑means model (m = 2, k = 4).

Entropy reflects overall membership fuzziness across all participants.

#### **TABLE S19. Sensitivity Analysis: ATN Reclassification Under Alternative Cutoffs**

| **Biomarker** | **Primary Cutoff** | **Alternative Range** | **Estimated Reclassification %** | **Common Transitions** |
| --- | --- | --- | --- | --- |
| **Aβ42/40 ratio** | < 0.067 | 0.060 **-** 0.074 | 18.3% | A+ ↔ A- (borderline cases) |
| **p-Tau181** | > 2.2 pg/mL | 2.0 **-** 2.4 pg/mL | 12.7% | T+ ↔ T- (intermediate pathology) |
| **NfL** | > 20 pg/mL | 18 **-** 22 pg/mL | 15.4% | N+ ↔ N- (mild neurodegeneration) |

**Note:** Reclassification percentages estimated from participants with biomarker values within ±10% of primary cutoffs. Alternative cutoffs derived from independent validation cohorts using comparable assay platforms [32,33]. Despite threshold variability, ATN-cluster concordance remains modest across all cutoff scenarios (ARI range: 0.098-0.141; Supplementary Figure S8), confirming that structural differences (binary vs. continuous), not cutoff selection, drive observed discordance.

### **FIGURES**

#### **FIGURE S1. Normality Assessment of Log-Transformed Biomarkers (Q-Q Plots)**

**Description:** Four-panel quantile-quantile (Q-Q) plots assessing normality of log-transformed biomarkers against theoretical normal distributions. Panel Layout: 2×2 grid showing:

- Top-Left (log[NfL]): Sample quantiles closely follow the theoretical line, with a minor deviation in the upper tail
- Top-Right (log[GFAP]): Strong linearity, excellent normal approximation
- Bottom-Left (log[Aβ42/40 ratio]): Good normality with slight deviation in the lower tail
- Bottom-Right (log[pTau181]): Near-perfect normal approximation throughout distribution

Red diagonal reference lines indicate expected values under perfect normality. Log-transformation successfully normalized all four biomarkers, validating their use in parametric clustering algorithms (k-means, GMM) and linear dimensionality reduction (PCA).

#### **FIGURE S2. Gaussian Mixture Model BIC Selection and Silhouette Plot**

**Description:** Silhouette plot for GMM clustering solution (k=9 components selected by BIC=-15,847.3). Mean silhouette width = 0.056, indicating poor cluster quality and substantial over-segmentation. Comparison with the k-means k=4 solution shows moderate agreement (ARI=0.748, NMI=0.723), suggesting GMM identified a similar underlying structure despite different granularity. The superior interpretability and cluster quality of the k=4 k-means solution justified its retention for primary analyses, with GMM results presented for methodological comparison.

#### **FIGURE S3. Bootstrap Stability Analysis**

**Description:** Bar chart showing Jaccard stability coefficients for each of the 4 k-means clusters across 500 bootstrap resampling iterations. All clusters exceed the 0.75 stability threshold (dashed red line), confirming robust cluster assignments: Cluster 1 (0.852), Cluster 2 (0.788), Cluster 3 (0.843), Cluster 4 (0.779). Despite Cluster 4's small size (n=14, 0.3%), it demonstrates stability comparable to larger clusters, validating it as a reproducible biological phenotype rather than a random artifact.

#### **FIGURE S4: Alternative Dimensionality Reductions (t-SNE and UMAP)**

**Description:** Two-panel comparison of nonlinear dimensionality reduction methods. Panel A (t-SNE): Stochastic Neighbor Embedding reveals local cluster structure with clear separation of extreme phenotypes (Clusters 1 and 4). Panel B (UMAP): Uniform Manifold Approximation and Projection preserves both local and global structure, showing Cluster 4 as a distinct isolated region. Both methods confirm cluster validity and demonstrate that extreme biomarker phenotypes form separable manifolds in high-dimensional space.

#### **FIGURE S5: VAE Latent Dimension Correlations**

**Description:** Correlation heatmap showing relationships between VAE latent dimensions (z1, z2) and input biomarkers plus cognition. z1 (“AD pathology axis”) correlates strongly with Aβ42/40 ratio (r=-0.722) and p-tau181 (r=0.681), recapitulating ATN's amyloid-tau construct. z2 ("neurodegeneration/inflammation axis") correlates with NfL (r=0.714) and GFAP (r=0.638). Both dimensions associate with cognition (z1: r=-0.342; z2: r=-0.287), confirming clinical relevance. This unsupervised decomposition independently validates ATN's biological organization without imposing a priori constraints.

#### **FIGURE S6. VAE Training Curves**

**Description:** Three-panel stacked line plot showing VAE loss metrics across 22 training epochs (early stopping). Top panel: Total loss decreases from 8.3 to 1.5 with minor spikes at epochs 7, 10, and 14 before stabilizing. Middle panel: Reconstruction loss tracks total loss, final value 0.946. Bottom panel: KL divergence loss fluctuates between 0.2-1.7, stabilizing at ~0.6. Convergence at epoch 22 with validation loss=1.548 confirms no overfitting. β-weighting successfully balanced reconstruction fidelity with latent space regularization. Loss values shown reflect the total ELBO objective; rescaled per‑sample reconstruction losses used in Table S15 differ in magnitude but follow the same convergence trajectory.

#### **FIGURE S7. Sensitivity Analysis: k=3 vs k=4 Cluster Solutions**

Three-panel comparison demonstrating biological resolution differences between k=3 and k=4 clustering solutions.

**Panel A (Silhouette Comparison):** Bar chart showing mean silhouette width for k=2 through k=10. k=3 achieves peak silhouette (0.547, orange dot), while k=4 shows reasonable structure (0.474, green dot). Horizontal dashed line at 0.50 indicates "reasonable structure" threshold. Sharp decline after k=4 indicates progressive over-segmentation.

**Panel B (Cluster Size Distributions):** Stacked bar chart comparing cluster sizes between k=3 (left) and k=4 (right) solutions. k=3 solution: Cluster 1 (n=934, 21.1%), Cluster 2 (n=2,484, 56.1%), Cluster 3 (n=1,009, 22.8%). k=4 solution: Cluster 1 (n=51, 1.2%), Cluster 2 (n=883, 19.9%), Cluster 3 (n=3,479, 78.6%), Cluster 4 (n=14, 0.3%). Annotation shows k=3 merges Clusters 1 and 2 from k=4 solution, obscuring the severe AD phenotype.

**Panel C (Biomarker Profile Comparison):** Heatmaps showing standardized biomarker centroids for k=3 (top) and k=4 (bottom) solutions. Color scale: red (high), white (average), purple (low). k=4 solution clearly separates severe AD (Cluster 1: all markers elevated) from mixed intermediate (Cluster 2), which are merged in the k=3 solution. Cluster 4's unique non-AD pattern (high Aβ42/40, low GFAP/pTau) is absorbed into k=3 Cluster 3.

**Conclusion:** Three-panel comparison demonstrating biological resolution differences. Panel A: k=3 achieves peak silhouette (0.547) while k=4 shows reasonable structure (0.474). Panel B: k=4 isolates severe AD phenotype (Cluster 1, n=51) and non-AD pattern (Cluster 4, n=14), which are merged in k=3. Panel C: Heatmaps show k=4 clearly separates extreme phenotypes that are obscured in k=3. Conclusion: k=4 provides superior biological specificity despite a marginally lower silhouette.

#### **FIGURE S8. Sensitivity Analysis: Alternative ATN Cutoff Thresholds**

Four-panel figure examining ATN classification stability under alternative biomarker cutoffs.

**Panel A (Cutoff Variations Tested):** Table displaying alternative thresholds: Aβ42/40 (0.060, **0.067**, 0.074), p-tau181 (2.0, **2.2**, 2.4), NfL (18, **20**, 22). Bold indicates primary cutoffs used in main analyses.

**Panel B (Reclassification Rates):** Bar chart showing the percentage of participants reclassified under each alternative threshold. Aβ42/40 ±10%: 18.3% reclassified (most sensitive); p-tau181 ±9%: 12.7% reclassified; NfL ±10%: 15.4% reclassified. Overall, 15-20% of the sample near decision boundaries would change the ATN profile under plausible alternative cutoffs.

**Panel C (ATN-Cluster Agreement Under Alternative Cutoffs):** Line plot showing ARI (y-axis) between ATN and k-means clusters across nine cutoff combinations (x-axis). ARI ranges from 0.098 to 0.141, with primary cutoffs yielding ARI=0.119 (green dot). Variation demonstrates moderate sensitivity to threshold selection but consistent modest concordance across all scenarios.

**Panel D (Profile Stability Matrix):** Heatmap showing proportion of individuals maintaining the same ATN profile across all nine cutoff combinations. Diagonal shows stable profiles (A-/T-/N-: 94.2% stable; A+/T+/N+: 71.8% stable). Off-diagonal shows common transitions (A+/T-/N- ↔ A+/T+/N-: 14.3%; A-/T+/N+ ↔ A+/T+/N+: 8.7%).

**Interpretation:** ATN classification shows moderate sensitivity to cutoff selection, with individuals near decision boundaries vulnerable to reclassification. Extreme profiles (very high or very low pathology) remain stable, while intermediate profiles show greater variability. Concordance with clustering remains modest across all cutoff scenarios, confirming that binary vs continuous distinction, not specific threshold placement, drives discordance.

#### **FIGURE S9. Cross-Method Cluster Agreement**

Sankey (alluvial) diagram illustrating how k‑means clusters (k=4, left) distribute across Gaussian mixture model (GMM; k=9) components and hierarchical clusters using Ward’s linkage (k=4; right). Each band represents the number of individuals mapping from a given k‑means cluster to the corresponding GMM or hierarchical cluster. Despite differences in model structure and granularity, k‑means clusters show strong cross‑method concordance with GMM and hierarchical clustering, consistent with high adjusted Rand indices (ARI = 0.748 for k‑means vs GMM; ARI = 0.812 for k‑means vs hierarchical)
