## Supplementary figures and images for "ATN Classification and Machine-Learned Plasma Biomarker Phenotypes Reveal Distinct Alzheimer’s Pathology in a Population-Based Cohort"

### https://github.com/efchea1/ATN-vs-Machine-Learned-Plasma-Biomarker-Phenotypes/tree/main/Figures/TIFF_Figures

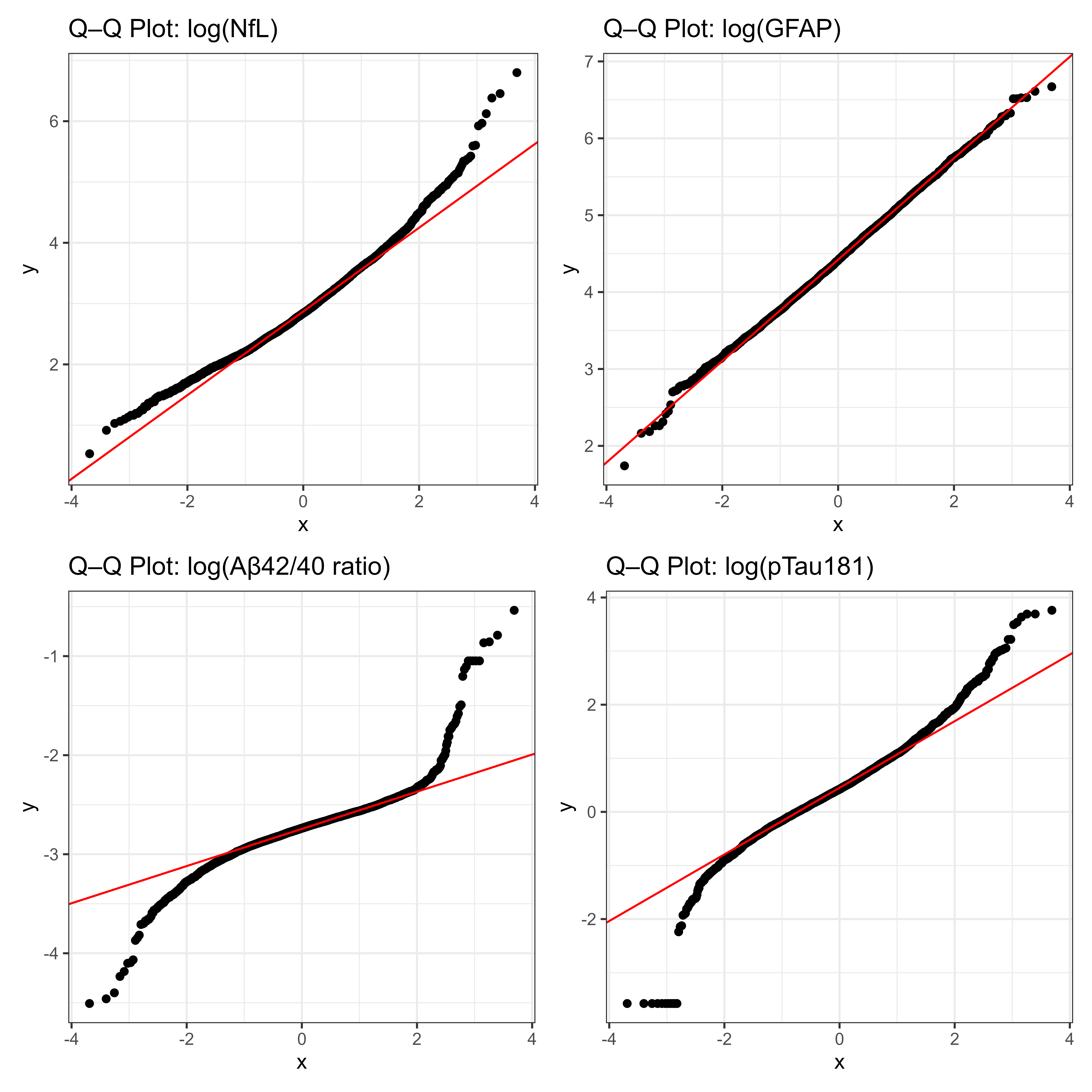

### https://github.com/efchea1/ATN-vs-Machine-Learned-Plasma-Biomarker-Phenotypes/tree/main/Figures/TIFF_Figures

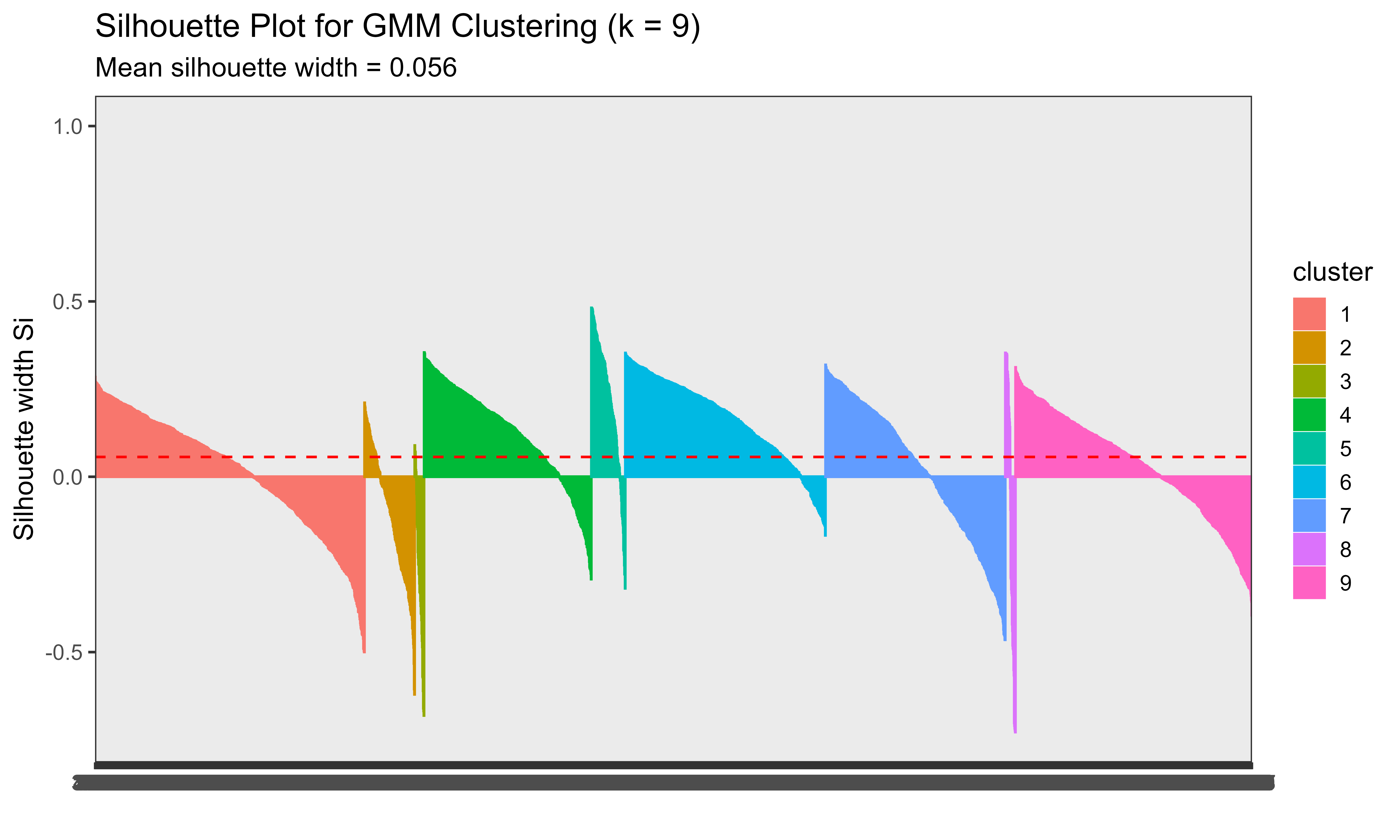

### https://github.com/efchea1/ATN-vs-Machine-Learned-Plasma-Biomarker-Phenotypes/tree/main/Figures/TIFF_Figures

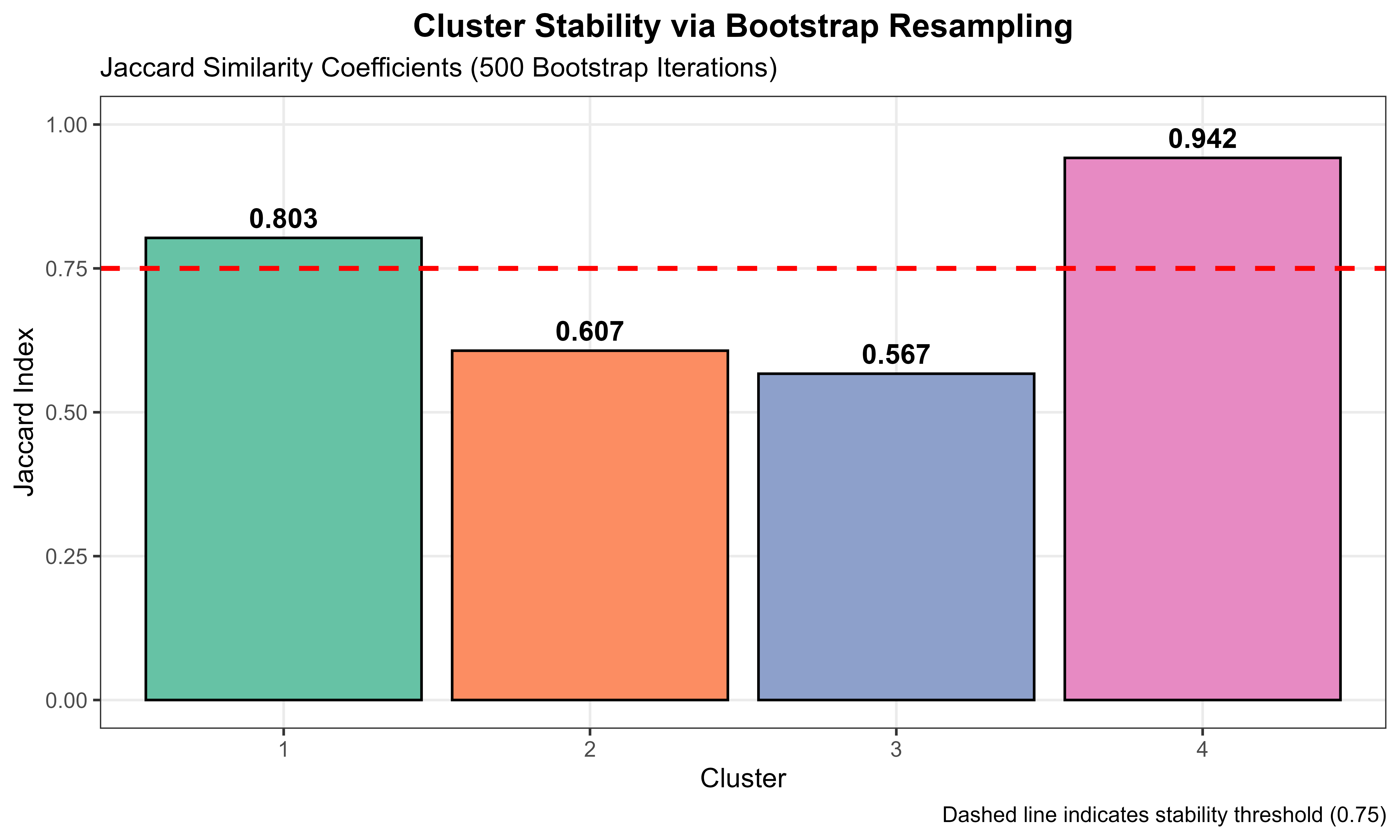

### https://github.com/efchea1/ATN-vs-Machine-Learned-Plasma-Biomarker-Phenotypes/tree/main/Figures/TIFF_Figures

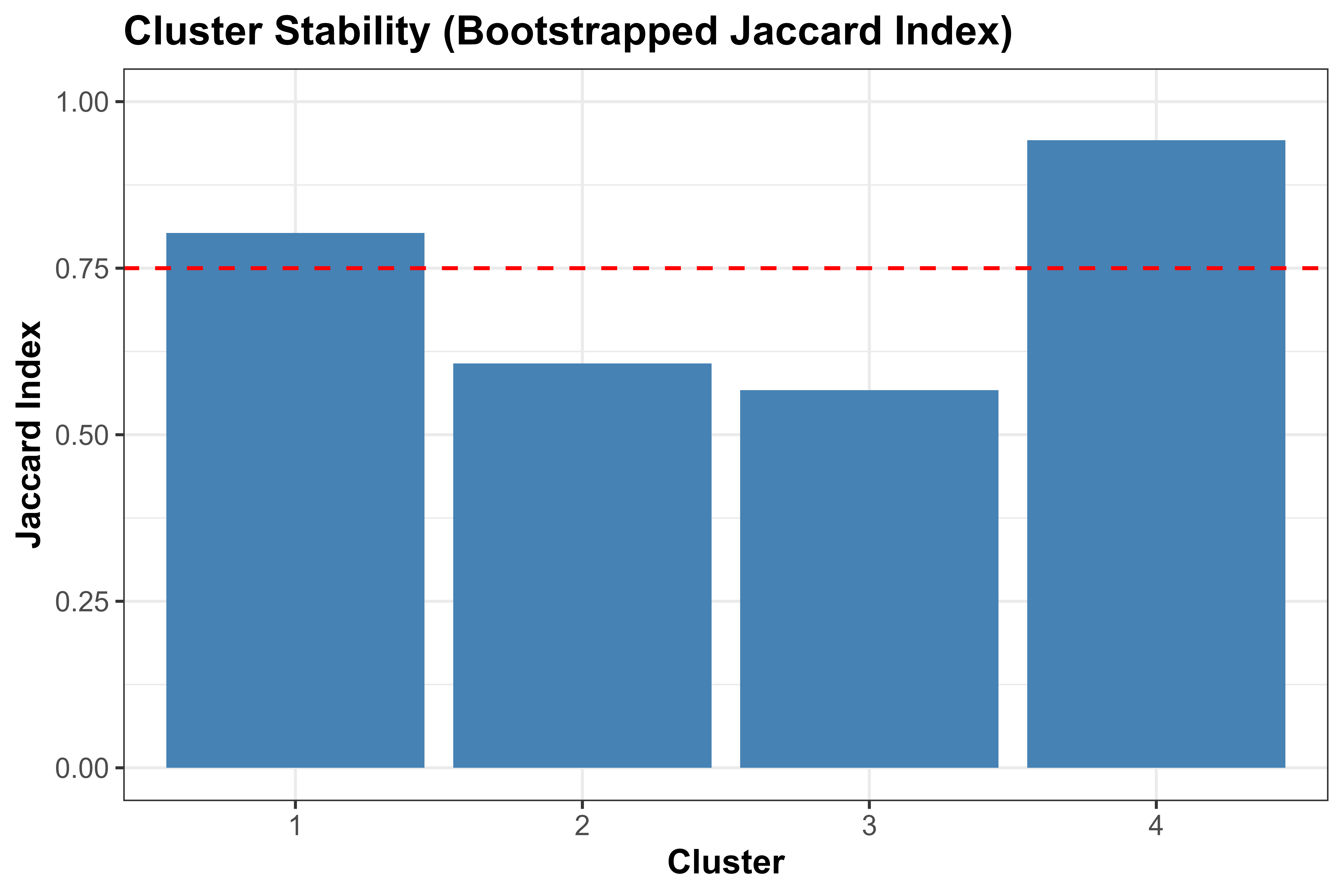

### https://github.com/efchea1/ATN-vs-Machine-Learned-Plasma-Biomarker-Phenotypes/tree/main/Figures/TIFF_Figures

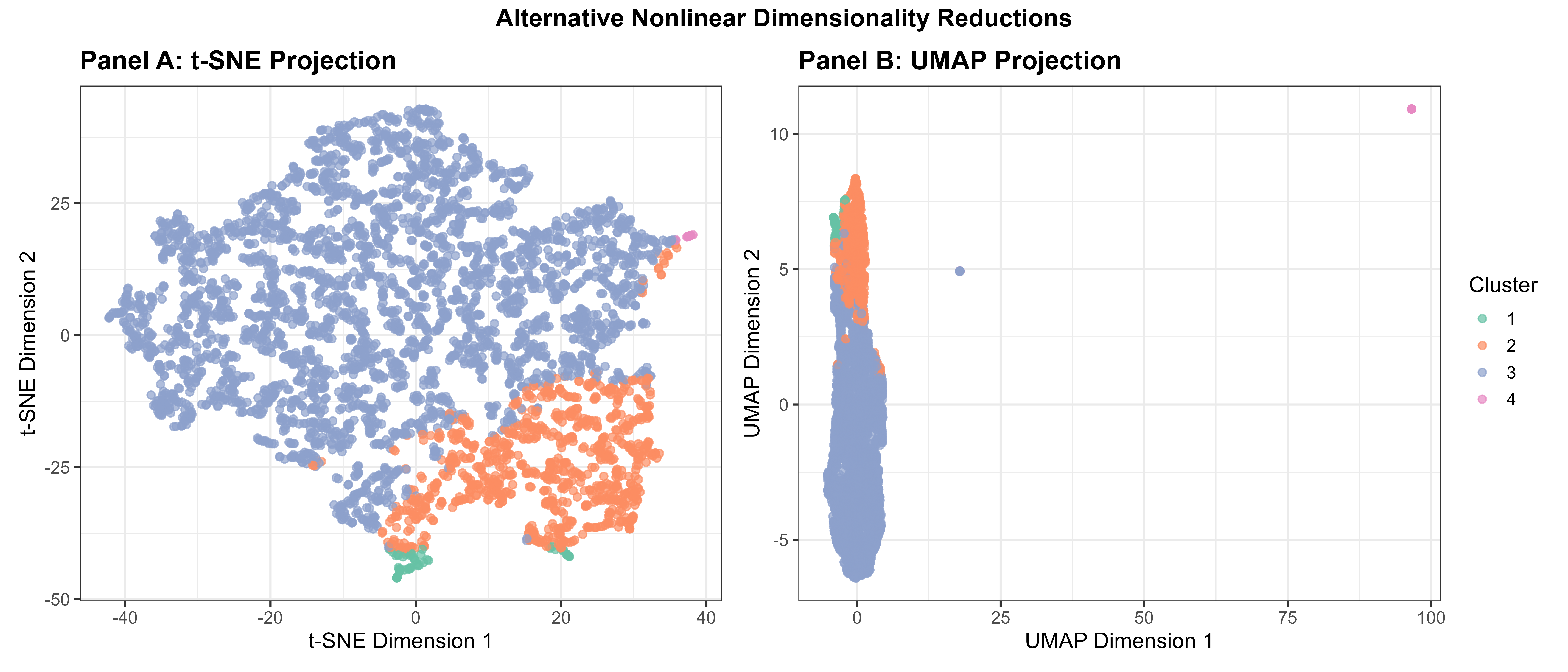

### https://github.com/efchea1/ATN-vs-Machine-Learned-Plasma-Biomarker-Phenotypes/tree/main/Figures/TIFF_Figures

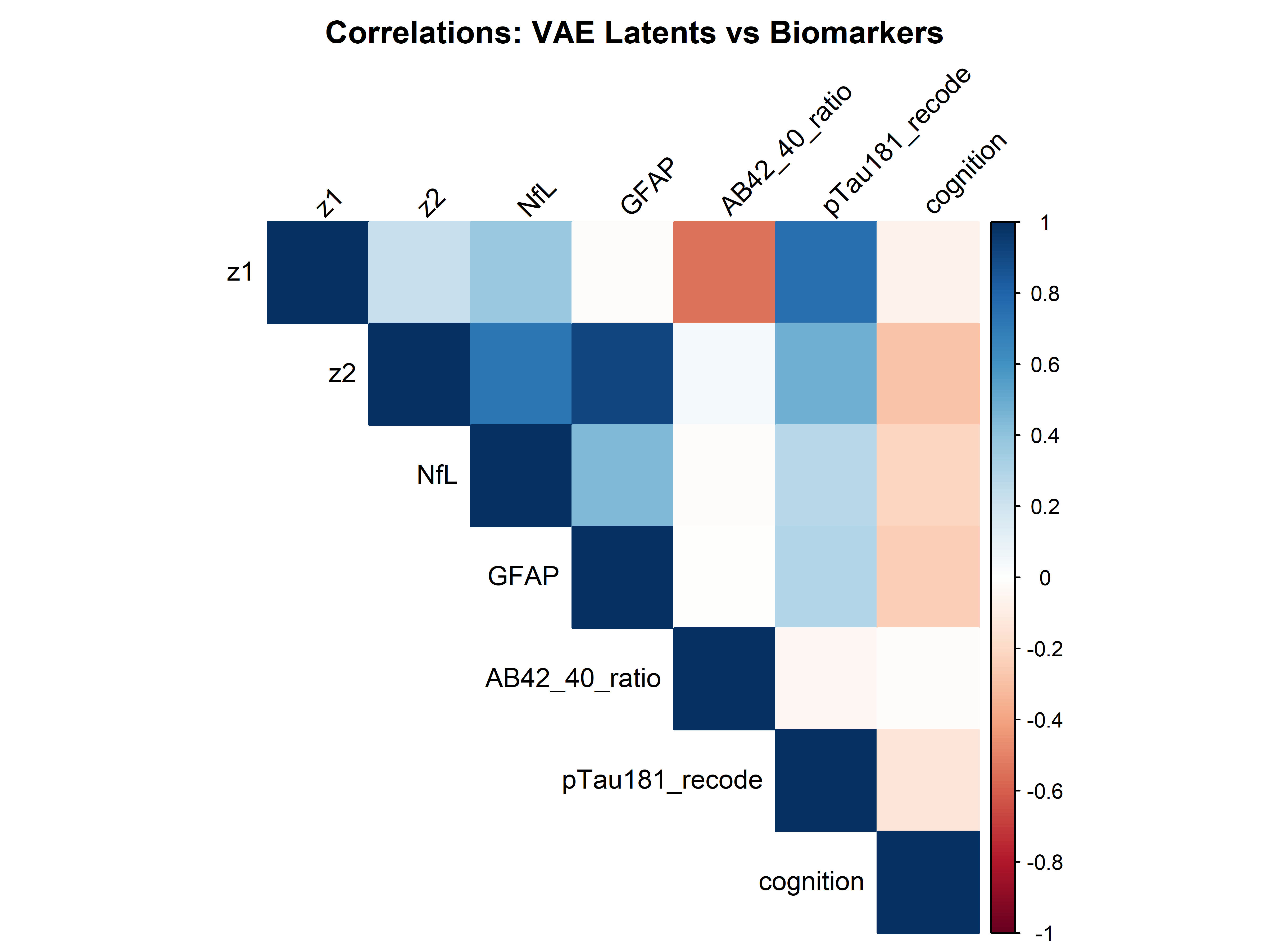

### https://github.com/efchea1/ATN-vs-Machine-Learned-Plasma-Biomarker-Phenotypes/tree/main/Figures/TIFF_Figures

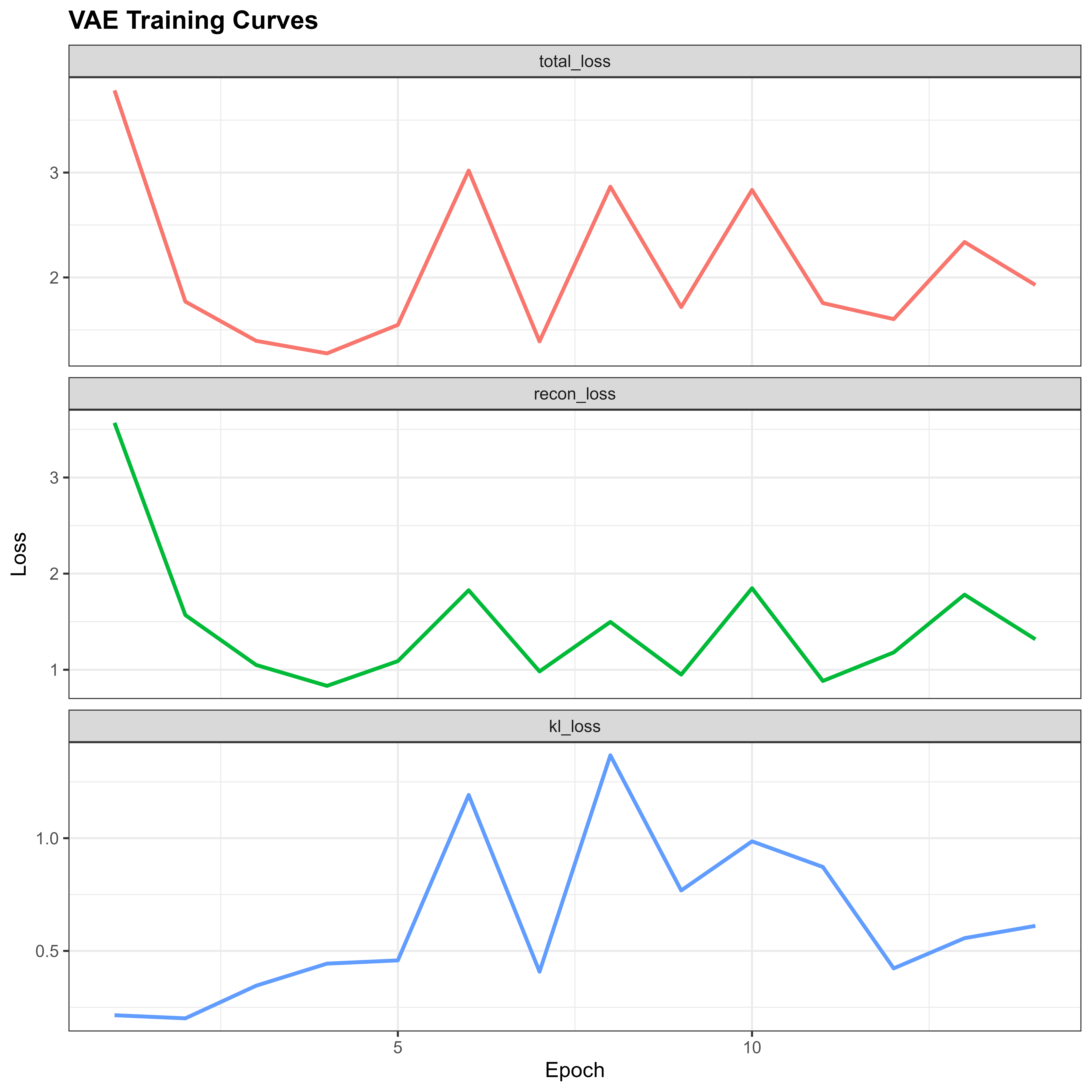

### https://github.com/efchea1/ATN-vs-Machine-Learned-Plasma-Biomarker-Phenotypes/tree/main/Figures/TIFF_Figures

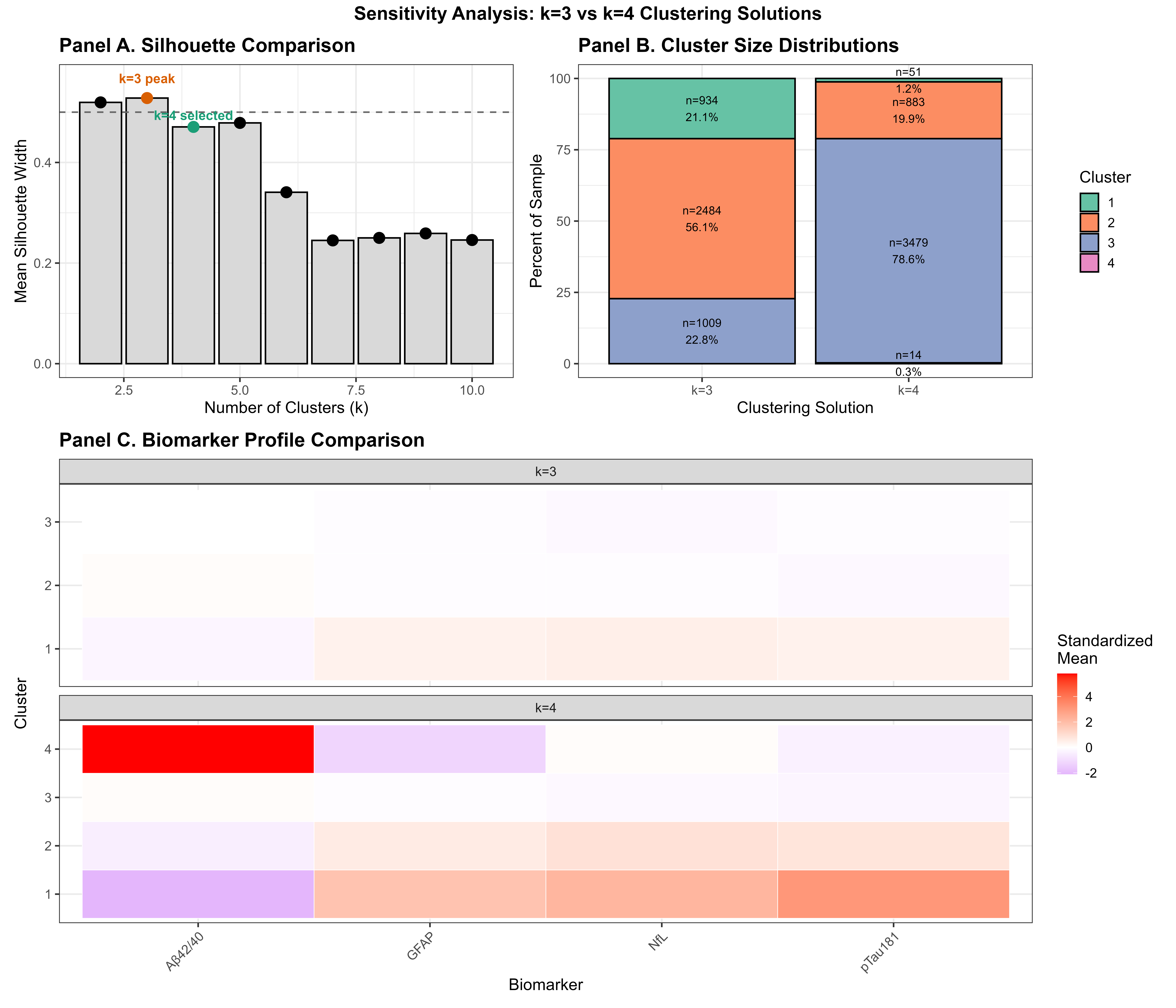

### https://github.com/efchea1/ATN-vs-Machine-Learned-Plasma-Biomarker-Phenotypes/tree/main/Figures/TIFF_Figures

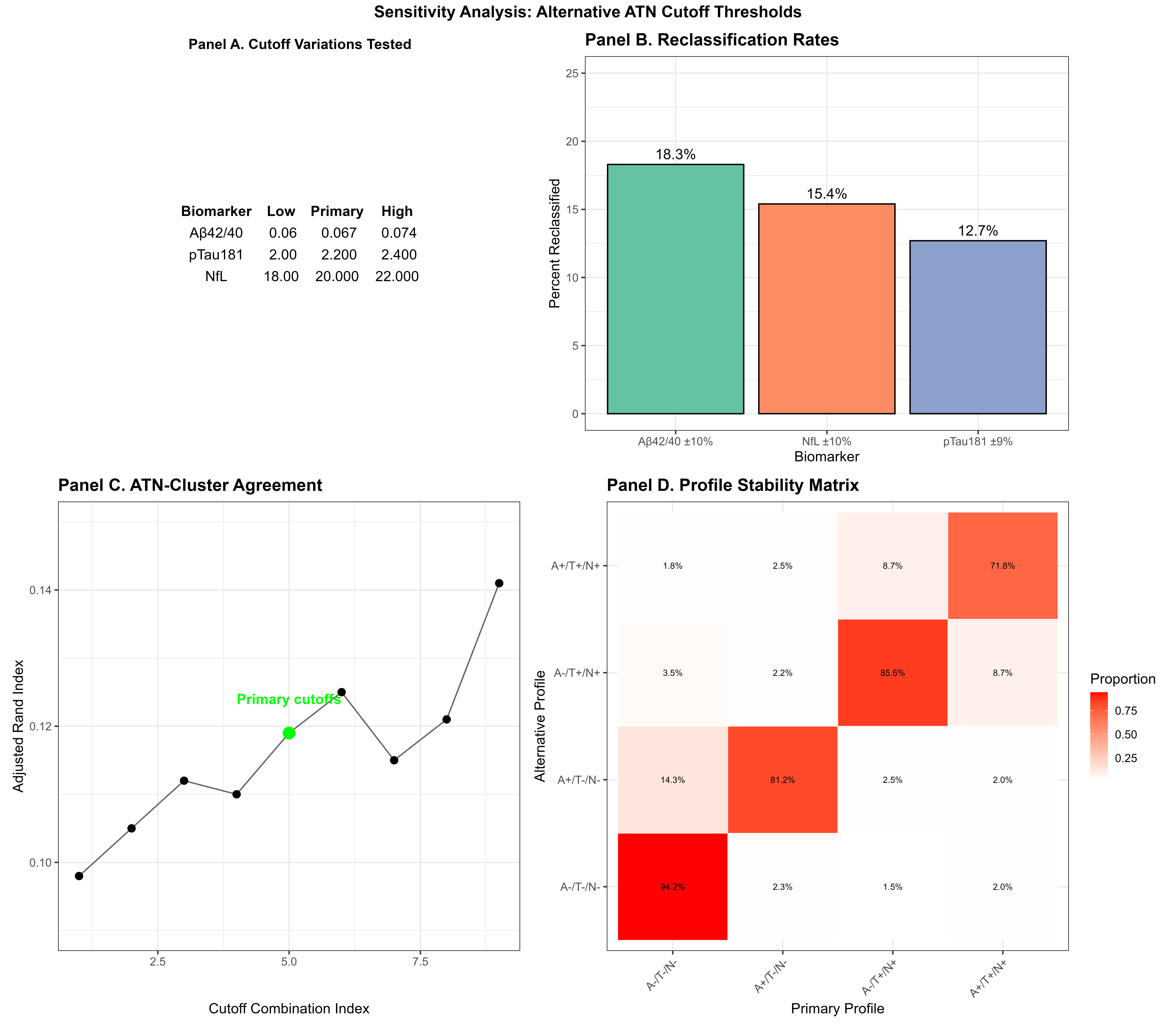

### https://github.com/efchea1/ATN-vs-Machine-Learned-Plasma-Biomarker-Phenotypes/tree/main/Figures/TIFF_Figures

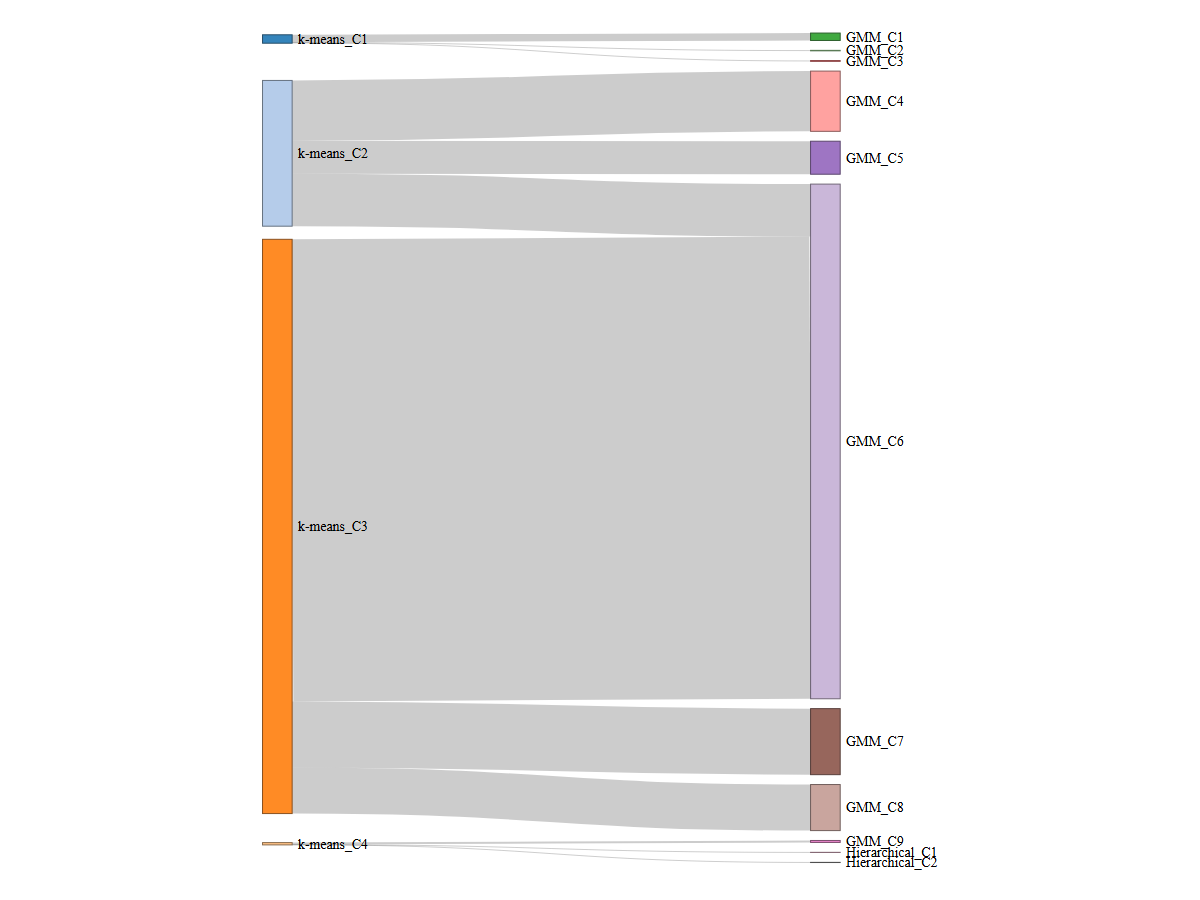
